# Assessing non-white ethnic participation in type 2 diabetes mellitus randomized clinical trials: A Meta-Analysis

**DOI:** 10.1101/2022.06.28.22275821

**Authors:** Rabeeyah Ahmed, Russell J. de Souza, Vincent Li, Sonia S. Anand

## Abstract

**Importance:** The prevalence of type 2 diabetes mellitus (T2DM) is increasing globally, and the greatest burden is borne by non-white ethnic groups. Randomized clinical trials (RCTs) provide evidence regarding the optimal medical therapy for the treatment of T2DM patients and inform national and international guidelines. However, there are concerns that the enrollment of ethnically diverse people into these trials is limited, which has resulted in a lack of ethnic diversity in RCTs of T2DM. Furthermore, the extent of underrepresentation may differ according to whether a trial is government-funded or industry-funded.

**Objective:** To systematically review and meta-analyze the proportion of non-white and white participants relative to their disease burden of T2DM included in large, influential government- and industry-funded RCTs of T2DM pharmacotherapies.

**Data Sources:** The PubMed electronic database was searched from January 2000 through January 2021.

**Study Selection:** Reports of RCTs of T2DM medications with a total sample size of at least 100 participants published in the year 2000 onwards, in high impact general medical journals (i.e., impact factor >10), were included.

**Data Extraction and Synthesis:** Data including the number of participants, proportion of participants by ethnicity, and funding sources, were extracted from trial reports.

**Main Outcomes and Measures:** The main outcome was the participation-to-prevalence ratio (PPR), which was calculated for each trial by dividing the percentage of white and non-white participants in the trial by the percentage of white and non-white participants with T2DM for the countries or regions of recruitment represented in each trial. A random-effects meta-analysis was used to generate the pooled PPR and 95% confidence intervals (CI) across study types. A PPR <0.80 indicates underrepresentation and >1.20 indicates overrepresentation.

**Results:** A total of 82 trials were included involving 296,964 participants: 14 were government-funded trials, and 68 were industry-funded trials. For government trials, the PPR for white participants was 1.11 (95% CI; 1.00-1.23) and for non-white participants was 0.73 (95% CI:0.62-0.86). Among industry trials, the PPR for white participants was 2.19 (95%CI: 1.91-2.50), and the PPR for non-white participants was 0.33 (95%CI: 0.29-0.38). Heterogeneity was high across all PPRs.

**Conclusions and Relevance:** Non-white participants are underrepresented in both government- and industry-funded T2DM trials, compared to white participants. The greatest disparity in ethnic diversity in RCTs is observed for industry-funded trials.

**Key Points:** *Question:* What is the representation of non-white participants in type 2 diabetes randomized clinical trials relative to their disease burden?

*Findings:* In this meta-analysis, non-white participants are underrepresented in both government-and industry-funded type 2 diabetes randomized trials, compared to white participants. The greatest disparity in ethnic diversity in randomized trials was observed for those funded by industry.

*Meaning:* Deliberate strategies to improve recruitment and enrolment of diverse participants proportional to the type 2 diabetes disease burden into industry and government-funded randomized controlled trials are needed to enhance the generalizability of research findings.

## Introduction

The burden of type 2 diabetes (T2DM) is disproportionately higher in non-white ethnic groups compared to white individuals.^1^ However, individuals from non-white ethnic groups are generally underrepresented in clinical trials.^1^ Randomized clinical trials (RCTs) are designed to produce the most reliable evidence on the efficacy and safety of various interventions.^2^ As such, they inform treatment recommendations and prevention strategies for T2DM. Underrepresentation of non-white individuals in RCTs can limit the generalizability and uptake of the trial findings among the groups with the highest disease burden.^3^

In some countries like the United States, the National Institutes of Health (NIH) provides guidelines for the inclusion of minority groups in NIH-funded clinical research,^4^ a measure taken to improve the generalizability of research findings. Industry-funded trials do not have the same requirements as the NIH, nor do many other government-established funding agencies.^5–8^ For example, Canada and Australia do not have guidelines for the recruitment of diverse populations into clinical trials. The “Guidance for Industry: Standards for Clinical Trials in Type 2 Diabetes in Canada” (2007) document does not mention the terms ‘race’ or ‘ethnicity,’ nor does it provide guidelines on participant recruitment.^7^ Similarly, the Government of Australia’s guidelines for clinical trials do not include any regulations pertaining to ethnic recruitment.^8^ In the United Kingdom (UK), there is also no requirement to record and report ethnicity in research studies.^5^

The NIH requirements are guided by the Public Health Service Act sec. 492B, 42 U.S.C. sec. 289a-2 and are designed to enhance the inclusion of minority groups in NIH-funded research.^4^ Efforts to increase representation of minority groups in RCTs have been made internationally as well. For example, some research institutions in the UK are taking measures to address the lack of ethnic participant recruitment guidelines for clinical trials. In 2018, the UK’s National Institute for Health Research (NIHR) launched the INCLUDE project, which was designed to improve the diversity of research participants in clinical studies.^5^ The NIHR also released new guidelines in 2020 in response to the COVID-19 pandemic. While these guidelines are intended to promote improvements in research practices related to COVID, the guidelines may also help improve the inclusion of diverse groups in other research areas.^5^

We conducted a systematic review of RCTs testing at least one T2DM pharmacotherapy from January 2000 to January 2021 and investigated: i) the non-white ethnic participation relative to the disease burden of T2DM in large RCTs in which T2DM therapies were evaluated, and ii) the differences in non-white ethnic participation comparing industry and government-funded trials.

## Methods

The systematic review was developed in accordance with the Preferred Reporting Items for Systematic Reviews and Meta-analyses (PRISMA) reporting guidelines.

### Data Sources and Searches

Guided by a clinical expert, a broad search strategy using medical subject heading (MeSH) terms and keywords related to T2DM randomized trials with relevant search terms (eTable 1 in the Supplement). Initially, the search was limited to trials published in “top-tier journals”, defined as an impact factor >/=30, which included the New England Journal of Medicine, The Lancet, Journal of the American Medical Association, and British Medical Journal. This criterion was kept for industry trials; however, it was expanded to include publications from three journals (Circulation, Diabetes Care, and Annals of Internal Medicine) with an impact factor ≥10 for government trials because few government-funded trials were published in journals with an impact factor ≥30. The search was also initially limited to trials that recruited from at least two countries. This criterion was kept for industry trials; however, it was removed for government trials because government-funded trials are often conducted solely within the country that funded the research. A complete list of study selection criteria can be found in eTable 2 in the Supplement. Two reviewers independently searched the PubMed database from January 1, 2000 to January 1, 2021 to identify relevant studies. PubMed was used because top journals are all indexed in the database, which eliminated the need for multiple database searching. After removing duplicates, the records were reviewed independently by one reviewer at the title and abstract level, followed by full-text screening by two reviewers in duplicate based on predetermined study selection criteria. Discrepancies were resolved by discussion with the senior author (SSA). Covidence (https://www.covidence.org/) was used for data management.

### Data Extraction

Three reviewers independently abstracted the following information from published studies: title, year of publication, journal, funding source, pharmacotherapy intervention, total number of participants, number of white and non-white participants, number of female participants, and country or region of greatest participant recruitment. Discrepancies were cross-checked, and where necessary, were resolved by discussion with the senior author (SSA). The resource ClinicalTrials.gov and other publicly available web resources, were consulted to fill in any missing information that was not in the main articles or supplementary materials.

### Outcome Measures

There were two outcomes of interest for this systematic review: the proportion of white participants in government and industry-funded trials relative to the T2DM disease burden in the population, and the proportion of non-white participants in government and industry-funded trials relative to the T2DM disease burden in the population.

### Statistical Analysis

The participation-to-prevalence ratio (PPR) metric was used to estimate the representation of white participants, and the representation of non-white ethnic participants compared to their respective disease burden, separately in industry and government-funded trials. The PPRs for white participants and non-white participants in each trial was calculated using the respective formulas below.

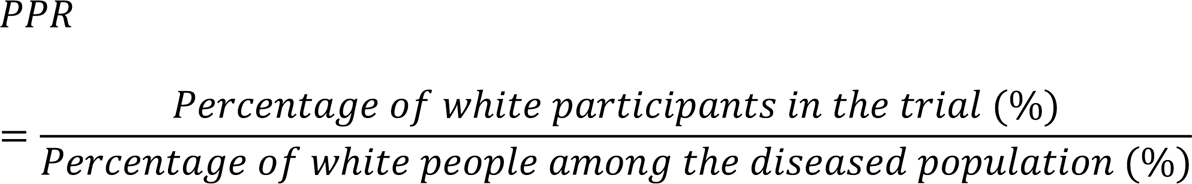

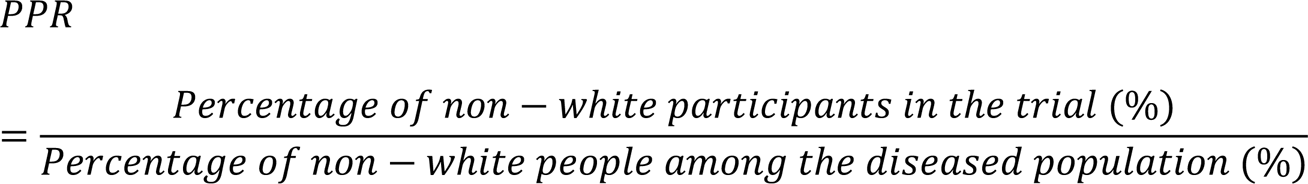

A PPR of 1 suggests that the specific ethnic group makes up the same proportion of participants in the trial as of the total cases in the countries from which the trial recruited, i.e., 80% are non-white participants in the trial and 80% of the cases of diabetes in the country are in non-white ethnic groups. For this review, an ethnic group is considered to be underrepresented when the PPR is <0.80, and overrepresented when the PPR is >1.20. This assessment is consistent with a 2020 study evaluating the participation of women in cardiovascular RCTs.^9^

The numerator for the PPR was known for each trial. The denominator, however, was calculated for each trial using prevalence and demographic data. The denominator of the PPR represents the proportion of white and non-white people with T2DM based on prevalence and population data for the country or region. If these data were not available, we used an estimate of 14.8% white people and 85.2% non-white people as described in detail below. The proportion of non-white and white people worldwide was determined by assuming that all people in Europe, Canada, the United States, and Australia were white; this resulted in an estimate of 14.8% white people and 85.2% non-white people. By estimating the “expected” cases for white people and non-white people, it is possible to calculate a PPR using the observed number of white and non-white participants in a trial and comparing this to the number one would expect to see if the trial recruited participants in proportion to the diabetes prevalence in ethnic groups for the country/region from which the trial recruited the greatest number of people. Appendix A provides a detailed explanation of PPR calculations. A list of estimates used for the PPR calculations is provided in eTable 4 in the Supplement.

Once the PPRs for white and non-white people were calculated for each trial, a random-effects meta-analysis was used to pool the individual-study PPRs and compute the overall 95% confidence intervals for the pooled PPRs. The random-effects model was used for this analysis because it provides appropriately wider confidence intervals and study weights in the presence of heterogeneity, which we expected to see across trial conditions and countries of conduct.

Because this review is not focused on the clinical outcomes of the trials, but rather on the recruitment strategy, the random-effects estimate is preferable. Under this model, it can be assumed that the true PPR may differ according to the setting, country, or type of trial.

We conducted a sensitivity analysis by perturbing the 14.8% white and 85.2% non-white assumption when data on expected prevalence in studies for which this was not available. We systematically altered the prevalence of non-whites to values from 80 through 90% by increments of 5%, and of white participants from 10% through 20% by increments of 5%.

All study-specific PPR estimates were calculated in Microsoft Excel, and the pooled PPRs across studies along with the 95% confidence intervals were calculated using Review Manager version 5.4 (RevMan).

## Results

Of the 573 records that were assessed for eligibility following removal of duplicates, 82 RCTs with a total of 296 964 participants were included (eFigure 1 in the Supplement). The list of included RCTs is provided in Table 2. The RCTs that were excluded after full-text review are provided in eTable 3 in the Supplement. Of the 82 trials included in the review, 82.9% were sponsored by industry,^10–77^ and 17.1% were government-funded,^78–91^ and most studies (63.4%) were published between the years 2011-2020. The proportion of non-white participants in this set of trials increased significantly from 10.9% in the years 2000-2005 to 23.8% between 2006-2011 and remained relatively constant between 2006-2020. Over one-third of the studies in the review (39.0%) recruited the greatest number of participants from the United States, and the most proportional recruitment of non-white populations occurred in trials that recruited primarily from North America (31.9%) and the United States (26.4%). The most common pharmacotherapy interventions were from the GLP-1 agonist, SGLT-2 inhibitor, and DPP-4 inhibitor classes. The percentage of non-white participants across all trials was 23.4%. Government-funded trials had a higher overall percentage of non-white participants (25.3%) than industry-funded studies (23.1%) (Table 1).

**Table 1.**
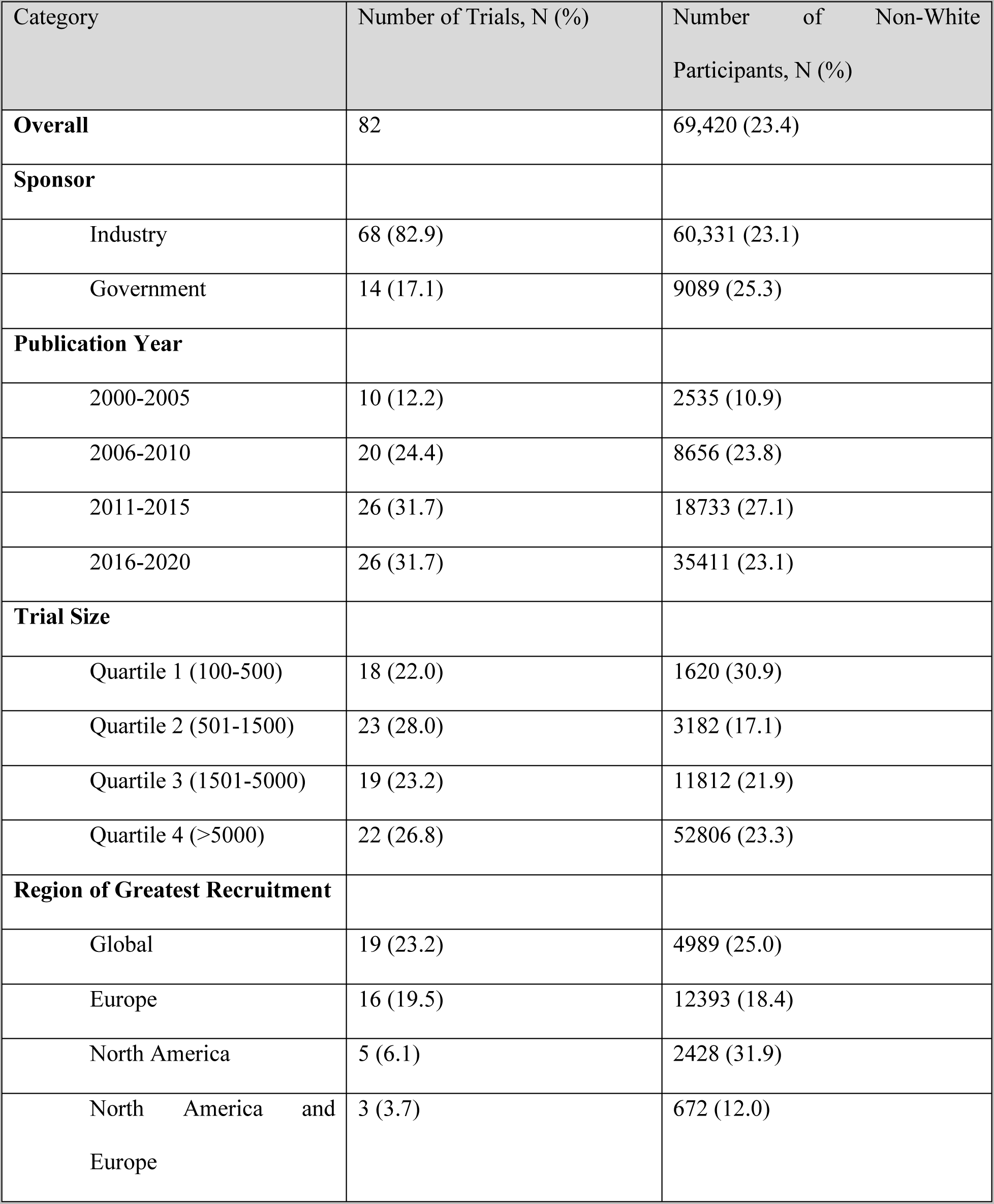
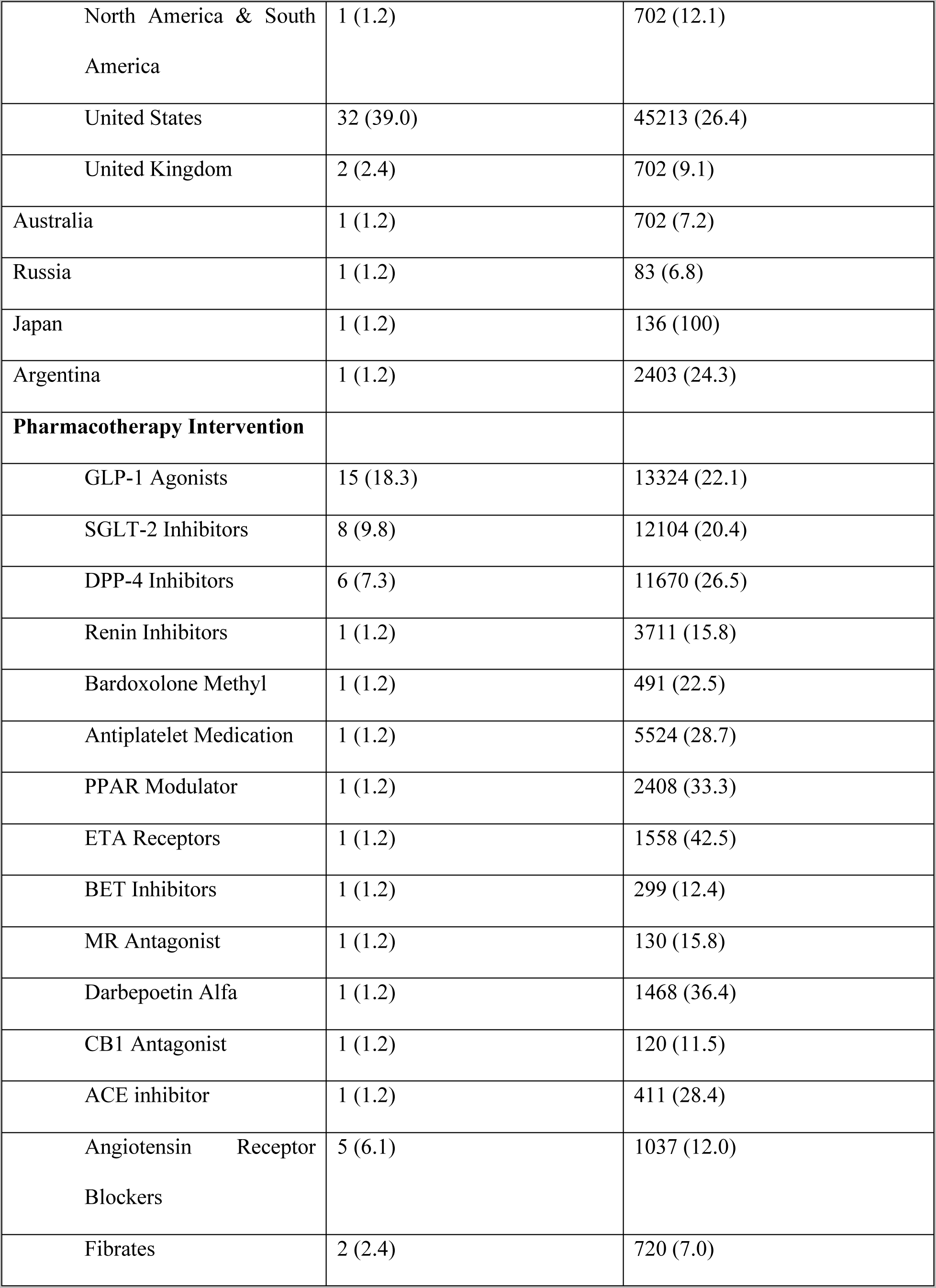
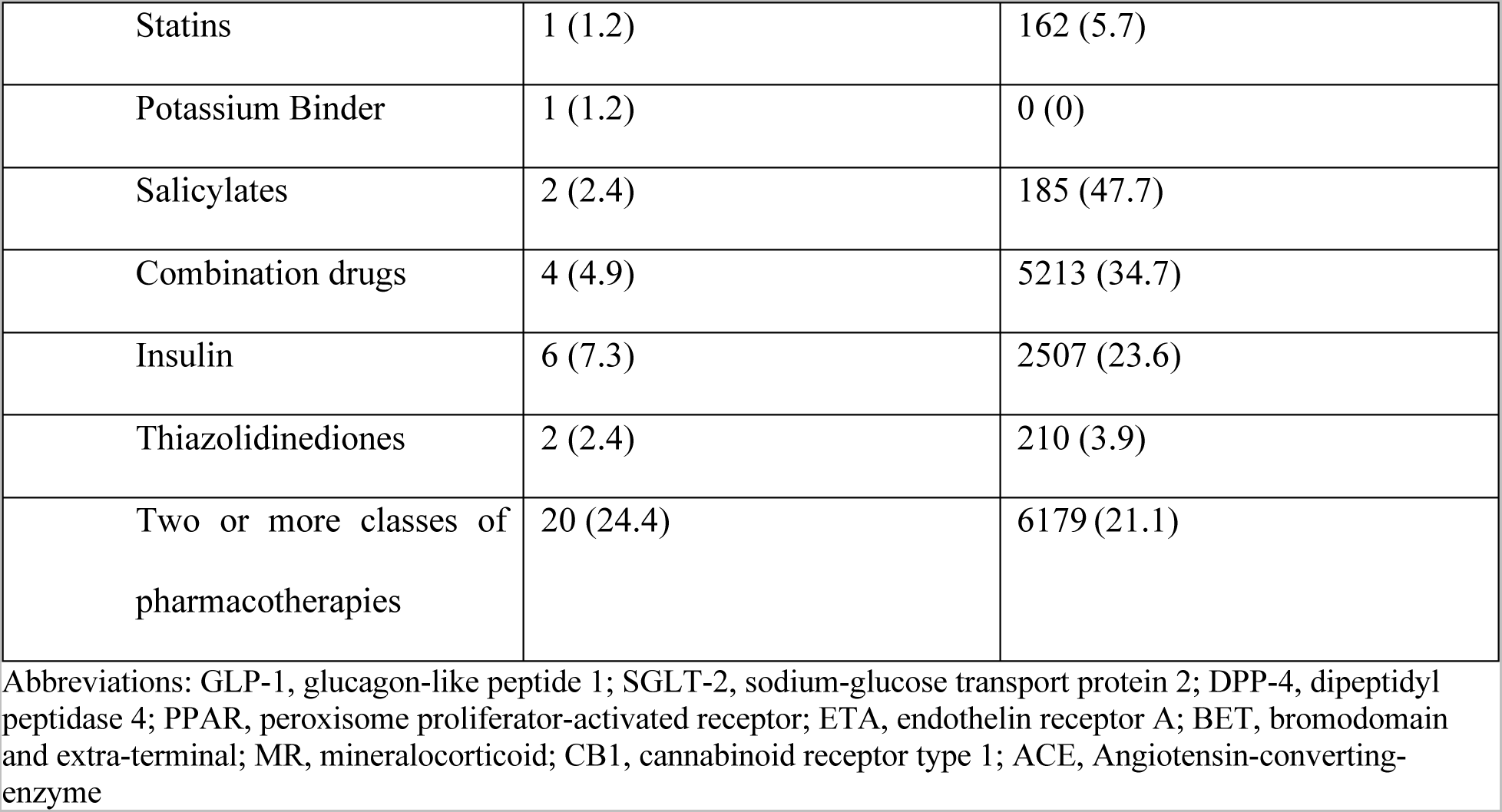
Baseline Characteristics of the 52 Clinical Trials.

**Table 2.**
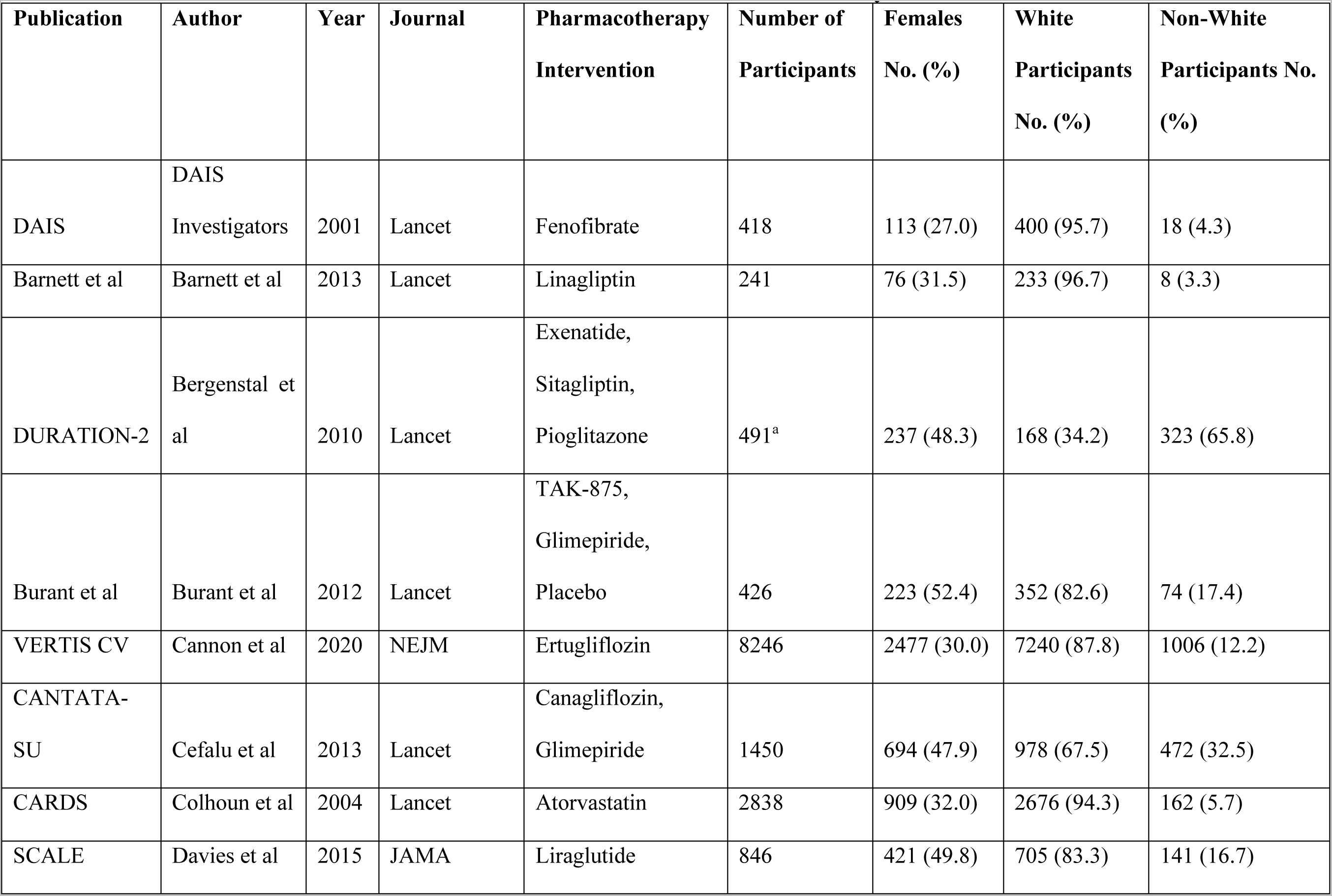
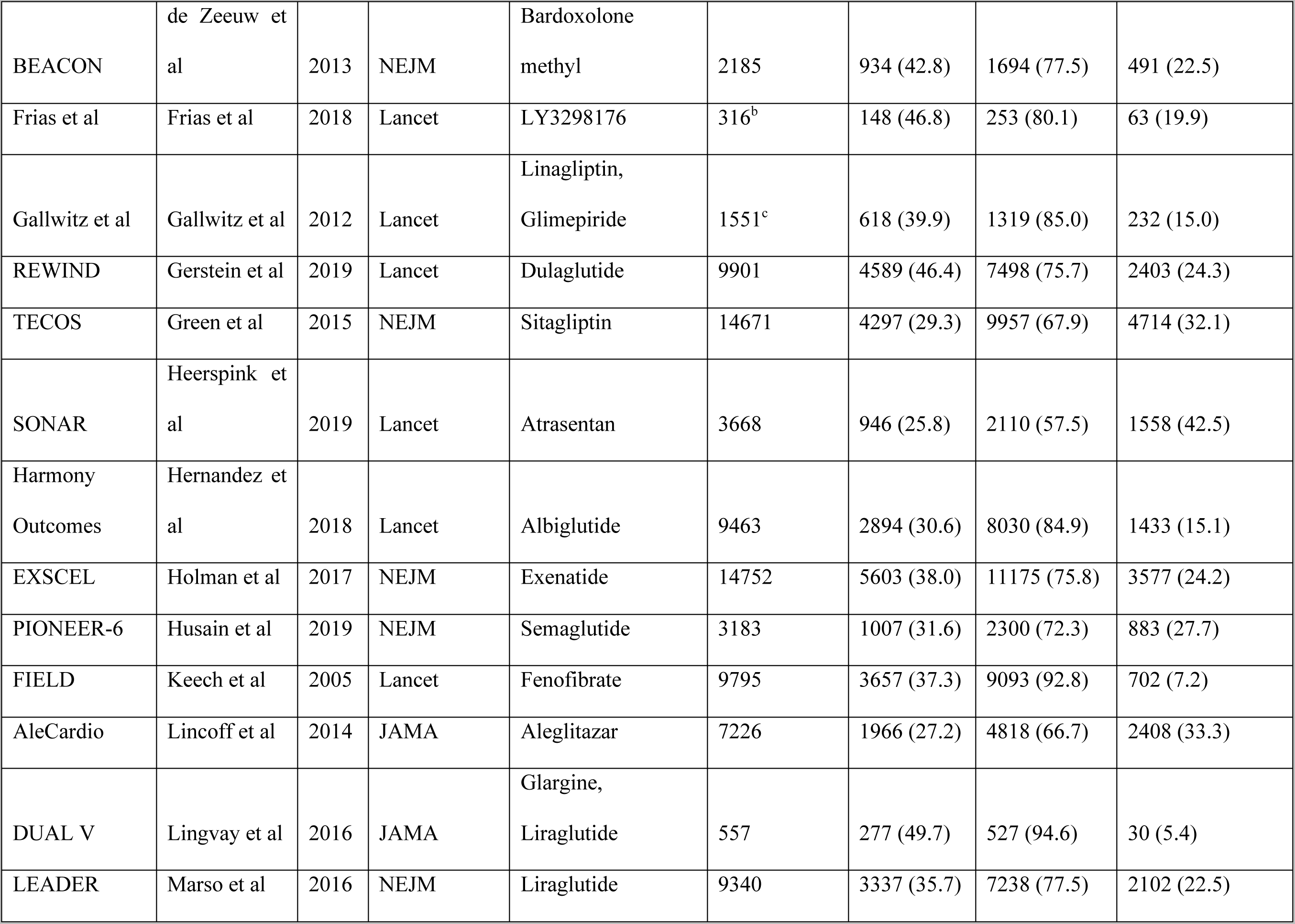
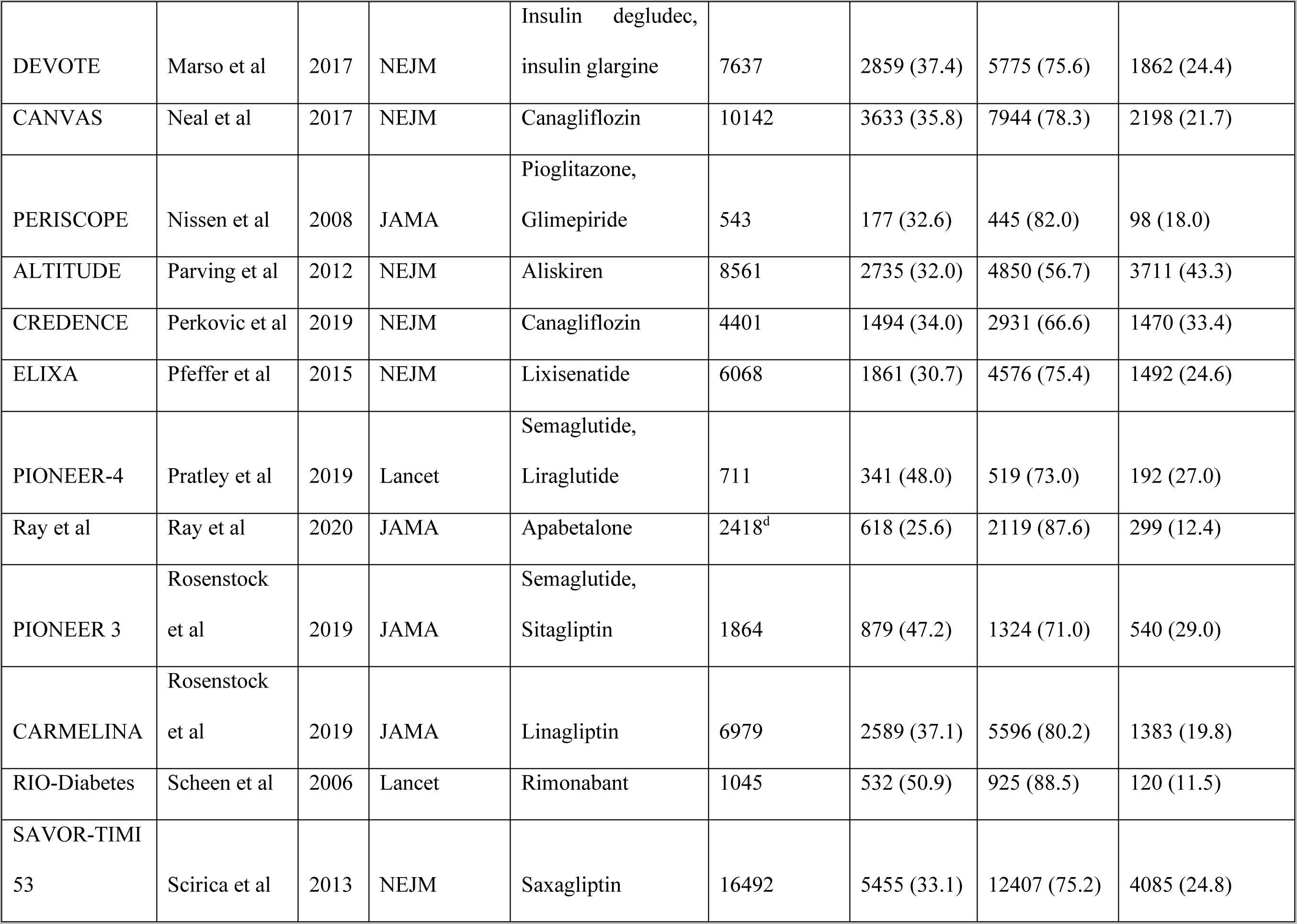
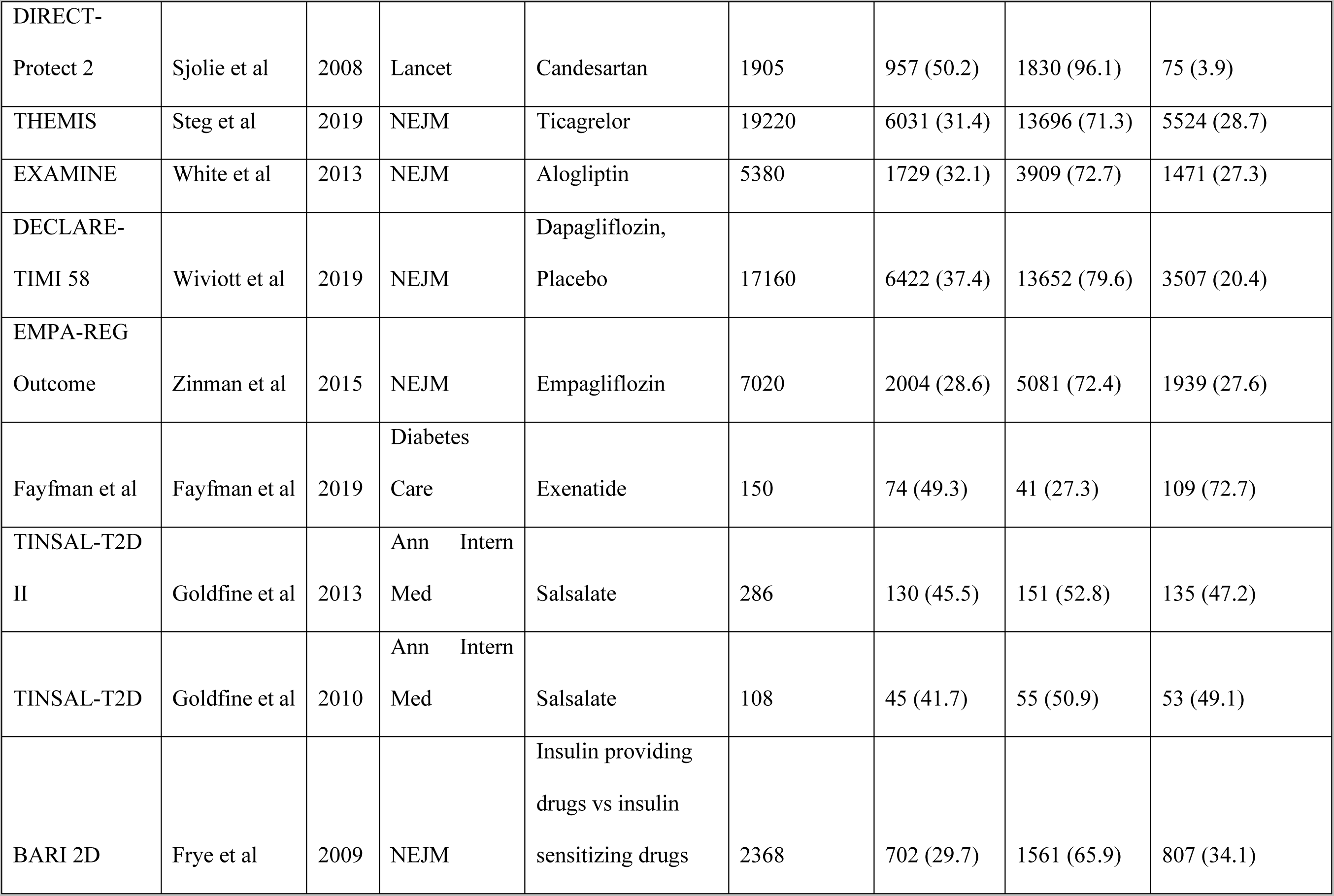
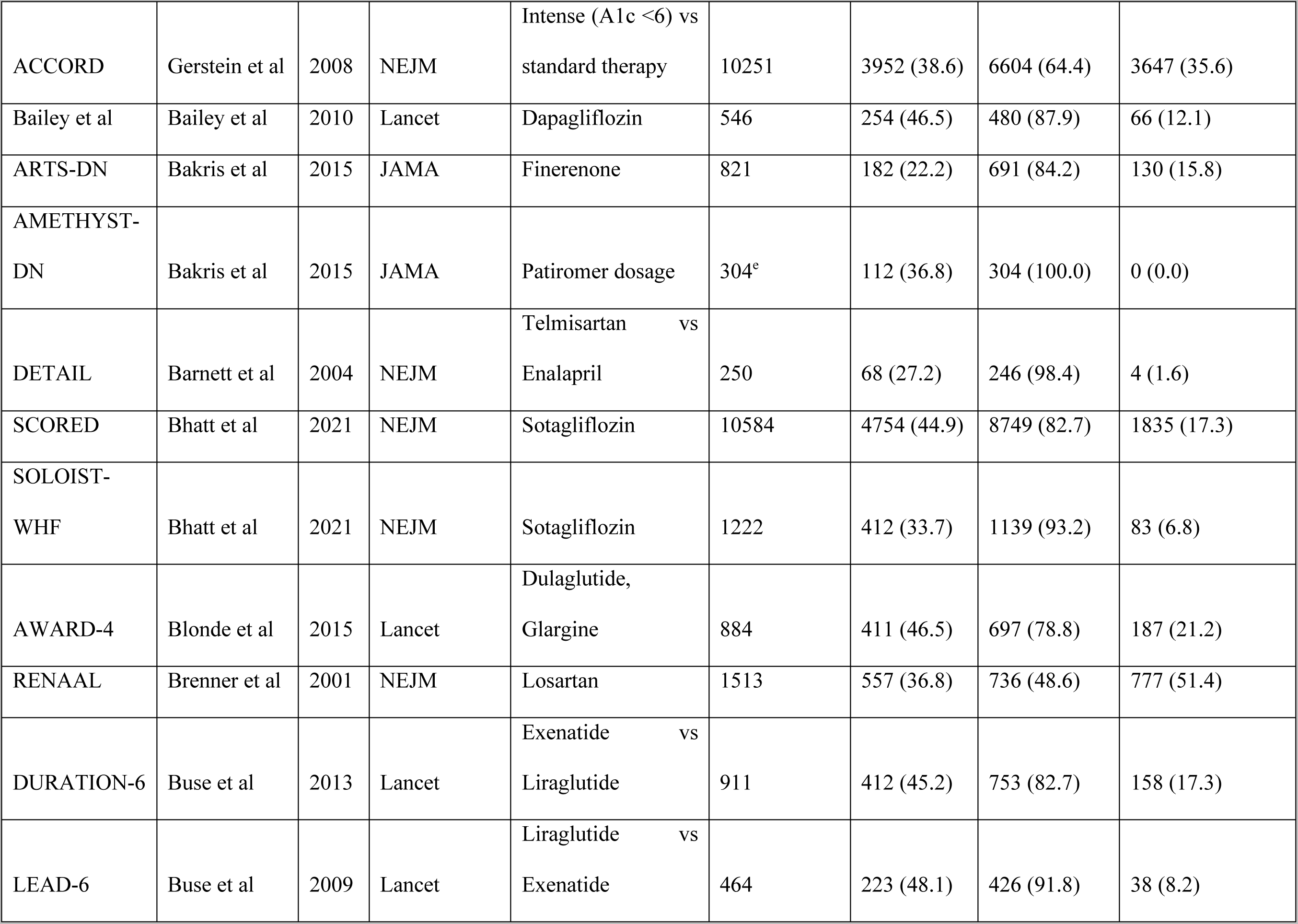
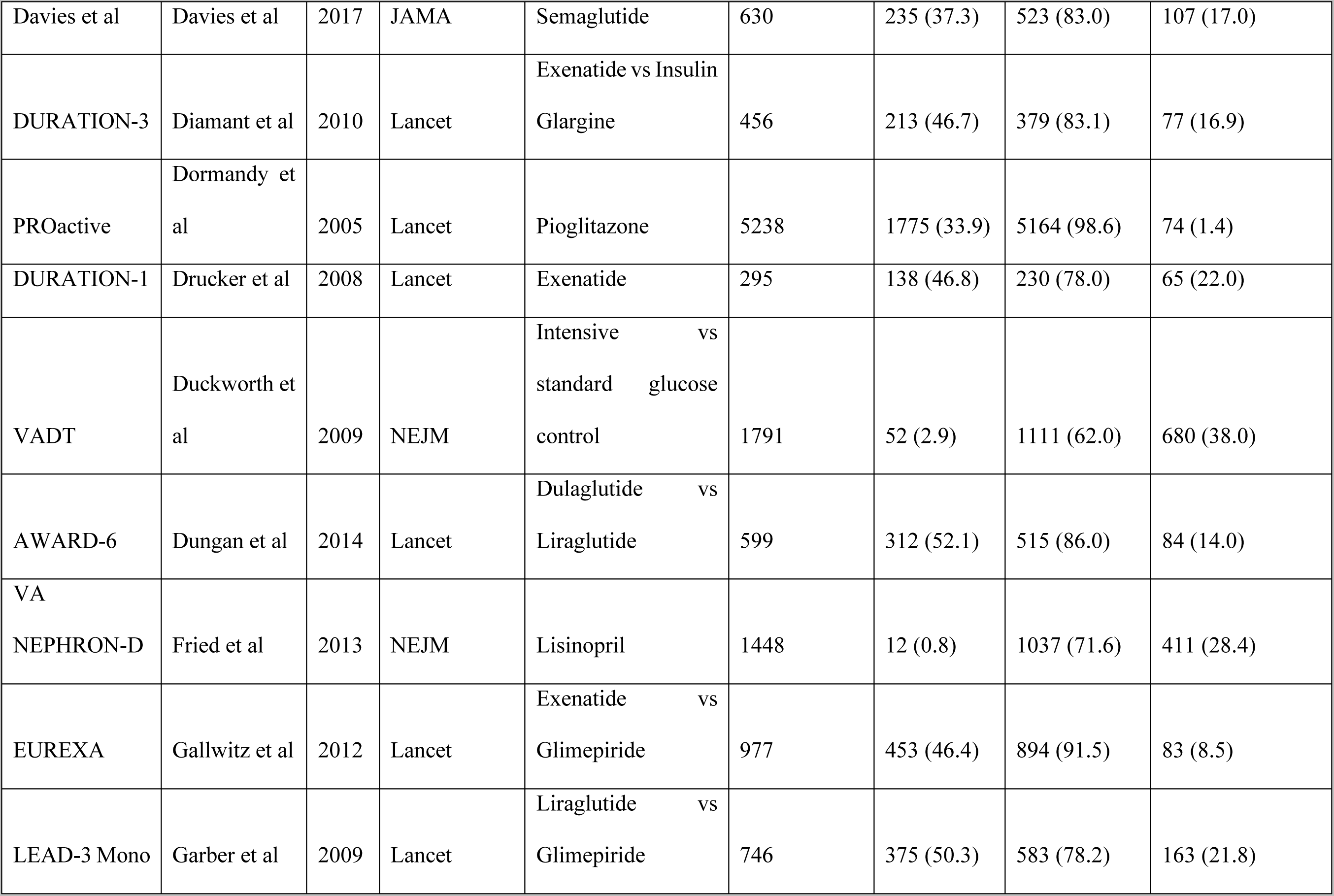
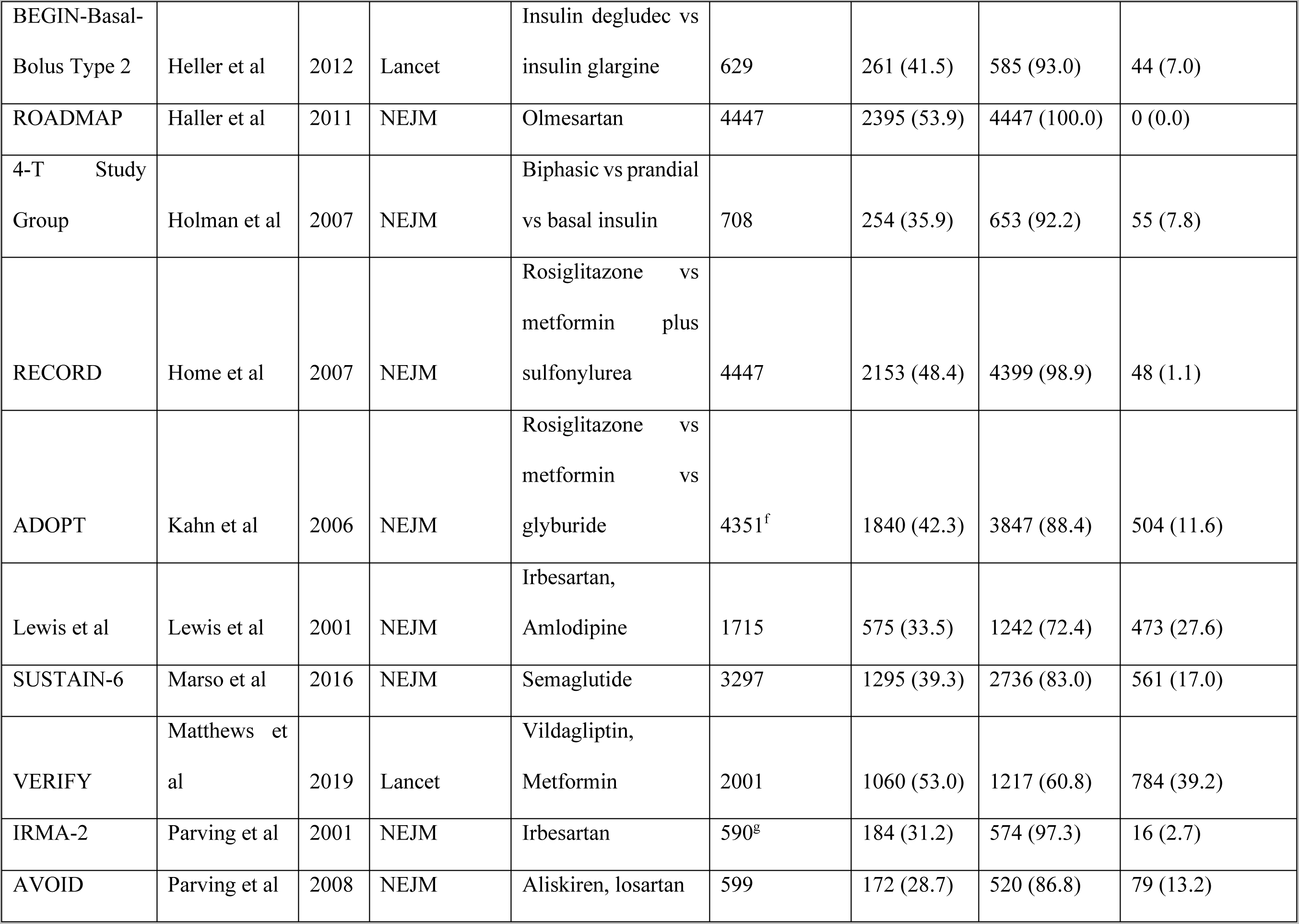
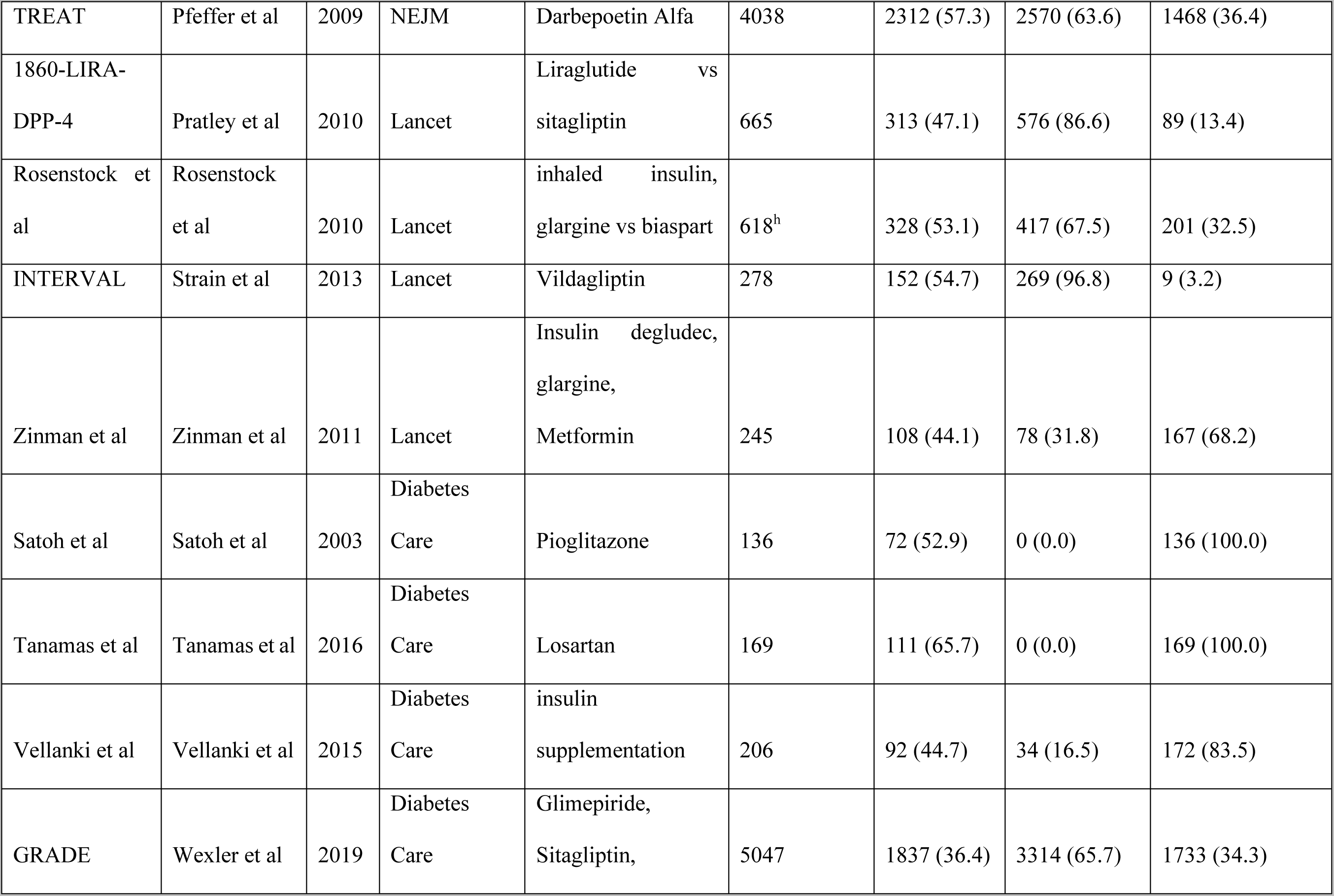
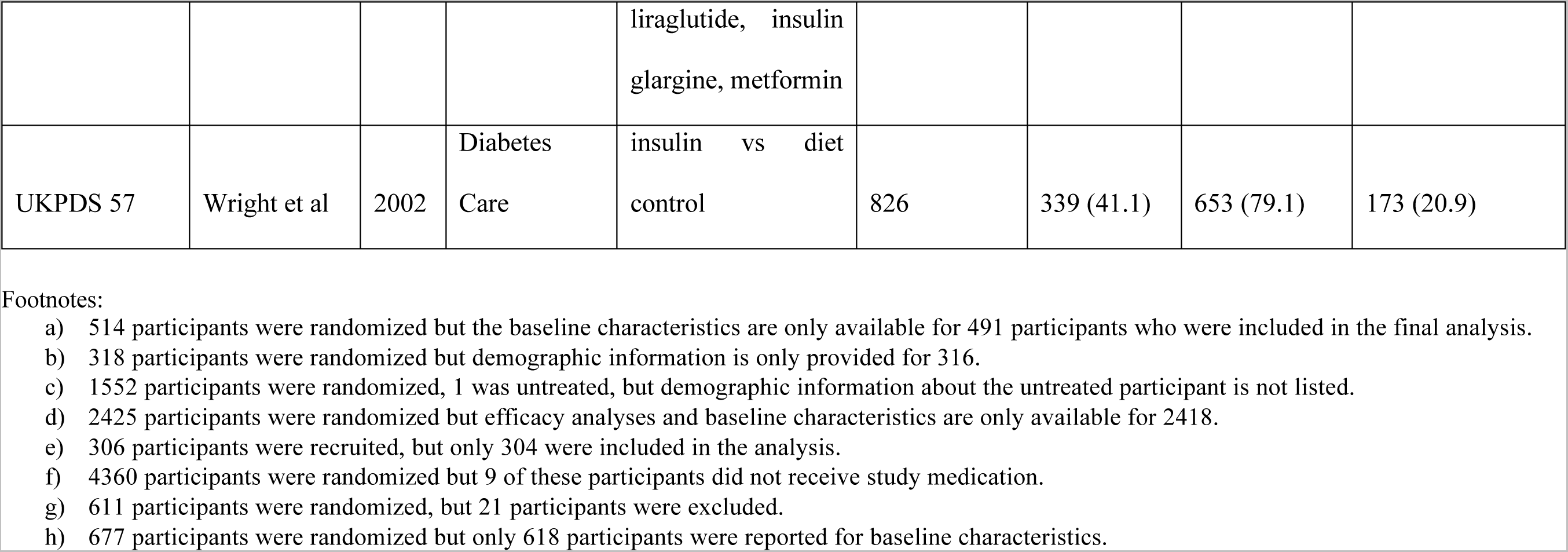
List of Randomized Clinical Trials Included in Systematic Review.

The pooled PPR for the white group is 2.19 (95%CI: 1.91, 2.50) for industry trials, consistent with overrepresentation, and 1.11 (95%CI: 1.00, 1.23) in government trials, consistent with proportional representation. The PPR for non-white ethnic group(s) is 0.33 (95%CI: 0.29, 0.38) for industry trials, and 0.73 (95%CI: 0.62, 0.86) for government trials, both consistent with underrepresentation (Figure 1).

**Figure 1.**
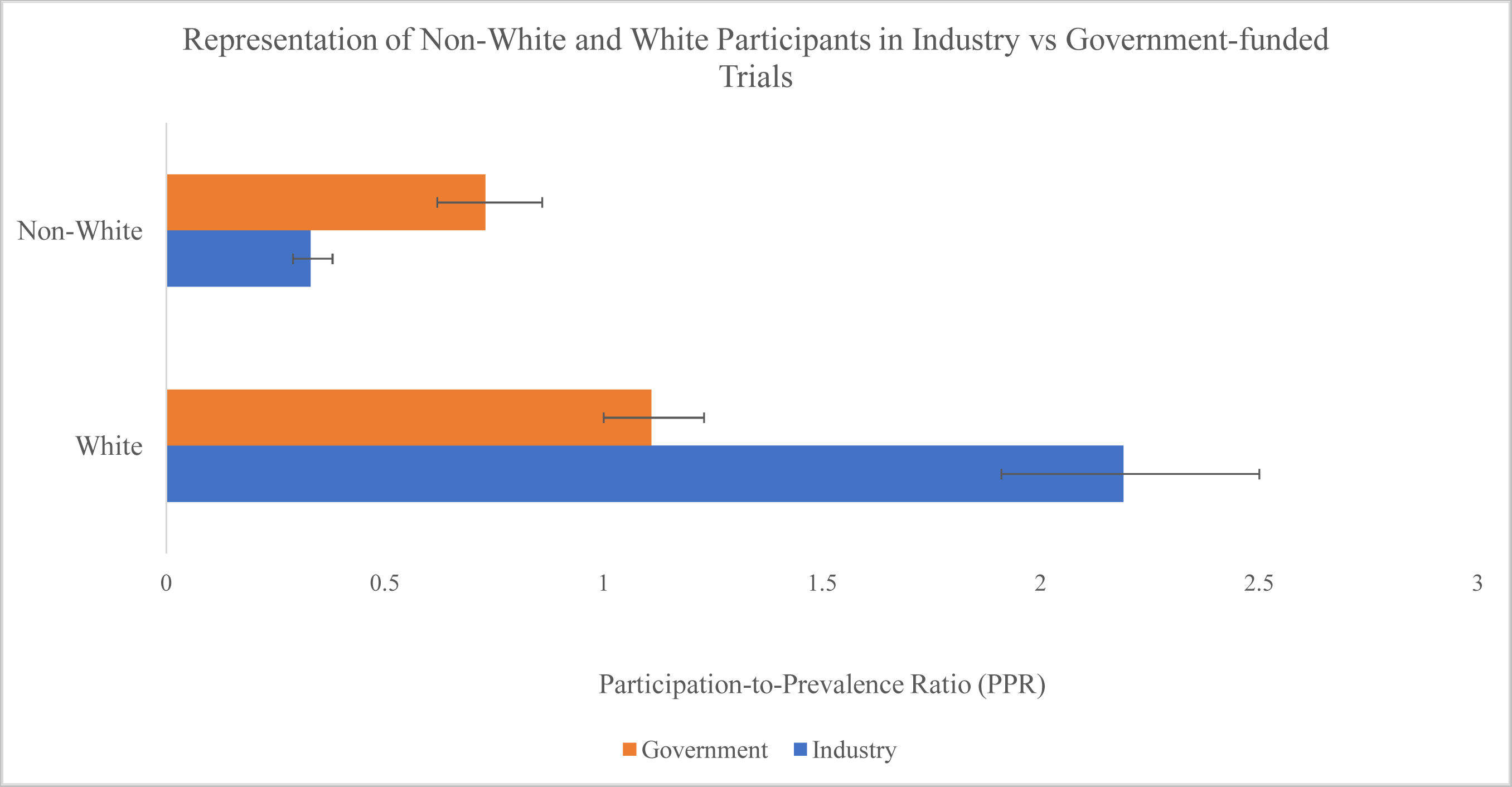
Representation of Non-White and White Participants in Industry vs. Government Funded Trials. Figure legend: Bars represent 95% confidence intervals.

### Sensitivity Analysis

Nineteen industry-funded trials in this review did not provide country or region-specific data indicating from where participants were recruited.^10–13, 19, 22, 27, 39, 54–57, 63, 68–71, 75, 77^ For these trials, the worldwide proportion of white and non-white people was estimated by assuming that all people in Europe, Canada, the United States, and Australia were white; this resulted in an estimate of 14.8% white people and 85.2% non-white people. A series of sensitivity analyses that varied the estimated proportion of white and non-white people for these seven trials in which we imputed these values was conducted. The proportion of white participants was varied from 80% through 90%; and of non-white participants, from 10% through 20%. These did not result in appreciably different estimates from the main analyses. The full data for these sensitivity analyses are shown in Appendix B.

## Discussion

T2DM disproportionately affects non-white populations worldwide.^92^ The objective of this meta-analysis was to document the underrepresentation of non-white ethnic participants in large RCTs of T2DM therapies, and to investigate the differences in ethnic representation between industry-funded trials and government-funded trials. This analysis shows that white individuals are over-enrolled, and non-white individuals are under-enrolled in diabetes RCTs. Government-funded T2DM trials tend to have better representation of non-white participants than do industry-funded trials.

The finding that government-funded trials recruit more diverse participants may reflect adherence to the more stringent funding conditions associated with accepting funding from government bodies. For example, the NIH emphasizes the inclusion and appropriate representation of minority groups in clinical research.^4^ This measure attempts to ensure that visible minority populations are more proportionately represented in NIH-funded clinical research, and that research findings are generalizable across all ethnic groups. Industry-funded trials do not have the same requirements as the NIH, and therefore, control over recruitment is held entirely by the trial Steering Committee, sponsor, and research staff.

Issues with diverse representation extend beyond RCTs of T2DM. Underrepresentation of non-white populations was apparent in 2020 COVID-19 trials as well.^93^ Non-white populations, specifically Black, Native Americans, and LatinX people were disproportionately impacted by COVID-19, however, they were inadequately represented in remdesivir clinical trials.^93^ The underrepresentation of non-white participants limits the ability to generalize results and safety outcomes for all ethnic groups and may limit the update of this evidence by the non-white ethnic communities, which adds to the disadvantages they face.

For industry trials, white participants tend to be overrepresented relative to their disease burden (Figure 2a). However, this estimate was heterogenous. For example, some studies had PPR values that were significantly higher than the others (i.e., > 1.78). ^10–13, 19, 22, 27, 39, 54–57, 63, 68–71, 75, 77^ This is because these RCTs did not provide country or region-specific data regarding from where participants were recruited. As such, worldwide estimates were determined for both prevalence and demographic data. Prevalence estimates were obtained from Saeedi et al^94^ and are listed in eTable 4 in the Supplement. The worldwide white and non-white population proportion estimates do not necessarily reflect the demographics of where participants were recruited for these studies. Therefore, the PPRs for white participants in industry trials are likely overestimates, however, our results were not seriously sensitive to altering this assumption (Appendix B). This is because the proportion of non-white participants in these trials was relatively small, and therefore the varying worldwide proportion estimates did not greatly affect the pooled PPR. Greater variance was observed among the different proportions of white participants in the trials. Overall, the effects of the varying worldwide proportions of both white and non-white people on the PPRs were limited. This is because estimates were only altered for 19 out of the 68 industry trials and the participants in all 19 trials only comprise 7.6% of the total participants across the industry-funded trials in this review.

**Figure 2a.**
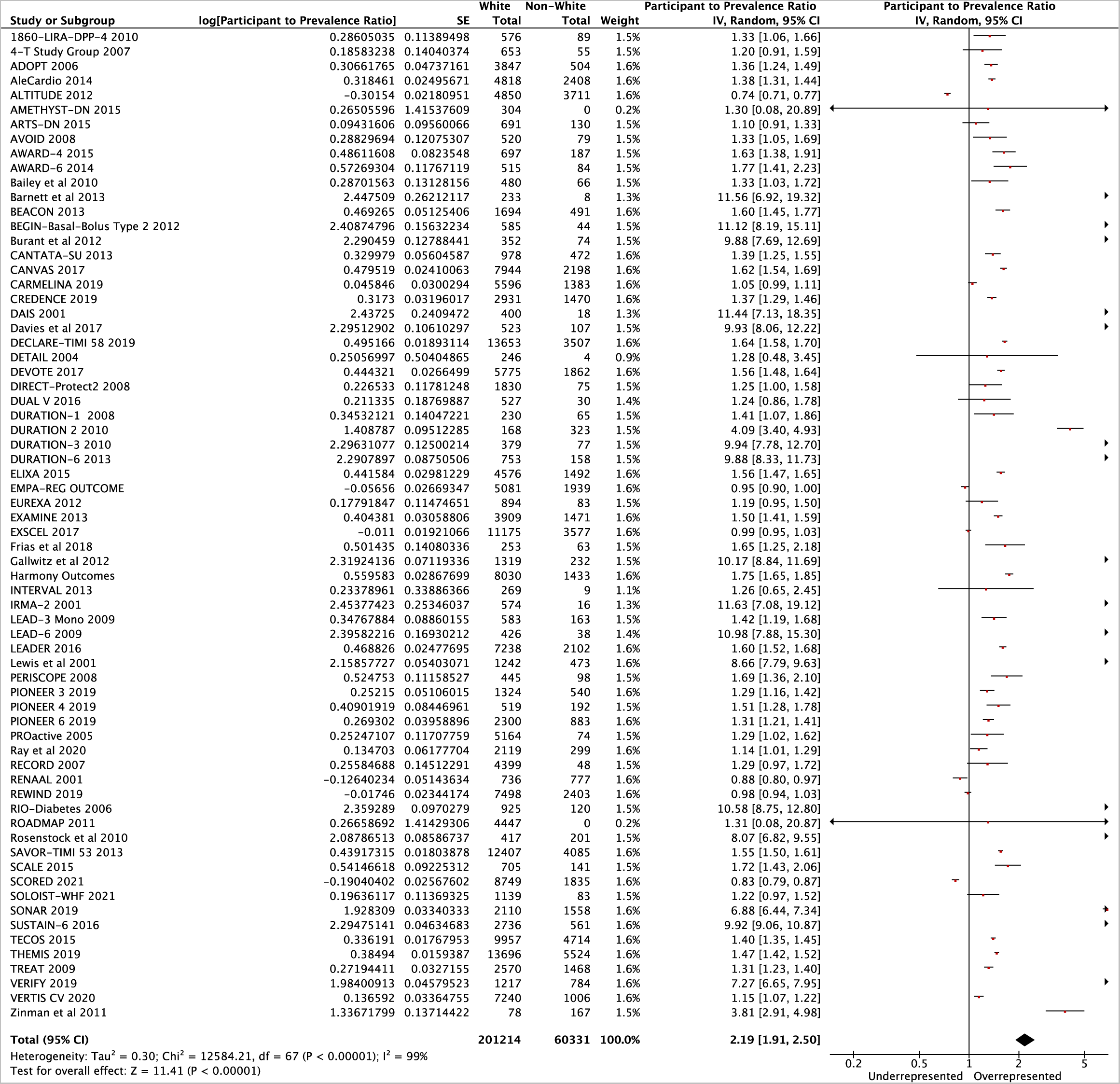
Forest plot of comparison: PPR White People – Industry Trials. Figure Legend: Individual PPRs of 68 industry-funded trials are shown along with a pooled PPR for the white population. Lines represent 95% confidence intervals.

Non-white participants tended to be underrepresented in industry-funded trials relative to their disease burden (Figure 4). Trials such as EXSCEL^24^, REWIND^20^, RENAAL^53^, EMPA-REG OUTCOME^45^, and ALTITUDE^32^ represent outliers to this trend with non-white PPRs of 1.04, 1.06, 1.15, 1.18, and 1.85, respectively. These trials all recruited over 1500 participants from at least 24 countries including regions of North America, Europe, South America, Asia, and Africa. Future industry-funded trials should consider enrolling participants from diverse communities and regions of the world especially when the disease burden is higher than in white European origin individuals and regions where white Europeans are the majority.

The potential explanations for the patterns of overrepresentation of white participants and underrepresentation of non-white participants are as follows: first, inclusion and exclusion criteria of RCTs may favour enrolling white participants over non-white participants. For example, specialty clinics and hospitals where research is taking place may be located in areas with lower proportions of non-white patients, affecting the overall diversity of participants in RCTs. Second, limited screening of non-white individuals for enrollment may occur because of possible implicit biases, and social or medical reasons that make their participation difficult; third, distrust and fear of medical institutions due to historical mistreatment of minority groups may result in a lack of willingness of non-white people to participate in clinical trials; fourth, minority groups may not enrol because of language barriers, cultural practices or related contextual factors that limit their participation, including socioeconomic disadvantage; fifth, logistical barriers may exist such as inflexible work schedules and additional costs associated with participating in research studies such as costs of public transportation and/or fuel.^93^

Furthermore, the lack of diversity among Principal Investigators, local investigators, and study staff of some RCTs may also be related to the lower enrolment of non-white participants.^93^ For example, in an NIH study on the diversity of the NIH-funded workforce, it was found that 71.9% of principal investigators on NIH-funded research project grants were white.^95^ Diverse representation among the study staff might increase the confidence of participants from minority communities and improve communication.^93^ Additionally, industry-funded trials could potentially benefit from having ethnic participant recruitment guidelines similar to the ones that exist for the NIH. Regulatory bodies could indicate that proportional representation of participants affected by the disease of interest by ethnicity is strongly recommended or mandatory such that industry trial leaders will more carefully consider who and from where to recruit within a given country or region. Future reviews of ethnic participant recruitment should be completed for trials that were conducted within a single country to assess country-specific trends. These reviews could guide clinical practice and be used to establish recruitment standards that are reasonable given the prevalence of T2DM, and demographics in a given country. Future studies should also be conducted to analyze ethnic participant recruitment trends in RCTs for other health conditions which are known to be common in non-white populations such as hypertension and stroke.

Our study has several strengths. The meta-analysis included trials that were published in journals with high impact factors (>/=30 for industry trials, >/=10 for government), which are likely to be highly cited and used to inform clinical guidelines. Analyzing ethnic participant recruitment in these studies allows for a finer assessment of the generalizability of the results of these studies to certain practice settings. The calculation and presentation of the PPR metric improves the summary of the findings of overrepresentation and underrepresentation of white and non-white participants (Figure 2a; Figure 2b; Figure 3a; Figure 3b).

**Figure 2b.**
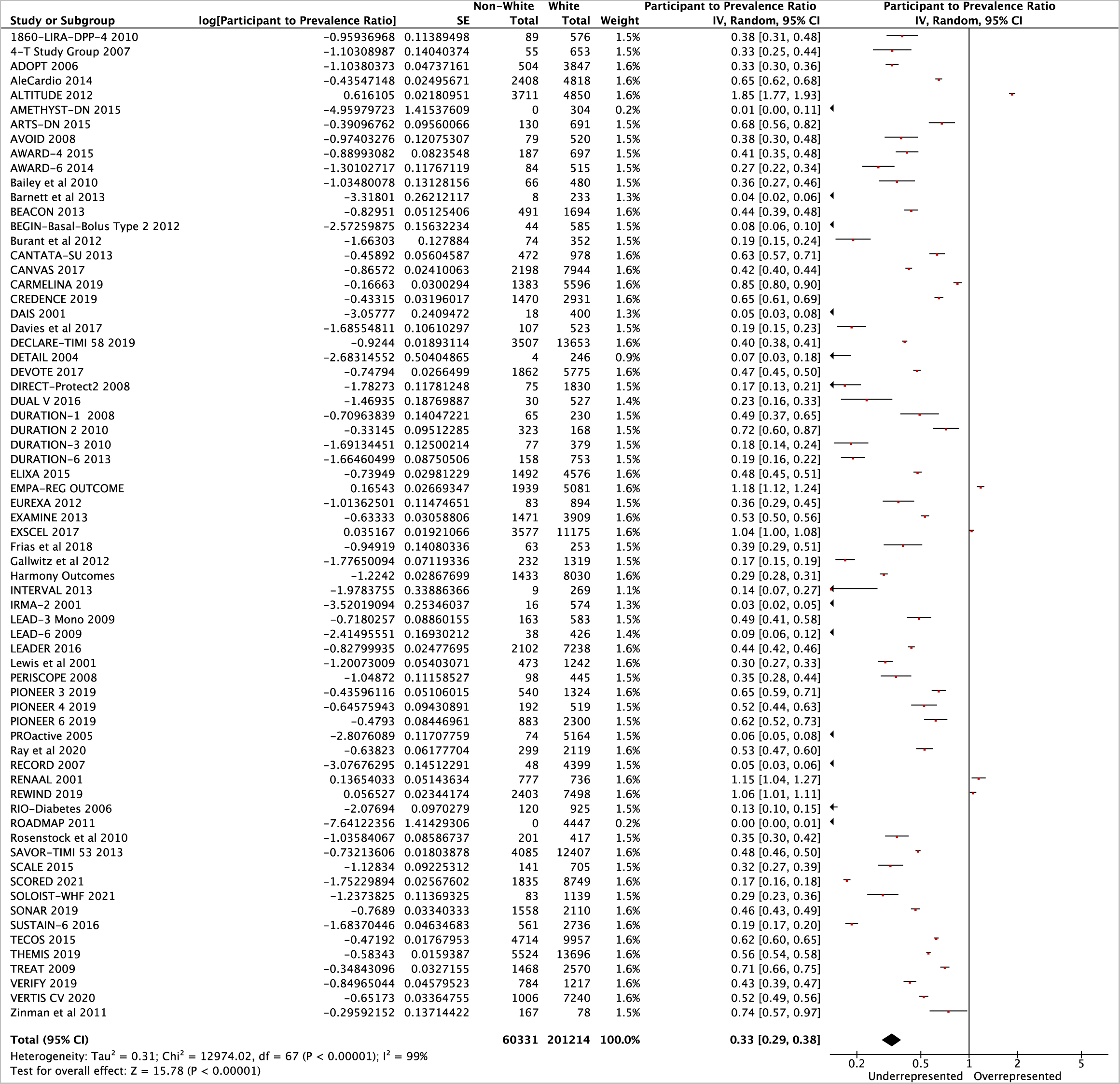
Forest plot of comparison: PPR Non-White People – Industry Trials. Figure Legend: Individual PPRs of 68 industry-funded trials are shown along with a pooled PPR for the non-white population. Lines represent 95% confidence intervals.

**Figure 3a.**
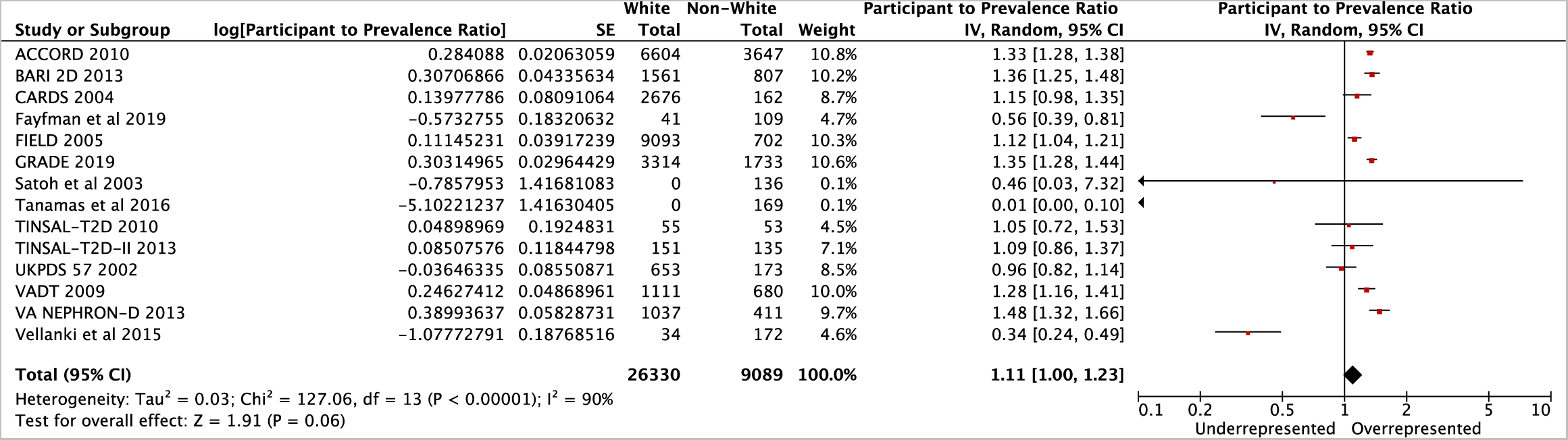
Forest plot of comparison: PPR White People – Government Trials. Figure Legend: Individual PPRs of 14 government-funded trials are shown along with a pooled PPR for the white population. Lines represent 95% confidence intervals.

**Figure 3b.**
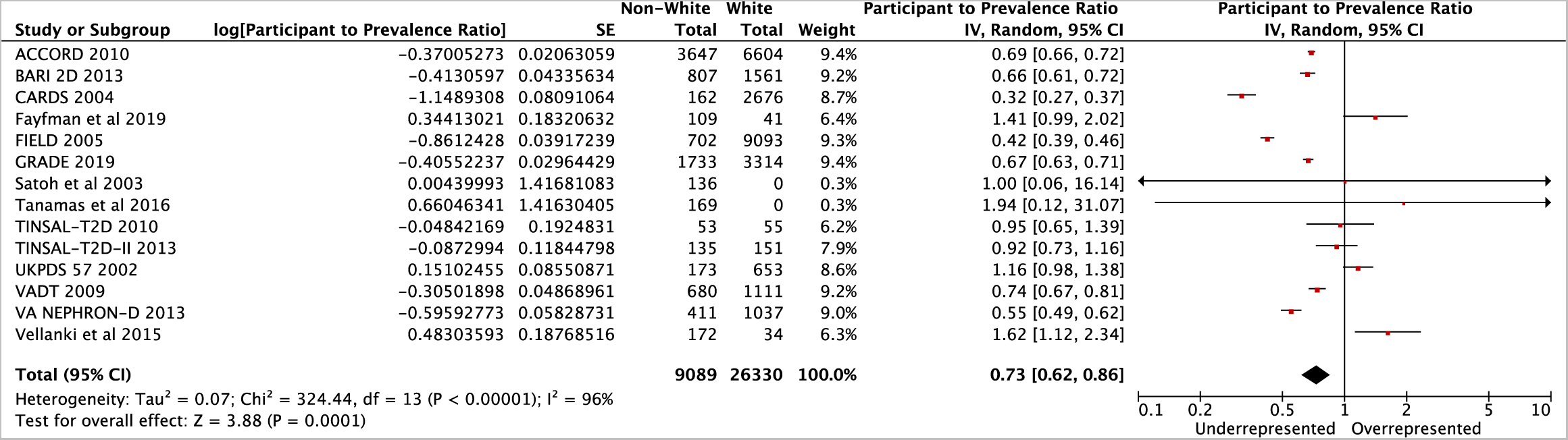
Forest plot of comparison: PPR Non-White People – Government Trials. Figure Legend: Individual PPRs of 14 government-funded trials are shown along with a pooled PPR for the non-white population. Lines represent 95% confidence intervals.

Our study has certain limitations. First, the PPR denominator calculations were based on prevalence and demographic data for white and non-white participants in different countries and regions of the world. While T2DM prevalence and demographic data are typically documented for specific countries, limited data exist for T2DM prevalence and demographics in larger regions of the world. When specific data were not available, these values were estimated using a standardized algorithm. Second, this meta-analysis only included studies that were published on January 1, 2000, and onwards. However, our estimates of the prevalence and demographic data do not correspond to the prevalence of T2DM in white and non-white people and the ethnic breakdown of countries or regions at the time that each trial was conducted, although we attempted to match them as closely as possible. Finally, our pooled meta-analytic estimates had high heterogeneity (I-squared >90%). This heterogeneity may stem from many factors, including study design, country or region of conduct, study size, or dates of enrolment. However, despite this, our results were consistent with respect to direction. Only 6 of 68 industry trials had PPR <1 for white people and PPR > 1 for non-white people, and these carried 9.6% of the weight in the pooled estimates, and only 5 of 14 government trials had PPR <1 for white people, and >1 for non-white people, and these carried 18.0% of the weight in the pooled estimates. Thus, statistical heterogeneity influenced the precision of our estimates, but not the direction or magnitude to an extent that would cause concern. Finally, we did not conduct risk of bias assessments for the trials used in this review. Our selection criteria were such that we only included large (n>100) randomized trials (lowest risk of bias design) published in high-impact journals, which helps ensure comparable, and low risk of bias across included studies.

## Conclusion

This review examined the representation of white and non-white populations in T2DM RCTs. Non-white participants appear to be underrepresented in both industry and government-funded T2DM trials relative to their disease burden, while white participants appear to be overrepresented. The systematic review shows that the greatest disparity in diversity between the groups in RCTs occurs in industry-funded trials. Strategies to improve recruitment and enrolment of diverse populations into international industry and government-funded RCTs should be explored.

## Supporting information

PRISMA

eTable 1

eTable 2

eTable 3

eTable 4

eFigure 1

## Data Availability

All data produced in the present work are contained in the manuscript.

## Acknowledgements

Dr. Anand holds a Canada Research Chair in Ethnic Diversity and Cardiovascular Disease (Tier 1) and Michael G DeGroote Heart and Stroke Chair in Population Health.

We thank Justin Wu (McMaster University) for his assistance with extracting data and creating summary tables.

## Disclosures

Dr. de Souza has served as an external resource person to the World Health Organization’s Nutrition Guidelines Advisory Group on trans fats, saturated fats, and polyunsaturated fats. The WHO paid for his travel and accommodation to attend meetings from 2012-2017 to present and discuss this work. He has also done contract research for the Canadian Institutes of Health Research’s Institute of Nutrition, Metabolism, and Diabetes, Health Canada, and the World Health Organization for which he received remuneration. He has received speaker’s fees from the University of Toronto, and McMaster Children’s Hospital. He has held grants from the Canadian Institutes of Health Research, Canadian Foundation for Dietetic Re-search, Population Health Research Institute, and Hamilton Health Sciences Corporation as a principal investigator, and is a co-investigator on several funded team grants from the Canadian Institutes of Health Research. He serves as a member of the Nutrition Science Advisory Committee to Health Canada (Government of Canada), and a co-opted member of the Scientific Advisory Committee on Nutrition (SACN) Subgroup on the Framework for the Evaluation of Evidence, and as an independent director of the Helderleigh Foundation (Canada). All other authors declare no conflicts of interest.

## Registration and Protocol

This study has been registered into an embargo on Open Science Framework (OSF) under the name: Assessing participation of non-white people in type 2 diabetes mellitus randomized clinical trials: A Meta-Analysis. A protocol was not prepared.

## Notes

### Funding Statement

This study did not receive any funding.

