## Supplementary material for "Assessing non-white ethnic participation in type 2 diabetes mellitus randomized clinical trials: A Meta-Analysis": eTable 1

**eTable 1. PubMed Search Strategy**

| N | Search Terms | Records |
| --- | --- | --- |
| 1 | ("The New England journal of medicine"[Journal] AND ("diabete"[All Fields] OR "diabetes mellitus"[MeSH Terms] OR ("diabetes"[All Fields] AND "mellitus"[All Fields]) OR "diabetes mellitus"[All Fields] OR "diabetes"[All Fields] OR "diabetes insipidus"[MeSH Terms] OR ("diabetes"[All Fields] AND "insipidus"[All Fields]) OR "diabetes insipidus"[All Fields] OR "diabetic"[All Fields] OR "diabetics"[All Fields] OR "diabets"[All Fields])) AND (2000/1/1:2021/1/1[pdat]) | 1331 |
| 2 | ("The New England journal of medicine"[Journal] AND ("diabete"[All Fields] OR "diabetes mellitus"[MeSH Terms] OR ("diabetes"[All Fields] AND "mellitus"[All Fields]) OR "diabetes mellitus"[All Fields] OR "diabetes"[All Fields] OR "diabetes insipidus"[MeSH Terms] OR ("diabetes"[All Fields] AND "insipidus"[All Fields]) OR "diabetes insipidus"[All Fields] OR "diabetic"[All Fields] OR "diabetics"[All Fields] OR "diabets"[All Fields]) AND 2000/01/01:2021/01/01[Date - Publication]) AND ((clinicaltrial[Filter] OR randomizedcontrolledtrial[Filter]) AND (2000/1/1:2021/1/1[pdat])) | 218 |
| 3 | ("lancet london england"[Journal] AND ("diabete"[All Fields] OR "diabetes mellitus"[MeSH Terms] OR ("diabetes"[All Fields] AND "mellitus"[All Fields]) OR "diabetes mellitus"[All Fields] OR "diabetes"[All Fields] OR "diabetes insipidus"[MeSH Terms] OR ("diabetes"[All Fields] AND "insipidus"[All Fields]) OR "diabetes insipidus"[All Fields] OR "diabetic"[All Fields] OR "diabetics"[All Fields] OR "diabets"[All Fields])) AND (2000/1/1:2021/1/1[pdat]) | 1295 |
| 4 | ("lancet london england"[Journal] AND ("diabete"[All Fields] OR "diabetes mellitus"[MeSH Terms] OR ("diabetes"[All Fields] AND "mellitus"[All Fields]) OR "diabetes mellitus"[All Fields] OR "diabetes"[All Fields] OR "diabetes insipidus"[MeSH Terms] OR ("diabetes"[All Fields] AND "insipidus"[All Fields]) OR "diabetes insipidus"[All Fields] OR "diabetic"[All Fields] OR "diabetics"[All Fields] OR "diabets"[All Fields])) AND ((clinicaltrial[Filter] OR randomizedcontrolledtrial[Filter]) AND (2000/1/1:2021/1/1[pdat])) | 180 |
| 5 | ("JAMA"[Journal] AND ("diabete"[All Fields] OR "diabetes mellitus"[MeSH Terms] OR ("diabetes"[All Fields] AND "mellitus"[All Fields]) OR "diabetes mellitus"[All Fields] OR "diabetes"[All Fields] OR "diabetes insipidus"[MeSH Terms] OR ("diabetes"[All Fields] AND "insipidus"[All Fields]) OR "diabetes insipidus"[All Fields] OR "diabetic"[All Fields] OR "diabetics"[All Fields] OR "diabets"[All Fields])) AND (2000/1/1:2021/1/1[pdat]) | 1193 |
| 6 | ("JAMA"[Journal] AND ("diabete"[All Fields] OR "diabetes mellitus"[MeSH Terms] OR ("diabetes"[All Fields] AND "mellitus"[All Fields]) OR "diabetes mellitus"[All Fields] OR "diabetes"[All Fields] OR "diabetes insipidus"[MeSH Terms] OR ("diabetes"[All Fields] AND "insipidus"[All Fields]) OR "diabetes insipidus"[All Fields] OR "diabetic"[All Fields] OR "diabetics"[All Fields] OR "diabets"[All Fields])) AND ((clinicaltrial[Filter] OR randomizedcontrolledtrial[Filter]) AND (2000/1/1:2021/1/1[pdat])) | 143 |
| 7 | (("British medical journal"[Journal] OR "bmj clinical research ed"[Journal]) AND ("diabete"[All Fields] OR "diabetes mellitus"[MeSH Terms] OR ("diabetes"[All Fields] AND "mellitus"[All Fields]) OR "diabetes mellitus"[All Fields] OR "diabetes"[All Fields] OR "diabetes insipidus"[MeSH Terms] OR ("diabetes"[All Fields] AND "insipidus"[All Fields]) OR "diabetes insipidus"[All Fields] OR "diabetic"[All Fields] OR "diabetics"[All Fields] OR "diabets"[All Fields])) AND (2000/1/1:2021/1/1[pdat]) | 1394 |
| 8 | (("British medical journal"[Journal] OR "bmj clinical research ed"[Journal]) AND ("diabete"[All Fields] OR "diabetes mellitus"[MeSH Terms] OR ("diabetes"[All Fields] AND "mellitus"[All Fields]) OR "diabetes mellitus"[All Fields] OR "diabetes"[All Fields] OR "diabetes insipidus"[MeSH Terms] OR ("diabetes"[All Fields] AND "insipidus"[All Fields]) OR "diabetes insipidus"[All Fields] OR "diabetic"[All Fields] OR "diabetics"[All Fields] OR "diabets"[All Fields])) AND ((clinicaltrial[Filter] OR randomizedcontrolledtrial[Filter]) AND (2000/1/1:2021/1/1[pdat])) | 64 |
| 9 | ("Circulation"[Journal] AND ("diabete"[All Fields] OR "diabetes mellitus"[MeSH Terms] OR ("diabetes"[All Fields] AND "mellitus"[All Fields]) OR "diabetes mellitus"[All Fields] OR "diabetes"[All Fields] OR "diabetes insipidus"[MeSH Terms] OR ("diabetes"[All Fields] AND "insipidus"[All Fields]) OR "diabetes insipidus"[All Fields] OR "diabetic"[All Fields] OR "diabetics"[All Fields] OR "diabets"[All Fields])) AND (2000/1/1:2021/1/1[pdat]) | 1703 |
| 10 | ("Circulation"[Journal] AND ("diabete"[All Fields] OR "diabetes mellitus"[MeSH Terms] OR ("diabetes"[All Fields] AND "mellitus"[All Fields]) OR "diabetes mellitus"[All Fields] OR "diabetes"[All Fields] OR "diabetes insipidus"[MeSH Terms] OR ("diabetes"[All Fields] AND "insipidus"[All Fields]) OR "diabetes insipidus"[All Fields] OR "diabetic"[All Fields] OR "diabetics"[All Fields] OR "diabets"[All Fields])) AND ((clinicaltrial[Filter] OR randomizedcontrolledtrial[Filter]) AND (2000/1/1:2021/1/1[pdat])) | 305 |
| 11 | ("Annals of internal medicine"[Journal] AND ("diabete"[All Fields] OR "diabetes mellitus"[MeSH Terms] OR ("diabetes"[All Fields] AND "mellitus"[All Fields]) OR "diabetes mellitus"[All Fields] OR "diabetes"[All Fields] OR "diabetes insipidus"[MeSH Terms] OR ("diabetes"[All Fields] AND "insipidus"[All Fields]) OR "diabetes insipidus"[All Fields] OR "diabetic"[All Fields] OR "diabetics"[All Fields] OR "diabets"[All Fields])) AND (2000/1/1:2021/1/1[pdat]) | 939 |
| 12 | ("Annals of internal medicine"[Journal] AND ("diabete"[All Fields] OR "diabetes mellitus"[MeSH Terms] OR ("diabetes"[All Fields] AND "mellitus"[All Fields]) OR "diabetes mellitus"[All Fields] OR "diabetes"[All Fields] OR "diabetes insipidus"[MeSH Terms] OR ("diabetes"[All Fields] AND "insipidus"[All Fields]) OR "diabetes insipidus"[All Fields] OR "diabetic"[All Fields] OR "diabetics"[All Fields] OR "diabets"[All Fields])) AND ((clinicaltrial[Filter] OR randomizedcontrolledtrial[Filter]) AND (2000/1/1:2021/1/1[pdat])) | 68 |
| 13 | ("Diabetes care"[Journal] AND ("diabete"[All Fields] OR "diabetes mellitus"[MeSH Terms] OR ("diabetes"[All Fields] AND "mellitus"[All Fields]) OR "diabetes mellitus"[All Fields] OR "diabetes"[All Fields] OR "diabetes insipidus"[MeSH Terms] OR ("diabetes"[All Fields] AND "insipidus"[All Fields]) OR "diabetes insipidus"[All Fields] OR "diabetic"[All Fields] OR "diabetics"[All Fields] OR "diabets"[All Fields])) AND (2000/1/1:2021/1/1[pdat]) | 12294 |
| 14 | ("Diabetes care"[Journal] AND ("diabete"[All Fields] OR "diabetes mellitus"[MeSH Terms] OR ("diabetes"[All Fields] AND "mellitus"[All Fields]) OR "diabetes mellitus"[All Fields] OR "diabetes"[All Fields] OR "diabetes insipidus"[MeSH Terms] OR ("diabetes"[All Fields] AND "insipidus"[All Fields]) OR "diabetes insipidus"[All Fields] OR "diabetic"[All Fields] OR "diabetics"[All Fields] OR "diabets"[All Fields])) AND ((clinicaltrial[Filter] OR randomizedcontrolledtrial[Filter]) AND (2000/1/1:2021/1/1[pdat])) | 2082 |
| 15 | 1 and 3 and 5 and 7 and 9 and 11 and 13 (“Journal, time frame and topic filter”) | 20149 |
| **16** | **2 and 4 and 6 and 8 and 10 and 12 and 14 (+ “Randomized trial filter”)** | **3060** |
