## Supplementary material for "Assessing non-white ethnic participation in type 2 diabetes mellitus randomized clinical trials: A Meta-Analysis": eTable 2

**eTable 2. Study Selection Criteria**

| Criterion | Industry | Government |
| --- | --- | --- |
| Described the results of an RCT of a T2DM pharmacotherapy | 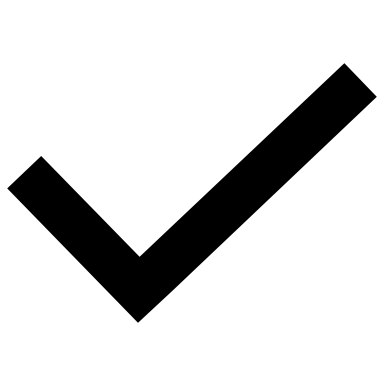 | 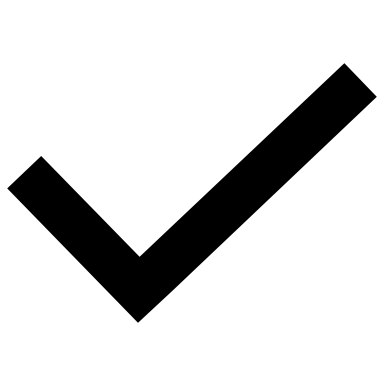 |
| Published in a journal with an impact factor (IF)>30 | 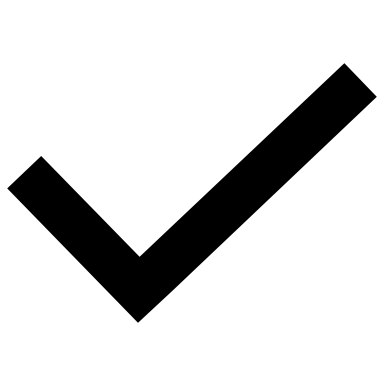 |  |
| Published in a journal with an IF>10^a^ |  | 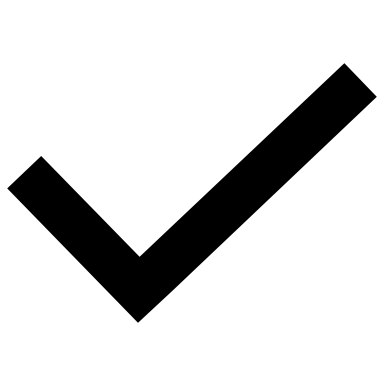 |
| Sample size of at least 100 participants^b^ | 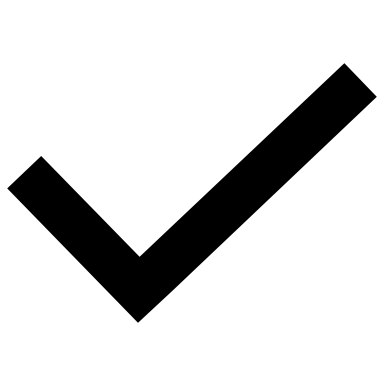 | 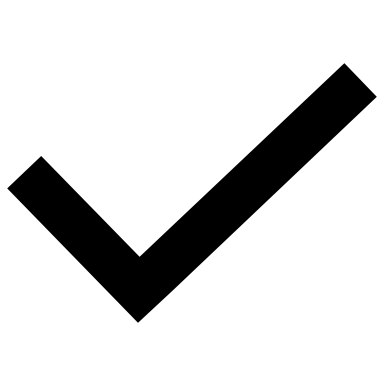 |
| All participants had T2DM | 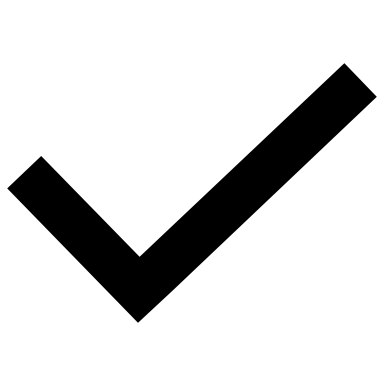 | 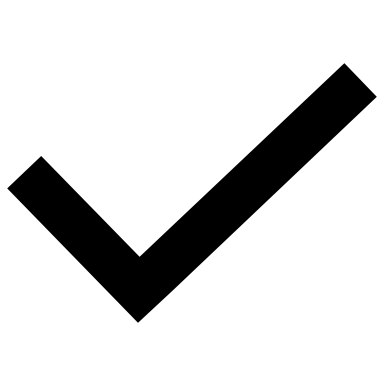 |
| Participants enrolled from at least two countries^c^ | 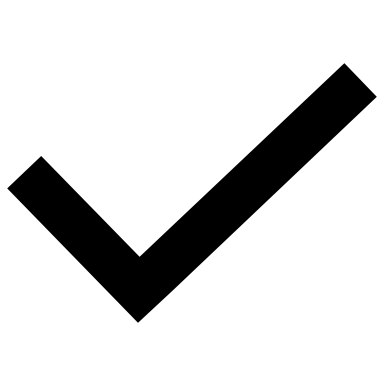 |  |
| Published in or after the year 2000 | 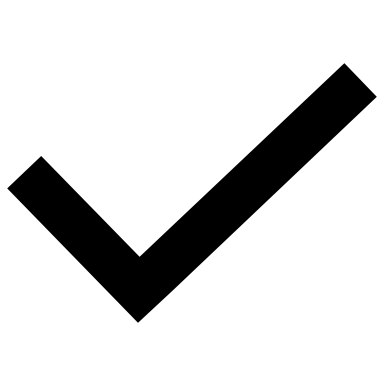 | 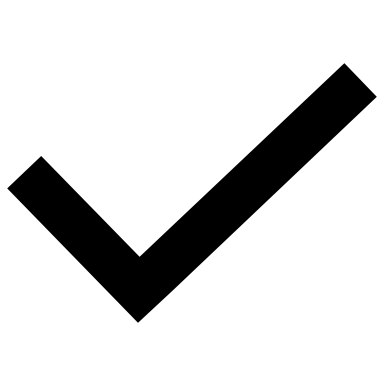 |
| English language | 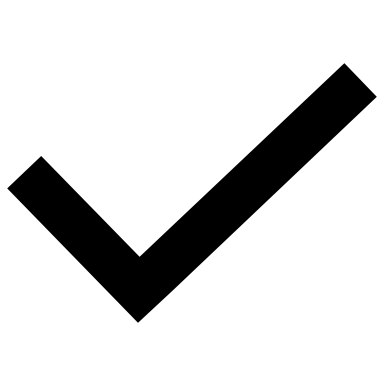 | 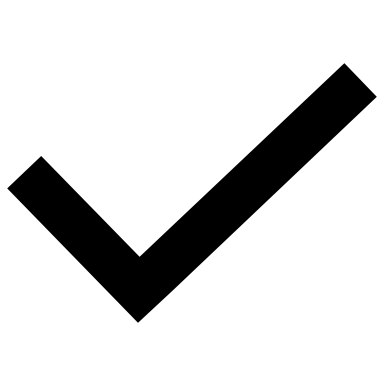 |

a) The IF criterion was lowered for government trials to allow a greater number of government-funded studies to be captured in the search.
b) A sample size of at least 100 participants was required to help ensure the selection of the most robust findings and avoid small study bias.

Government trials were not held to the same multi-country standard as industry trials because they were typically conducted within a single country.
