## Supplementary material for "Assessing non-white ethnic participation in type 2 diabetes mellitus randomized clinical trials: A Meta-Analysis": eTable 3

**eTable 3. List of Excluded Studies After Full Text Review**

| **Study** | **Reason for exclusion** |
| --- | --- |
| Clerici 2011 Novel soy germ pasta improves endothelial function, blood pressure, and oxidative stress in patients with type 2 diabetes Clerici, C.; Nardi, E.; Battezzati, P. M.; Asciutti, S.; Castellani, D.; Corazzi, N.; Giuliano, V.; Gizzi, S.; Perriello, G.; Di Matteo, G.; Galli, F.; Setchell, K. D. Diabetes Care Sep 2011;34(9):1946-8 | <100 Subjects |
| Larsen 2001 Glucagon-like peptide-1 infusion must be maintained for 24 h/day to obtain acceptable glycemia in type 2 diabetic patients who are poorly controlled on sulphonylurea treatment Larsen, J.; Hylleberg, B.; Ng, K.; Damsbo, P.  Diabetes Care Aug 2001;24(8):1416-21 | <100 Subjects |
| Lund 2009 Combining insulin with metformin or an insulin secretagogue in non-obese patients with type 2 diabetes: 12 month, randomised, double blind trial Lund, S. S.; Tarnow, L.; Frandsen, M.; Nielsen, B. B.; Hansen, B. V.; Pedersen, O.; Parving, H. H.; Vaag, A. A.  Bmj Nov 9 2009;339():b4324 | <100 Subjects |
| Herman 2005 A clinical trial of continuous subcutaneous insulin infusion versus multiple daily injections in older adults with type 2 diabetes Herman, W. H.; Ilag, L. L.; Johnson, S. L.; Martin, C. L.; Sinding, J.; Al Harthi, A.; Plunkett, C. D.; LaPorte, F. B.; Burke, R.; Brown, M. B.; Halter, J. B.; Raskin, P.  Diabetes Care Jul 2005;28(7):1568-73 | <100 Subjects |
| Wang 2010 Dose-response effects of insulin glargine in type 2 diabetes Wang, Z.; Hedrington, M. S.; Gogitidze Joy, N.; Briscoe, V. J.; Richardson, M. A.; Younk, L.; Nicholson, W.; Tate, D. B.; Davis, S. N.  Diabetes Care Jul 2010;33(7):1555-60 | <100 Subjects |
| Mingrone 2015 Bariatric-metabolic surgery versus conventional medical treatment in obese patients with type 2 diabetes: 5 year follow-up of an open-label, single-centre, randomised controlled trial Mingrone, G.; Panunzi, S.; De Gaetano, A.; Guidone, C.; Iaconelli, A.; Nanni, G.; Castagneto, M.; Bornstein, S.; Rubino, F.  Lancet Sep 5 2015;386(9997):964-73 | <100 Subjects |
| Johansen 2017 Effect of an Intensive Lifestyle Intervention on Glycemic Control in Patients With Type 2 Diabetes: A Randomized Clinical Trial Johansen, M. Y.; MacDonald, C. S.; Hansen, K. B.; Karstoft, K.; Christensen, R.; Pedersen, M.; Hansen, L. S.; Zacho, M.; Wedell-Neergaard, A. S.; Nielsen, S. T.; Iepsen, U. W.; Langberg, H.; Vaag, A. A.; Pedersen, B. K.; Ried-Larsen, M.  Jama Aug 15 2017;318(7):637-646 | <100 Subjects |
| Elbrønd 2002 Pharmacokinetics, pharmacodynamics, safety, and tolerability of a single-dose of NN2211, a long-acting glucagon-like peptide 1 derivative, in healthy male subjects Elbrønd, B.; Jakobsen, G.; Larsen, S.; Agersø, H.; Jensen, L. B.; Rolan, P.; Sturis, J.; Hatorp, V.; Zdravkovic, M.  Diabetes Care Aug 2002;25(8):1398-404 | <100 Subjects |
| Szendroedi 2009 Effects of high-dose simvastatin therapy on glucose metabolism and ectopic lipid deposition in nonobese type 2 diabetic patients Szendroedi, J.; Anderwald, C.; Krssak, M.; Bayerle-Eder, M.; Esterbauer, H.; Pfeiler, G.; Brehm, A.; Nowotny, P.; Hofer, A.; Waldhäusl, W.; Roden, M.  Diabetes Care Feb 2009;32(2):209-14 | <100 Subjects |
| Ueno 2014 Exploratory trial of intranasal administration of glucagon-like peptide-1 in Japanese patients with type 2 diabetes Ueno, H.; Mizuta, M.; Shiiya, T.; Tsuchimochi, W.; Noma, K.; Nakashima, N.; Fujihara, M.; Nakazato, M.  Diabetes Care Jul 2014;37(7):2024-7 | <100 Subjects |
| Mansourati 2001 Lipid lowering does not improve endothelial function in subjects with poorly controlled diabetes Mansourati, J.; Newman, L. G.; Roman, S. H.; Travis, A.; Rafey, M.; Phillips, R. A.  Diabetes Care Dec 2001;24(12):2152-3 | <100 Subjects |
| Kadoglou 2007 Beneficial effects of combined treatment with rosiglitazone and exercise on cardiovascular risk factors in patients with type 2 diabetes Kadoglou, N. P.; Iliadis, F.; Liapis, C. D.; Perrea, D.; Angelopoulou, N.; Alevizos, M.  Diabetes Care Sep 2007;30(9):2242-4 | <100 Subjects |
| Moran 2009 Insulin therapy to improve BMI in cystic fibrosis-related diabetes without fasting hyperglycemia: results of the cystic fibrosis related diabetes therapy trial Moran, A.; Pekow, P.; Grover, P.; Zorn, M.; Slovis, B.; Pilewski, J.; Tullis, E.; Liou, T. G.; Allen, H.  Diabetes Care Oct 2009;32(10):1783-8 | less than 100 participants |
| Broedbaek 2011 Long-term effects of Irbesartan treatment and smoking on nucleic acid oxidation in patients with type 2 diabetes and microalbuminuria: an Irbesartan in patients with type 2 diabetes and Microalbuminuria (IRMA 2) substudy Broedbaek, K.; Henriksen, T.; Weimann, A.; Petersen, M.; Andersen, J. T.; Afzal, S.; Jimenez-Solem, E.; Persson, F.; Parving, H. H.; Rossing, P.; Poulsen, H. E.  Diabetes Care May 2011;34(5):1192-8 | less than 100 participants |
| Vakkilainen 2002 Fenofibrate lowers plasma triglycerides and increases LDL particle diameter in subjects with type 2 diabetes Vakkilainen, J.; Steiner, G.; Ansquer, J. C.; Perttunen-Nio, H.; Taskinen, M. R.  Diabetes Care Mar 2002;25(3):627-8 | less than 100 participants |
| Ancillary Publication | 105 |
| Levin 2000 Effect of intensive glycemic control on microalbuminuria in type 2 diabetes. Veterans Affairs Cooperative Study on Glycemic Control and Complications in Type 2 Diabetes Feasibility Trial Investigators Levin, S. R.; Coburn, J. W.; Abraira, C.; Henderson, W. G.; Colwell, J. A.; Emanuele, N. V.; Nuttall, F. Q.; Sawin, C. T.; Comstock, J. P.; Silbert, C. K.  Diabetes Care Oct 2000;23(10):1478-85 | Ancillary Publication |
| Navarro-González 2018 Effects of Pentoxifylline on Soluble Klotho Concentrations and Renal Tubular Cell Expression in Diabetic Kidney Disease Navarro-González, J. F.; Sánchez-Niño, M. D.; Donate-Correa, J.; Martín-Núñez, E.; Ferri, C.; Pérez-Delgado, N.; Górriz, J. L.; Martínez-Castelao, A.; Ortiz, A.; Mora-Fernández, C.  Diabetes Care Aug 2018;41(8):1817-1820 | Ancillary Publication |
| Saito 2017 Low-Dose Aspirin for Primary Prevention of Cardiovascular Events in Patients With Type 2 Diabetes Mellitus: 10-Year Follow-Up of a Randomized Controlled Trial Saito, Y.; Okada, S.; Ogawa, H.; Soejima, H.; Sakuma, M.; Nakayama, M.; Doi, N.; Jinnouchi, H.; Waki, M.; Masuda, I.; Morimoto, T.  Circulation Feb 14 2017;135(7):659-670 | Ancillary Publication |
| Bethel 2019 Changes in Serum Calcitonin Concentrations, Incidence of Medullary Thyroid Carcinoma, and Impact of Routine Calcitonin Concentration Monitoring in the EXenatide Study of Cardiovascular Event Lowering (EXSCEL) Bethel, M. A.; Patel, R. A.; Thompson, V. P.; Merrill, P.; Reed, S. D.; Li, Y.; Ahmadi, S.; Katona, B. G.; Gustavson, S. M.; Ohman, P.; Iqbal, N.; Gagel, R. F.; Hernandez, A. F.; Buse, J. B.; Holman, R. R.  Diabetes Care Jun 2019;42(6):1075-1080 | Ancillary Publication |
| Koska 2013 The effect of intensive glucose lowering on lipoprotein particle profiles and inflammatory markers in the Veterans Affairs Diabetes Trial (VADT) Koska, J.; Saremi, A.; Bahn, G.; Yamashita, S.; Reaven, P. D.  Diabetes Care Aug 2013;36(8):2408-14 | Ancillary Publication |
| Agrawal 2011 Observation on renal outcomes in the Veterans Affairs Diabetes Trial Agrawal, L.; Azad, N.; Emanuele, N. V.; Bahn, G. D.; Kaufman, D. G.; Moritz, T. E.; Duckworth, W. C.; Abraira, C.  Diabetes Care Sep 2011;34(9):2090-4 | Ancillary Publication |
| Saito 2011 Low-dose aspirin therapy in patients with type 2 diabetes and reduced glomerular filtration rate: subanalysis from the JPAD trial Saito, Y.; Morimoto, T.; Ogawa, H.; Nakayama, M.; Uemura, S.; Doi, N.; Jinnouchi, H.; Waki, M.; Soejima, H.; Sugiyama, S.; Okada, S.; Akai, Y.  Diabetes Care Feb 2011;34(2):280-5 | Ancillary Publication |
| Johansen 2012 Effect of intensive multifactorial treatment compared with routine care on aortic stiffness and central blood pressure among individuals with screen-detected type 2 diabetes: the ADDITION-Denmark study Johansen, N. B.; Charles, M.; Vistisen, D.; Rasmussen, S. S.; Wiinberg, N.; Borch-Johnsen, K.; Lauritzen, T.; Sandbæk, A.; Witte, D. R.  Diabetes Care Nov 2012;35(11):2207-14 | Ancillary Publication |
| Abdul-Ghani 2021 Durability of Triple Combination Therapy Versus Stepwise Addition Therapy in Patients With New-Onset T2DM: 3-Year Follow-up of EDICT Abdul-Ghani, M.; Puckett, C.; Adams, J.; Khattab, A.; Baskoy, G.; Cersosimo, E.; Triplitt, C.; DeFronzo, R. A.  Diabetes Care Feb 2021;44(2):433-439 | Ancillary Publication |
| Okada 2018 Effect of Aspirin on Cancer Chemoprevention in Japanese Patients With Type 2 Diabetes: 10-Year Observational Follow-up of a Randomized Controlled Trial Okada, S.; Morimoto, T.; Ogawa, H.; Sakuma, M.; Matsumoto, C.; Soejima, H.; Nakayama, M.; Doi, N.; Jinnouchi, H.; Waki, M.; Masuda, I.; Saito, Y.  Diabetes Care Aug 2018;41(8):1757-1764 | Ancillary Publication |
| Okada 2011 Differential effect of low-dose aspirin for primary prevention of atherosclerotic events in diabetes management: a subanalysis of the JPAD trial Okada, S.; Morimoto, T.; Ogawa, H.; Kanauchi, M.; Nakayama, M.; Uemura, S.; Doi, N.; Jinnouchi, H.; Waki, M.; Soejima, H.; Sakuma, M.; Saito, Y.  Diabetes Care Jun 2011;34(6):1277-83 | Ancillary Publication |
| Leinonen 2002 Reduced IGFBP-1 is associated with thickening of the carotid wall in type 2 diabetes Leinonen, E. S.; Salonen, J. T.; Salonen, R. M.; Koistinen, R. A.; Leinonen, P. J.; Sarna, S. S.; Taskinen, M. R.  Diabetes Care Oct 2002;25(10):1807-12 | Ancillary Publication |
| Tripathy 2014 Baseline adiponectin levels do not influence the response to pioglitazone in ACT NOW Tripathy, D.; Clement, S. C.; Schwenke, D. C.; Banerji, M.; Bray, G. A.; Buchanan, T. A.; Gastaldelli, A.; Henry, R. R.; Kitabchi, A. E.; Mudaliar, S.; Ratner, R. E.; Stentz, F. B.; Musi, N.; Reaven, P. D.; DeFronzo, R. A.  Diabetes Care Jun 2014;37(6):1706-11 | Ancillary Publication |
| Anderson 2011 Blood pressure and cardiovascular disease risk in the Veterans Affairs Diabetes Trial Anderson, R. J.; Bahn, G. D.; Moritz, T. E.; Kaufman, D.; Abraira, C.; Duckworth, W.  Diabetes Care Jan 2011;34(1):34-8 | Ancillary Publication |
| Kato 2019 Effect of Dapagliflozin on Heart Failure and Mortality in Type 2 Diabetes Mellitus Kato, E. T.; Silverman, M. G.; Mosenzon, O.; Zelniker, T. A.; Cahn, A.; Furtado, R. H. M.; Kuder, J.; Murphy, S. A.; Bhatt, D. L.; Leiter, L. A.; McGuire, D. K.; Wilding, J. P. H.; Bonaca, M. P.; Ruff, C. T.; Desai, A. S.; Goto, S.; Johansson, P. A.; Gause-Nilsson, I.; Johanson, P.; Langkilde, A. M.; Raz, I.; Sabatine, M. S.; Wiviott, S. D.  Circulation May 28 2019;139(22):2528-2536 | Ancillary Publication |
| Everett 2015 Troponin and Cardiac Events in Stable Ischemic Heart Disease and Diabetes Everett, B. M.; Brooks, M. M.; Vlachos, H. E.; Chaitman, B. R.; Frye, R. L.; Bhatt, D. L.  N Engl J Med Aug 13 2015;373(7):610-20 | Ancillary Publication |
| Leiter 2015 Canagliflozin provides durable glycemic improvements and body weight reduction over 104 weeks versus glimepiride in patients with type 2 diabetes on metformin: a randomized, double-blind, phase 3 study Leiter, L. A.; Yoon, K. H.; Arias, P.; Langslet, G.; Xie, J.; Balis, D. A.; Millington, D.; Vercruysse, F.; Canovatchel, W.; Meininger, G.  Diabetes Care Mar 2015;38(3):355-64 | Ancillary Publication |
| Hegedüs 2018 No Evidence of Increase in Calcitonin Concentrations or Development of C-Cell Malignancy in Response to Liraglutide for Up to 5 Years in the LEADER Trial Hegedüs, L.; Sherman, S. I.; Tuttle, R. M.; von Scholten, B. J.; Rasmussen, S.; Karsbøl, J. D.; Daniels, G. H.  Diabetes Care Mar 2018;41(3):620-622 | Ancillary Publication |
| Cahn 2020 Efficacy and Safety of Dapagliflozin in the Elderly: Analysis From the DECLARE-TIMI 58 Study Cahn, A.; Mosenzon, O.; Wiviott, S. D.; Rozenberg, A.; Yanuv, I.; Goodrich, E. L.; Murphy, S. A.; Bhatt, D. L.; Leiter, L. A.; McGuire, D. K.; Wilding, J. P. H.; Gause-Nilsson, I. A. M.; Fredriksson, M.; Johansson, P. A.; Langkilde, A. M.; Sabatine, M. S.; Raz, I.  Diabetes Care Feb 2020;43(2):468-475 | Ancillary Publication |
| Pop-Busui 2013 Impact of glycemic control strategies on the progression of diabetic peripheral neuropathy in the Bypass Angioplasty Revascularization Investigation 2 Diabetes (BARI 2D) Cohort Pop-Busui, R.; Lu, J.; Brooks, M. M.; Albert, S.; Althouse, A. D.; Escobedo, J.; Green, J.; Palumbo, P.; Perkins, B. A.; Whitehouse, F.; Jones, T. L.  Diabetes Care Oct 2013;36(10):3208-15 | Ancillary Publication |
| Cavender 2017 Serial Measurement of High-Sensitivity Troponin I and Cardiovascular Outcomes in Patients With Type 2 Diabetes Mellitus in the EXAMINE Trial (Examination of Cardiovascular Outcomes With Alogliptin Versus Standard of Care) Cavender, M. A.; White, W. B.; Jarolim, P.; Bakris, G. L.; Cushman, W. C.; Kupfer, S.; Gao, Q.; Mehta, C. R.; Zannad, F.; Cannon, C. P.; Morrow, D. A.  Circulation May 16 2017;135(20):1911-1921 | Ancillary Publication |
| Standl 2018 Increased Risk of Severe Hypoglycemic Events Before and After Cardiovascular Outcomes in TECOS Suggests an At-Risk Type 2 Diabetes Frail Patient Phenotype Standl, E.; Stevens, S. R.; Armstrong, P. W.; Buse, J. B.; Chan, J. C. N.; Green, J. B.; Lachin, J. M.; Scheen, A.; Travert, F.; Van de Werf, F.; Peterson, E. D.; Holman, R. R.  Diabetes Care Mar 2018;41(3):596-603 | Ancillary Publication |
| O'Connor 2012 Effect of intensive versus standard blood pressure control on depression and health-related quality of life in type 2 diabetes: the ACCORD trial O'Connor, P. J.; Narayan, K. M.; Anderson, R.; Feeney, P.; Fine, L.; Ali, M. K.; Simmons, D. L.; Hire, D. G.; Sperl-Hillen, J. M.; Katz, L. A.; Margolis, K. L.; Sullivan, M. D.  Diabetes Care Jul 2012;35(7):1479-81 | Ancillary Publication |
| Mosenzon 2017 Effect of Saxagliptin on Renal Outcomes in the SAVOR-TIMI 53 Trial Mosenzon, O.; Leibowitz, G.; Bhatt, D. L.; Cahn, A.; Hirshberg, B.; Wei, C.; Im, K.; Rozenberg, A.; Yanuv, I.; Stahre, C.; Ray, K. K.; Iqbal, N.; Braunwald, E.; Scirica, B. M.; Raz, I.  Diabetes Care Jan 2017;40(1):69-76 | Ancillary Publication |
| Biessels 2019 Effect of Linagliptin on Cognitive Performance in Patients With Type 2 Diabetes and Cardiorenal Comorbidities: The CARMELINA Randomized Trial Biessels, G. J.; Verhagen, C.; Janssen, J.; van den Berg, E.; Zinman, B.; Rosenstock, J.; George, J. T.; Passera, A.; Schnaidt, S.; Johansen, O. E.  Diabetes Care Oct 2019;42(10):1930-1938 | Ancillary Publication |
| Zinman 2018 Hypoglycemia, Cardiovascular Outcomes, and Death: The LEADER Experience Zinman, B.; Marso, S. P.; Christiansen, E.; Calanna, S.; Rasmussen, S.; Buse, J. B.  Diabetes Care Aug 2018;41(8):1783-1791 | Ancillary Publication |
| Margolis 2014 Outcomes of combined cardiovascular risk factor management strategies in type 2 diabetes: the ACCORD randomized trial Margolis, K. L.; O'Connor, P. J.; Morgan, T. M.; Buse, J. B.; Cohen, R. M.; Cushman, W. C.; Cutler, J. A.; Evans, G. W.; Gerstein, H. C.; Grimm, R. H., Jr.; Lipkin, E. W.; Narayan, K. M.; Riddle, M. C., Jr.; Sood, A.; Goff, D. C., Jr.  Diabetes Care Jun 2014;37(6):1721-8 | Ancillary Publication |
| Bach 2013 Rosiglitazone and outcomes for patients with diabetes mellitus and coronary artery disease in the Bypass Angioplasty Revascularization Investigation 2 Diabetes (BARI 2D) trial Bach, R. G.; Brooks, M. M.; Lombardero, M.; Genuth, S.; Donner, T. W.; Garber, A.; Kennedy, L.; Monrad, E. S.; Pop-Busui, R.; Kelsey, S. F.; Frye, R. L.  Circulation Aug 20 2013;128(8):785-94 | Ancillary Publication |
| McGuire 2019 Linagliptin Effects on Heart Failure and Related Outcomes in Individuals With Type 2 Diabetes Mellitus at High Cardiovascular and Renal Risk in CARMELINA McGuire, D. K.; Alexander, J. H.; Johansen, O. E.; Perkovic, V.; Rosenstock, J.; Cooper, M. E.; Wanner, C.; Kahn, S. E.; Toto, R. D.; Zinman, B.; Baanstra, D.; Pfarr, E.; Schnaidt, S.; Meinicke, T.; George, J. T.; von Eynatten, M.; Marx, N.  Circulation Jan 15 2019;139(3):351-361 | Ancillary Publication |
| Nauck 2019 Effects of Liraglutide Compared With Placebo on Events of Acute Gallbladder or Biliary Disease in Patients With Type 2 Diabetes at High Risk for Cardiovascular Events in the LEADER Randomized Trial Nauck, M. A.; Muus Ghorbani, M. L.; Kreiner, E.; Saevereid, H. A.; Buse, J. B.  Diabetes Care Oct 2019;42(10):1912-1920 | Ancillary Publication |
| Zannad 2015 Heart failure and mortality outcomes in patients with type 2 diabetes taking alogliptin versus placebo in EXAMINE: a multicentre, randomised, double-blind trial Zannad, F.; Cannon, C. P.; Cushman, W. C.; Bakris, G. L.; Menon, V.; Perez, A. T.; Fleck, P. R.; Mehta, C. R.; Kupfer, S.; Wilson, C.; Lam, H.; White, W. B.  Lancet May 23 2015;385(9982):2067-76 | Ancillary Publication |
| Brooks 2010 Health status after treatment for coronary artery disease and type 2 diabetes mellitus in the Bypass Angioplasty Revascularization Investigation 2 Diabetes trial Brooks, M. M.; Chung, S. C.; Helmy, T.; Hillegass, W. B.; Escobedo, J.; Melsop, K. A.; Massaro, E. M.; McBane, R. D.; Hyde, P.; Hlatky, M. A.  Circulation Oct 26 2010;122(17):1690-9 | Ancillary Publication |
| Miller 2014 Effects of randomization to intensive glucose control on adverse events, cardiovascular disease, and mortality in older versus younger adults in the ACCORD Trial Miller, M. E.; Williamson, J. D.; Gerstein, H. C.; Byington, R. P.; Cushman, W. C.; Ginsberg, H. N.; Ambrosius, W. T.; Lovato, L.; Applegate, W. B.  Diabetes Care 2014;37(3):634-43 | Ancillary Publication |
| Cosentino 2020 Efficacy of Ertugliflozin on Heart Failure-Related Events in Patients With Type 2 Diabetes Mellitus and Established Atherosclerotic Cardiovascular Disease: Results of the VERTIS CV Trial Cosentino, F.; Cannon, C. P.; Cherney, D. Z. I.; Masiukiewicz, U.; Pratley, R.; Dagogo-Jack, S.; Frederich, R.; Charbonnel, B.; Mancuso, J.; Shih, W. J.; Terra, S. G.; Cater, N. B.; Gantz, I.; McGuire, D. K.  Circulation Dec 8 2020;142(23):2205-2215 | Ancillary Publication |
| Udell 2015 Saxagliptin and cardiovascular outcomes in patients with type 2 diabetes and moderate or severe renal impairment: observations from the SAVOR-TIMI 53 Trial Udell, J. A.; Bhatt, D. L.; Braunwald, E.; Cavender, M. A.; Mosenzon, O.; Steg, P. G.; Davidson, J. A.; Nicolau, J. C.; Corbalan, R.; Hirshberg, B.; Frederich, R.; Im, K.; Umez-Eronini, A. A.; He, P.; McGuire, D. K.; Leiter, L. A.; Raz, I.; Scirica, B. M.  Diabetes Care Apr 2015;38(4):696-705 | Ancillary Publication |
| Scirica 2014 Heart failure, saxagliptin, and diabetes mellitus: observations from the SAVOR-TIMI 53 randomized trial Scirica, B. M.; Braunwald, E.; Raz, I.; Cavender, M. A.; Morrow, D. A.; Jarolim, P.; Udell, J. A.; Mosenzon, O.; Im, K.; Umez-Eronini, A. A.; Pollack, P. S.; Hirshberg, B.; Frederich, R.; Lewis, B. S.; McGuire, D. K.; Davidson, J.; Steg, P. G.; Bhatt, D. L.  Circulation Oct 28 2014;130(18):1579-88 | Ancillary Publication |
| Wanner 2016 Empagliflozin and Progression of Kidney Disease in Type 2 Diabetes Wanner, C.; Inzucchi, S. E.; Lachin, J. M.; Fitchett, D.; von Eynatten, M.; Mattheus, M.; Johansen, O. E.; Woerle, H. J.; Broedl, U. C.; Zinman, B.  N Engl J Med Jul 28 2016;375(4):323-34 | Ancillary Publication |
| Ismail-Beigi 2010 Effect of intensive treatment of hyperglycaemia on microvascular outcomes in type 2 diabetes: an analysis of the ACCORD randomised trial Ismail-Beigi, F.; Craven, T.; Banerji, M. A.; Basile, J.; Calles, J.; Cohen, R. M.; Cuddihy, R.; Cushman, W. C.; Genuth, S.; Grimm, R. H., Jr.; Hamilton, B. P.; Hoogwerf, B.; Karl, D.; Katz, L.; Krikorian, A.; O'Connor, P.; Pop-Busui, R.; Schubart, U.; Simmons, D.; Taylor, H.; Thomas, A.; Weiss, D.; Hramiak, I.  Lancet Aug 7 2010;376(9739):419-30 | Ancillary Publication |
| Raz 2014 Incidence of pancreatitis and pancreatic cancer in a randomized controlled multicenter trial (SAVOR-TIMI 53) of the dipeptidyl peptidase-4 inhibitor saxagliptin Raz, I.; Bhatt, D. L.; Hirshberg, B.; Mosenzon, O.; Scirica, B. M.; Umez-Eronini, A.; Im, K.; Stahre, C.; Buskila, A.; Iqbal, N.; Greenberger, N.; Lerch, M. M.  Diabetes Care Sep 2014;37(9):2435-41 | Ancillary Publication |
| Dagenais 2011 Effects of optimal medical treatment with or without coronary revascularization on angina and subsequent revascularizations in patients with type 2 diabetes mellitus and stable ischemic heart disease Dagenais, G. R.; Lu, J.; Faxon, D. P.; Kent, K.; Lago, R. M.; Lezama, C.; Hueb, W.; Weiss, M.; Slater, J.; Frye, R. L.  Circulation Apr 12 2011;123(14):1492-500 | Ancillary Publication |
| Gerstein 2011 Long-term effects of intensive glucose lowering on cardiovascular outcomes Gerstein, H. C.; Miller, M. E.; Genuth, S.; Ismail-Beigi, F.; Buse, J. B.; Goff, D. C., Jr.; Probstfield, J. L.; Cushman, W. C.; Ginsberg, H. N.; Bigger, J. T.; Grimm, R. H., Jr.; Byington, R. P.; Rosenberg, Y. D.; Friedewald, W. T.  N Engl J Med Mar 3 2011;364(9):818-28 | Ancillary Publication |
| Mosenzon 2015 Incidence of Fractures in Patients With Type 2 Diabetes in the SAVOR-TIMI 53 Trial Mosenzon, O.; Wei, C.; Davidson, J.; Scirica, B. M.; Yanuv, I.; Rozenberg, A.; Hirshberg, B.; Cahn, A.; Stahre, C.; Strojek, K.; Bhatt, D. L.; Raz, I.  Diabetes Care Nov 2015;38(11):2142-50 | Ancillary Publication |
| Mychaleckyj 2012 Reversibility of fenofibrate therapy-induced renal function impairment in ACCORD type 2 diabetic participants Mychaleckyj, J. C.; Craven, T.; Nayak, U.; Buse, J.; Crouse, J. R.; Elam, M.; Kirchner, K.; Lorber, D.; Marcovina, S.; Sivitz, W.; Sperl-Hillen, J.; Bonds, D. E.; Ginsberg, H. N.  Diabetes Care May 2012;35(5):1008-14 | Ancillary Publication |
| Furtado 2019 Dapagliflozin and Cardiovascular Outcomes in Patients With Type 2 Diabetes Mellitus and Previous Myocardial Infarction Furtado, R. H. M.; Bonaca, M. P.; Raz, I.; Zelniker, T. A.; Mosenzon, O.; Cahn, A.; Kuder, J.; Murphy, S. A.; Bhatt, D. L.; Leiter, L. A.; McGuire, D. K.; Wilding, J. P. H.; Ruff, C. T.; Nicolau, J. C.; Gause-Nilsson, I. A. M.; Fredriksson, M.; Langkilde, A. M.; Sabatine, M. S.; Wiviott, S. D.  Circulation May 28 2019;139(22):2516-2527 | Ancillary Publication |
| Leiter 2015 Efficacy and safety of saxagliptin in older participants in the SAVOR-TIMI 53 trial Leiter, L. A.; Teoh, H.; Braunwald, E.; Mosenzon, O.; Cahn, A.; Kumar, K. M.; Smahelova, A.; Hirshberg, B.; Stahre, C.; Frederich, R.; Bonnici, F.; Scirica, B. M.; Bhatt, D. L.; Raz, I.  Diabetes Care Jun 2015;38(6):1145-53 | Ancillary Publication |
| Dhatariya 2018 The Impact of Liraglutide on Diabetes-Related Foot Ulceration and Associated Complications in Patients With Type 2 Diabetes at High Risk for Cardiovascular Events: Results From the LEADER Trial Dhatariya, K.; Bain, S. C.; Buse, J. B.; Simpson, R.; Tarnow, L.; Kaltoft, M. S.; Stellfeld, M.; Tornøe, K.; Pratley, R. E.  Diabetes Care Oct 2018;41(10):2229-2235 | Ancillary Publication |
| Buse 2017 Pancreatic Safety of Sitagliptin in the TECOS Study Buse, J. B.; Bethel, M. A.; Green, J. B.; Stevens, S. R.; Lokhnygina, Y.; Aschner, P.; Grado, C. R.; Tankova, T.; Wainstein, J.; Josse, R.; Lachin, J. M.; Engel, S. S.; Patel, K.; Peterson, E. D.; Holman, R. R.  Diabetes Care Feb 2017;40(2):164-170 | Ancillary Publication |
| Ting 2012 Benefits and safety of long-term fenofibrate therapy in people with type 2 diabetes and renal impairment: the FIELD Study Ting, R. D.; Keech, A. C.; Drury, P. L.; Donoghoe, M. W.; Hedley, J.; Jenkins, A. J.; Davis, T. M.; Lehto, S.; Celermajer, D.; Simes, R. J.; Rajamani, K.; Stanton, K.  Diabetes Care Feb 2012;35(2):218-25 | Ancillary Publication |
| Verma 2018 Effect of Liraglutide on Cardiovascular Events in Patients With Type 2 Diabetes Mellitus and Polyvascular Disease: Results of the LEADER Trial Verma, S.; Bhatt, D. L.; Bain, S. C.; Buse, J. B.; Mann, J. F. E.; Marso, S. P.; Nauck, M. A.; Poulter, N. R.; Pratley, R. E.; Zinman, B.; Michelsen, M. M.; Monk Fries, T.; Rasmussen, S.; Leiter, L. A.  Circulation May 15 2018;137(20):2179-2183 | Ancillary Publication |
| Reyes-Soffer 2013 Effect of combination therapy with fenofibrate and simvastatin on postprandial lipemia in the ACCORD lipid trial Reyes-Soffer, G.; Ngai, C. I.; Lovato, L.; Karmally, W.; Ramakrishnan, R.; Holleran, S.; Ginsberg, H. N.  Diabetes Care Feb 2013;36(2):422-8 | Ancillary Publication |
| Verma 2018 Effects of Liraglutide on Cardiovascular Outcomes in Patients With Type 2 Diabetes Mellitus With or Without History of Myocardial Infarction or Stroke Verma, S.; Poulter, N. R.; Bhatt, D. L.; Bain, S. C.; Buse, J. B.; Leiter, L. A.; Nauck, M. A.; Pratley, R. E.; Zinman, B.; Ørsted, D. D.; Monk Fries, T.; Rasmussen, S.; Marso, S. P.  Circulation Dec 18 2018;138(25):2884-2894 | Ancillary Publication |
| Anderson 2011 Effect of intensive glycemic lowering on health-related quality of life in type 2 diabetes: ACCORD trial Anderson, R. T.; Narayan, K. M.; Feeney, P.; Goff, D., Jr.; Ali, M. K.; Simmons, D. L.; Sperl-Hillen, J. A.; Bigger, T.; Cuddihy, R.; O'Conner, P. J.; Sood, A.; Zhang, P.; Sullivan, M. D.  Diabetes Care Apr 2011;34(4):807-12 | Ancillary Publication |
| Mahaffey 2019 Canagliflozin and Cardiovascular and Renal Outcomes in Type 2 Diabetes Mellitus and Chronic Kidney Disease in Primary and Secondary Cardiovascular Prevention Groups Mahaffey, K. W.; Jardine, M. J.; Bompoint, S.; Cannon, C. P.; Neal, B.; Heerspink, H. J. L.; Charytan, D. M.; Edwards, R.; Agarwal, R.; Bakris, G.; Bull, S.; Capuano, G.; de Zeeuw, D.; Greene, T.; Levin, A.; Pollock, C.; Sun, T.; Wheeler, D. C.; Yavin, Y.; Zhang, H.; Zinman, B.; Rosenthal, N.; Brenner, B. M.; Perkovic, V.  Circulation Aug 27 2019;140(9):739-750 | Ancillary Publication |
| Steinberg 2017 Amylase, Lipase, and Acute Pancreatitis in People With Type 2 Diabetes Treated With Liraglutide: Results From the LEADER Randomized Trial Steinberg, W. M.; Buse, J. B.; Ghorbani, M. L. M.; Ørsted, D. D.; Nauck, M. A.  Diabetes Care Jul 2017;40(7):966-972 | Ancillary Publication |
| Rådholm 2018 Canagliflozin and Heart Failure in Type 2 Diabetes Mellitus: Results From the CANVAS Program Rådholm, K.; Figtree, G.; Perkovic, V.; Solomon, S. D.; Mahaffey, K. W.; de Zeeuw, D.; Fulcher, G.; Barrett, T. D.; Shaw, W.; Desai, M.; Matthews, D. R.; Neal, B.  Circulation Jul 31 2018;138(5):458-468 | Ancillary Publication |
| Nauck 2018 Neoplasms Reported With Liraglutide or Placebo in People With Type 2 Diabetes: Results From the LEADER Randomized Trial Nauck, M. A.; Jensen, T. J.; Rosenkilde, C.; Calanna, S.; Buse, J. B.  Diabetes Care Aug 2018;41(8):1663-1671 | Ancillary Publication |
| Forsblom 2010 Effects of long-term fenofibrate treatment on markers of renal function in type 2 diabetes: the FIELD Helsinki substudy Forsblom, C.; Hiukka, A.; Leinonen, E. S.; Sundvall, J.; Groop, P. H.; Taskinen, M. R.  Diabetes Care Feb 2010;33(2):215-20 | Ancillary Publication |
| Gerstein 2014 Effects of intensive glycaemic control on ischaemic heart disease: analysis of data from the randomised, controlled ACCORD trial Gerstein, H. C.; Miller, M. E.; Ismail-Beigi, F.; Largay, J.; McDonald, C.; Lochnan, H. A.; Booth, G. L.  Lancet Nov 29 2014;384(9958):1936-41 | Ancillary Publication |
| Cannon 2020 Evaluating the Effects of Canagliflozin on Cardiovascular and Renal Events in Patients With Type 2 Diabetes Mellitus and Chronic Kidney Disease According to Baseline HbA1c, Including Those With HbA1c <7%: Results From the CREDENCE Trial Cannon, C. P.; Perkovic, V.; Agarwal, R.; Baldassarre, J.; Bakris, G.; Charytan, D. M.; de Zeeuw, D.; Edwards, R.; Greene, T.; Heerspink, H. J. L.; Jardine, M. J.; Levin, A.; Li, J. W.; Neal, B.; Pollock, C.; Wheeler, D. C.; Zhang, H.; Zinman, B.; Mahaffey, K. W.  Circulation Feb 4 2020;141(5):407-410 | Ancillary Publication |
| Bethel 2017 Assessing the Safety of Sitagliptin in Older Participants in the Trial Evaluating Cardiovascular Outcomes with Sitagliptin (TECOS) Bethel, M. A.; Engel, S. S.; Green, J. B.; Huang, Z.; Josse, R. G.; Kaufman, K. D.; Standl, E.; Suryawanshi, S.; Van de Werf, F.; McGuire, D. K.; Peterson, E. D.; Holman, R. R.  Diabetes Care Apr 2017;40(4):494-501 | Ancillary Publication |
| #240 Persistent Effects of Intensive Glycemic Control on Retinopathy in Type 2 Diabetes in the Action to Control Cardiovascular Risk in Diabetes (ACCORD) Follow-On Study Diabetes Care Jul 2016;39(7):1089-100 | Ancillary Publication |
| Bonaca 2020 Dapagliflozin and Cardiac, Kidney, and Limb Outcomes in Patients With and Without Peripheral Artery Disease in DECLARE-TIMI 58 Bonaca, M. P.; Wiviott, S. D.; Zelniker, T. A.; Mosenzon, O.; Bhatt, D. L.; Leiter, L. A.; McGuire, D. K.; Goodrich, E. L.; De Mendonca Furtado, R. H.; Wilding, J. P. H.; Cahn, A.; Gause-Nilsson, I. A. M.; Johanson, P.; Fredriksson, M.; Johansson, P. A.; Langkilde, A. M.; Raz, I.; Sabatine, M. S.  Circulation Aug 25 2020;142(8):734-747 | Ancillary Publication |
| Verma 2018 Cardiovascular Outcomes and Safety of Empagliflozin in Patients With Type 2 Diabetes Mellitus and Peripheral Artery Disease: A Subanalysis of EMPA-REG OUTCOME Verma, S.; Mazer, C. D.; Al-Omran, M.; Inzucchi, S. E.; Fitchett, D.; Hehnke, U.; George, J. T.; Zinman, B.  Circulation Jan 23 2018;137(4):405-407 | Ancillary Publication |
| Holman 2008 10-year follow-up of intensive glucose control in type 2 diabetes Holman, R. R.; Paul, S. K.; Bethel, M. A.; Matthews, D. R.; Neil, H. A.  N Engl J Med Oct 9 2008;359(15):1577-89 | Ancillary Publication |
| Niskanen 2001 Reduced cardiovascular morbidity and mortality in hypertensive diabetic patients on first-line therapy with an ACE inhibitor compared with a diuretic/beta-blocker-based treatment regimen: a subanalysis of the Captopril Prevention Project Niskanen, L.; Hedner, T.; Hansson, L.; Lanke, J.; Niklason, A.  Diabetes Care Dec 2001;24(12):2091-6 | Ancillary Publication |
| Mahaffey 2018 Canagliflozin for Primary and Secondary Prevention of Cardiovascular Events: Results From the CANVAS Program (Canagliflozin Cardiovascular Assessment Study) Mahaffey, K. W.; Neal, B.; Perkovic, V.; de Zeeuw, D.; Fulcher, G.; Erondu, N.; Shaw, W.; Fabbrini, E.; Sun, T.; Li, Q.; Desai, M.; Matthews, D. R.  Circulation Jan 23 2018;137(4):323-334 | Ancillary Publication |
| Althouse 2013 Favorable effects of insulin sensitizers pertinent to peripheral arterial disease in type 2 diabetes: results from the Bypass Angioplasty Revascularization Investigation 2 Diabetes (BARI 2D) trial Althouse, A. D.; Abbott, J. D.; Sutton-Tyrrell, K.; Forker, A. D.; Lombardero, M. S.; Buitrón, L. V.; Pena-Sing, I.; Tardif, J. C.; Brooks, M. M.  Diabetes Care Oct 2013;36(10):3269-75 | Ancillary Publication |
| Gerstein 2019 Dulaglutide and renal outcomes in type 2 diabetes: an exploratory analysis of the REWIND randomised, placebo-controlled trial Gerstein, H. C.; Colhoun, H. M.; Dagenais, G. R.; Diaz, R.; Lakshmanan, M.; Pais, P.; Probstfield, J.; Botros, F. T.; Riddle, M. C.; Rydén, L.; Xavier, D.; Atisso, C. M.; Dyal, L.; Hall, S.; Rao-Melacini, P.; Wong, G.; Avezum, A.; Basile, J.; Chung, N.; Conget, I.; Cushman, W. C.; Franek, E.; Hancu, N.; Hanefeld, M.; Holt, S.; Jansky, P.; Keltai, M.; Lanas, F.; Leiter, L. A.; Lopez-Jaramillo, P.; Cardona Munoz, E. G.; Pirags, V.; Pogosova, N.; Raubenheimer, P. J.; Shaw, J. E.; Sheu, W. H.; Temelkova-Kurktschiev, T.  Lancet Jul 13 2019;394(10193):131-138 | Ancillary Publication |
| Keech 2007 Effect of fenofibrate on the need for laser treatment for diabetic retinopathy (FIELD study): a randomised controlled trial Keech, A. C.; Mitchell, P.; Summanen, P. A.; O'Day, J.; Davis, T. M.; Moffitt, M. S.; Taskinen, M. R.; Simes, R. J.; Tse, D.; Williamson, E.; Merrifield, A.; Laatikainen, L. T.; d'Emden, M. C.; Crimet, D. C.; O'Connell, R. L.; Colman, P. G.  Lancet Nov 17 2007;370(9600):1687-97 | Ancillary Publication |
| Vakkilainen 2003 Relationships between low-density lipoprotein particle size, plasma lipoproteins, and progression of coronary artery disease: the Diabetes Atherosclerosis Intervention Study (DAIS) Vakkilainen, J.; Steiner, G.; Ansquer, J. C.; Aubin, F.; Rattier, S.; Foucher, C.; Hamsten, A.; Taskinen, M. R.  Circulation Apr 8 2003;107(13):1733-7 | Ancillary Publication |
| Bethel 2020 Microvascular and Cardiovascular Outcomes According to Renal Function in Patients Treated With Once-Weekly Exenatide: Insights From the EXSCEL Trial Bethel, M. A.; Mentz, R. J.; Merrill, P.; Buse, J. B.; Chan, J. C.; Goodman, S. G.; Iqbal, N.; Jakuboniene, N.; Katona, B.; Lokhnygina, Y.; Lopes, R. D.; Maggioni, A. P.; Ohman, P.; Tankova, T.; Bakris, G. L.; Hernandez, A. F.; Holman, R. R.  Diabetes Care Feb 2020;43(2):446-452 | Ancillary Publication |
| Ginsberg 2010 Effects of combination lipid therapy in type 2 diabetes mellitus Ginsberg, H. N.; Elam, M. B.; Lovato, L. C.; Crouse, J. R., 3rd; Leiter, L. A.; Linz, P.; Friedewald, W. T.; Buse, J. B.; Gerstein, H. C.; Probstfield, J.; Grimm, R. H.; Ismail-Beigi, F.; Bigger, J. T.; Goff, D. C., Jr.; Cushman, W. C.; Simons-Morton, D. G.; Byington, R. P.  N Engl J Med Apr 29 2010;362(17):1563-74 | Ancillary Publication |
| Bethel 2020 Exploring the Possible Impact of Unbalanced Open-Label Drop-In of Glucose-Lowering Medications on EXSCEL Outcomes Bethel, M. A.; Stevens, S. R.; Buse, J. B.; Choi, J.; Gustavson, S. M.; Iqbal, N.; Lokhnygina, Y.; Mentz, R. J.; Patel, R. A.; Öhman, P.; Schernthaner, G.; Lecube, A.; Hernandez, A. F.; Holman, R. R.  Circulation Apr 28 2020;141(17):1360-1370 | Ancillary Publication |
| Rajamani 2009 Effect of fenofibrate on amputation events in people with type 2 diabetes mellitus (FIELD study): a prespecified analysis of a randomised controlled trial Rajamani, K.; Colman, P. G.; Li, L. P.; Best, J. D.; Voysey, M.; D'Emden, M. C.; Laakso, M.; Baker, J. R.; Keech, A. C.  Lancet May 23 2009;373(9677):1780-8 | Ancillary Publication |
| Schwartz 2012 Intensive glycemic control is not associated with fractures or falls in the ACCORD randomized trial Schwartz, A. V.; Margolis, K. L.; Sellmeyer, D. E.; Vittinghoff, E.; Ambrosius, W. T.; Bonds, D. E.; Josse, R. G.; Schnall, A. M.; Simmons, D. L.; Hue, T. F.; Palermo, L.; Hamilton, B. P.; Green, J. B.; Atkinson, H. H.; O'Connor, P. J.; Force, R. W.; Bauer, D. C.  Diabetes Care Jul 2012;35(7):1525-31 | Ancillary Publication |
| Fitchett 2019 Empagliflozin Reduced Mortality and Hospitalization for Heart Failure Across the Spectrum of Cardiovascular Risk in the EMPA-REG OUTCOME Trial Fitchett, D.; Inzucchi, S. E.; Cannon, C. P.; McGuire, D. K.; Scirica, B. M.; Johansen, O. E.; Sambevski, S.; Kaspers, S.; Pfarr, E.; George, J. T.; Zinman, B.  Circulation Mar 12 2019;139(11):1384-1395 | Ancillary Publication |
| Garvey 2020 Efficacy and Safety of Liraglutide 3.0 mg in Individuals With Overweight or Obesity and Type 2 Diabetes Treated With Basal Insulin: The SCALE Insulin Randomized Controlled Trial Garvey, W. T.; Birkenfeld, A. L.; Dicker, D.; Mingrone, G.; Pedersen, S. D.; Satylganova, A.; Skovgaard, D.; Sugimoto, D.; Jensen, C.; Mosenzon, O.  Diabetes Care May 2020;43(5):1085-1093 | Ancillary Publication |
| Wanner 2018 Empagliflozin and Clinical Outcomes in Patients With Type 2 Diabetes Mellitus, Established Cardiovascular Disease, and Chronic Kidney Disease Wanner, C.; Lachin, J. M.; Inzucchi, S. E.; Fitchett, D.; Mattheus, M.; George, J.; Woerle, H. J.; Broedl, U. C.; von Eynatten, M.; Zinman, B.  Circulation Jan 9 2018;137(2):119-129 | Ancillary Publication |
| Reed 2020 Within-Trial Evaluation of Medical Resources, Costs, and Quality of Life Among Patients With Type 2 Diabetes Participating in the Exenatide Study of Cardiovascular Event Lowering (EXSCEL) Reed, S. D.; Li, Y.; Dakin, H. A.; Becker, F.; Leal, J.; Gustavson, S. M.; Kartman, B.; Wittbrodt, E.; Mentz, R. J.; Pagidipati, N. J.; Bethel, M. A.; Gray, A. M.; Holman, R. R.; Hernandez, A. F.  Diabetes Care Feb 2020;43(2):374-381 | Ancillary Publication |
| Cahn 2016 Predisposing Factors for Any and Major Hypoglycemia With Saxagliptin Versus Placebo and Overall: Analysis From the SAVOR-TIMI 53 Trial Cahn, A.; Raz, I.; Mosenzon, O.; Leibowitz, G.; Yanuv, I.; Rozenberg, A.; Iqbal, N.; Hirshberg, B.; Sjostrand, M.; Stahre, C.; Im, K.; Kanevsky, E.; Scirica, B. M.; Bhatt, D. L.; Braunwald, E.  Diabetes Care Aug 2016;39(8):1329-37 | Ancillary Publication |
| Mann 2017 Liraglutide and Renal Outcomes in Type 2 Diabetes Mann, J. F. E.; Ørsted, D. D.; Brown-Frandsen, K.; Marso, S. P.; Poulter, N. R.; Rasmussen, S.; Tornøe, K.; Zinman, B.; Buse, J. B.  N Engl J Med Aug 31 2017;377(9):839-848 | Ancillary Publication |
| Linz 2014 Paradoxical reduction in HDL-C with fenofibrate and thiazolidinedione therapy in type 2 diabetes: the ACCORD Lipid Trial Linz, P. E.; Lovato, L. C.; Byington, R. P.; O'Connor, P. J.; Leiter, L. A.; Weiss, D.; Force, R. W.; Crouse, J. R.; Ismail-Beigi, F.; Simmons, D. L.; Papademetriou, V.; Ginsberg, H. N.; Elam, M. B.  Diabetes Care 2014;37(3):686-93 | Ancillary Publication |
| Mann 2018 Effects of Liraglutide Versus Placebo on Cardiovascular Events in Patients With Type 2 Diabetes Mellitus and Chronic Kidney Disease Mann, J. F. E.; Fonseca, V.; Mosenzon, O.; Raz, I.; Goldman, B.; Idorn, T.; von Scholten, B. J.; Poulter, N. R.  Circulation Dec 18 2018;138(25):2908-2918 | Ancillary Publication |
| Anderson 2014 Blood pressure and pulse pressure effects on renal outcomes in the Veterans Affairs Diabetes Trial (VADT) Anderson, R. J.; Bahn, G. D.; Emanuele, N. V.; Marks, J. B.; Duckworth, W. C.  Diabetes Care Oct 2014;37(10):2782-8 | Ancillary Publication |
| Rossing 2003 Comparative effects of Irbesartan on ambulatory and office blood pressure: a substudy of ambulatory blood pressure from the Irbesartan in Patients with Type 2 Diabetes and Microalbuminuria study Rossing, K.; Christensen, P. K.; Andersen, S.; Hovind, P.; Hansen, H. P.; Parving, H. H.  Diabetes Care Mar 2003;26(3):569-74 | Ancillary Publication |
| Holman 2009 Three-year efficacy of complex insulin regimens in type 2 diabetes Holman, R. R.; Farmer, A. J.; Davies, M. J.; Levy, J. C.; Darbyshire, J. L.; Keenan, J. F.; Paul, S. K.  N Engl J Med Oct 29 2009;361(18):1736-47 | Ancillary Publication |
| Patel 2008 Intensive blood glucose control and vascular outcomes in patients with type 2 diabetes Patel, A.; MacMahon, S.; Chalmers, J.; Neal, B.; Billot, L.; Woodward, M.; Marre, M.; Cooper, M.; Glasziou, P.; Grobbee, D.; Hamet, P.; Harrap, S.; Heller, S.; Liu, L.; Mancia, G.; Mogensen, C. E.; Pan, C.; Poulter, N.; Rodgers, A.; Williams, B.; Bompoint, S.; de Galan, B. E.; Joshi, R.; Travert, F.  N Engl J Med Jun 12 2008;358(24):2560-72 | Ancillary Publication |
| Chew 2010 Effects of medical therapies on retinopathy progression in type 2 diabetes Chew, E. Y.; Ambrosius, W. T.; Davis, M. D.; Danis, R. P.; Gangaputra, S.; Greven, C. M.; Hubbard, L.; Esser, B. A.; Lovato, J. F.; Perdue, L. H.; Goff, D. C., Jr.; Cushman, W. C.; Ginsberg, H. N.; Elam, M. B.; Genuth, S.; Gerstein, H. C.; Schubart, U.; Fine, L. J.  N Engl J Med Jul 15 2010;363(3):233-44 | Ancillary Publication |
| Zoungas 2009 Combined effects of routine blood pressure lowering and intensive glucose control on macrovascular and microvascular outcomes in patients with type 2 diabetes: New results from the ADVANCE trial Zoungas, S.; de Galan, B. E.; Ninomiya, T.; Grobbee, D.; Hamet, P.; Heller, S.; MacMahon, S.; Marre, M.; Neal, B.; Patel, A.; Woodward, M.; Chalmers, J.; Cass, A.; Glasziou, P.; Harrap, S.; Lisheng, L.; Mancia, G.; Pillai, A.; Poulter, N.; Perkovic, V.; Travert, F.  Diabetes Care Nov 2009;32(11):2068-74 | Ancillary Publication |
| Hayward 2015 Follow-up of glycemic control and cardiovascular outcomes in type 2 diabetes Hayward, R. A.; Reaven, P. D.; Wiitala, W. L.; Bahn, G. D.; Reda, D. J.; Ge, L.; McCarren, M.; Duckworth, W. C.; Emanuele, N. V.  N Engl J Med Jun 4 2015;372(23):2197-206 | Ancillary Publication |
| Zoungas 2014 Follow-up of blood-pressure lowering and glucose control in type 2 diabetes Zoungas, S.; Chalmers, J.; Neal, B.; Billot, L.; Li, Q.; Hirakawa, Y.; Arima, H.; Monaghan, H.; Joshi, R.; Colagiuri, S.; Cooper, M. E.; Glasziou, P.; Grobbee, D.; Hamet, P.; Harrap, S.; Heller, S.; Lisheng, L.; Mancia, G.; Marre, M.; Matthews, D. R.; Mogensen, C. E.; Perkovic, V.; Poulter, N.; Rodgers, A.; Williams, B.; MacMahon, S.; Patel, A.; Woodward, M.  N Engl J Med Oct 9 2014;371(15):1392-406 | Ancillary Publication |
| Ahrén 2005 Improved meal-related beta-cell function and insulin sensitivity by the dipeptidyl peptidase-IV inhibitor vildagliptin in metformin-treated patients with type 2 diabetes over 1 year Ahrén, B.; Pacini, G.; Foley, J. E.; Schweizer, A.  Diabetes Care Aug 2005;28(8):1936-40 | Ancillary Publication |
| Home 2009 Rosiglitazone evaluated for cardiovascular outcomes in oral agent combination therapy for type 2 diabetes (RECORD): a multicentre, randomised, open-label trial Home, P. D.; Pocock, S. J.; Beck-Nielsen, H.; Curtis, P. S.; Gomis, R.; Hanefeld, M.; Jones, N. P.; Komajda, M.; McMurray, J. J.  Lancet Jun 20 2009;373(9681):2125-35 | Ancillary Publication |
| Woodward 2011 Does glycemic control offer similar benefits among patients with diabetes in different regions of the world? Results from the ADVANCE trial Woodward, M.; Patel, A.; Zoungas, S.; Liu, L.; Pan, C.; Poulter, N.; Januszewicz, A.; Tandon, N.; Joshi, P.; Heller, S.; Neal, B.; Chalmers, J.  Diabetes Care Dec 2011;34(12):2491-5 | Ancillary Publication |
| Sandbæk 2014 Effect of early multifactorial therapy compared with routine care on microvascular outcomes at 5 years in people with screen-detected diabetes: a randomized controlled trial: the ADDITION-Europe Study Sandbæk, A.; Griffin, S. J.; Sharp, S. J.; Simmons, R. K.; Borch-Johnsen, K.; Rutten, G. E.; van den Donk, M.; Wareham, N. J.; Lauritzen, T.; Davies, M. J.; Khunti, K.  Diabetes Care Jul 2014;37(7):2015-23 | Ancillary Publication |
| Gaede 2008 Effect of a multifactorial intervention on mortality in type 2 diabetes Gaede, P.; Lund-Andersen, H.; Parving, H. H.; Pedersen, O.  N Engl J Med Feb 7 2008;358(6):580-91 | Ancillary Publication |
| Barter 2011 Effect of torcetrapib on glucose, insulin, and hemoglobin A1c in subjects in the Investigation of Lipid Level Management to Understand its Impact in Atherosclerotic Events (ILLUMINATE) trial Barter, P. J.; Rye, K. A.; Tardif, J. C.; Waters, D. D.; Boekholdt, S. M.; Breazna, A.; Kastelein, J. J.  Circulation Aug 2 2011;124(5):555-62 | Ancillary Publication |
| Ceriello 2005 Comparison of effect of pioglitazone with metformin or sulfonylurea (monotherapy and combination therapy) on postload glycemia and composite insulin sensitivity index during an oral glucose tolerance test in patients with type 2 diabetes Ceriello, A.; Johns, D.; Widel, M.; Eckland, D. J.; Gilmore, K. J.; Tan, M. H.  Diabetes Care Feb 2005;28(2):266-72 | Ancillary Publication |
| Cushman 2010 Effects of intensive blood-pressure control in type 2 diabetes mellitus Cushman, W. C.; Evans, G. W.; Byington, R. P.; Goff, D. C., Jr.; Grimm, R. H., Jr.; Cutler, J. A.; Simons-Morton, D. G.; Basile, J. N.; Corson, M. A.; Probstfield, J. L.; Katz, L.; Peterson, K. A.; Friedewald, W. T.; Buse, J. B.; Bigger, J. T.; Gerstein, H. C.; Ismail-Beigi, F.  N Engl J Med Apr 29 2010;362(17):1575-85 | Ancillary Publication |
| Buse 2010 Switching to once-daily liraglutide from twice-daily exenatide further improves glycemic control in patients with type 2 diabetes using oral agents Buse, J. B.; Sesti, G.; Schmidt, W. E.; Montanya, E.; Chang, C. T.; Xu, Y.; Blonde, L.; Rosenstock, J.  Diabetes Care Jun 2010;33(6):1300-3 | Ancillary Publication |
| Zoungas 2010 Severe hypoglycemia and risks of vascular events and death Zoungas, S.; Patel, A.; Chalmers, J.; de Galan, B. E.; Li, Q.; Billot, L.; Woodward, M.; Ninomiya, T.; Neal, B.; MacMahon, S.; Grobbee, D. E.; Kengne, A. P.; Marre, M.; Heller, S.  N Engl J Med Oct 7 2010;363(15):1410-8 | Ancillary Publication |
| Tian 2020 Effects of Intensive Glycemic Control on Clinical Outcomes Among Patients With Type 2 Diabetes With Different Levels of Cardiovascular Risk and Hemoglobin A(1c) in the ADVANCE Trial Tian, J.; Ohkuma, T.; Cooper, M.; Harrap, S.; Mancia, G.; Poulter, N.; Wang, J. G.; Zoungas, S.; Woodward, M.; Chalmers, J.  Diabetes Care Jun 2020;43(6):1293-1299 | Ancillary Publication |
| Hata 2013 Effects of visit-to-visit variability in systolic blood pressure on macrovascular and microvascular complications in patients with type 2 diabetes mellitus: the ADVANCE trial Hata, J.; Arima, H.; Rothwell, P. M.; Woodward, M.; Zoungas, S.; Anderson, C.; Patel, A.; Neal, B.; Glasziou, P.; Hamet, P.; Mancia, G.; Poulter, N.; Williams, B.; Macmahon, S.; Chalmers, J.  Circulation Sep 17 2013;128(12):1325-34 | Ancillary Publication |
| Solomon 2010 Erythropoietic response and outcomes in kidney disease and type 2 diabetes Solomon, S. D.; Uno, H.; Lewis, E. F.; Eckardt, K. U.; Lin, J.; Burdmann, E. A.; de Zeeuw, D.; Ivanovich, P.; Levey, A. S.; Parfrey, P.; Remuzzi, G.; Singh, A. K.; Toto, R.; Huang, F.; Rossert, J.; McMurray, J. J.; Pfeffer, M. A.  N Engl J Med Sep 16 2010;363(12):1146-55 | Ancillary Publication |
| Barzilay 2014 The impact of salsalate treatment on serum levels of advanced glycation end products in type 2 diabetes Barzilay, J. I.; Jablonski, K. A.; Fonseca, V.; Shoelson, S. E.; Goldfine, A. B.; Strauch, C.; Monnier, V. M.  Diabetes Care Apr 2014;37(4):1083-91 | Ancillary Publication |
| Industry Trial in a journal with IF< 30 | 263 |
| Rosenstock 2016 Initial Combination Therapy With Canagliflozin Plus Metformin Versus Each Component as Monotherapy for Drug-Naïve Type 2 Diabetes Rosenstock, J.; Chuck, L.; González-Ortiz, M.; Merton, K.; Craig, J.; Capuano, G.; Qiu, R.  Diabetes Care Mar 2016;39(3):353-62 | Industry Trial in a journal with IF< 30 |
| Viberti 2002 A diabetes outcome progression trial (ADOPT): an international multicenter study of the comparative efficacy of rosiglitazone, glyburide, and metformin in recently diagnosed type 2 diabetes Viberti, G.; Kahn, S. E.; Greene, D. A.; Herman, W. H.; Zinman, B.; Holman, R. R.; Haffner, S. M.; Levy, D.; Lachin, J. M.; Berry, R. A.; Heise, M. A.; Jones, N. P.; Freed, M. I.  Diabetes Care Oct 2002;25(10):1737-43 | Industry Trial in a journal with IF< 30 |
| Gough 2013 Low-volume insulin degludec 200 units/ml once daily improves glycemic control similarly to insulin glargine with a low risk of hypoglycemia in insulin-naive patients with type 2 diabetes: a 26-week, randomized, controlled, multinational, treat-to-target trial: the BEGIN LOW VOLUME trial Gough, S. C.; Bhargava, A.; Jain, R.; Mersebach, H.; Rasmussen, S.; Bergenstal, R. M.  Diabetes Care Sep 2013;36(9):2536-42 | Industry Trial in a journal with IF< 30 |
| Hermansen 2004 Intensive therapy with inhaled insulin via the AERx insulin diabetes management system: a 12-week proof-of-concept trial in patients with type 2 diabetes Hermansen, K.; Rönnemaa, T.; Petersen, A. H.; Bellaire, S.; Adamson, U.  Diabetes Care Jan 2004;27(1):162-7 | Industry Trial in a journal with IF< 30 |
| Ziegler 2009 Treatment of symptomatic polyneuropathy with actovegin in type 2 diabetic patients Ziegler, D.; Movsesyan, L.; Mankovsky, B.; Gurieva, I.; Abylaiuly, Z.; Strokov, I.  Diabetes Care Aug 2009;32(8):1479-84 | Industry Trial in a journal with IF< 30 |
| Buse 2016 Randomized Clinical Trial Comparing Basal Insulin Peglispro and Insulin Glargine in Patients With Type 2 Diabetes Previously Treated With Basal Insulin: IMAGINE 5 Buse, J. B.; Rodbard, H. W.; Trescoli Serrano, C.; Luo, J.; Ivanyi, T.; Bue-Valleskey, J.; Hartman, M. L.; Carey, M. A.; Chang, A. M.  Diabetes Care Jan 2016;39(1):92-100 | Industry Trial in a journal with IF< 30 |
| Leiter 2014 Efficacy and safety of the once-weekly GLP-1 receptor agonist albiglutide versus sitagliptin in patients with type 2 diabetes and renal impairment: a randomized phase III study Leiter, L. A.; Carr, M. C.; Stewart, M.; Jones-Leone, A.; Scott, R.; Yang, F.; Handelsman, Y.  Diabetes Care Oct 2014;37(10):2723-30 | Industry Trial in a journal with IF< 30 |
| Buse 2011 The DURAbility of Basal versus Lispro mix 75/25 insulin Efficacy (DURABLE) trial: comparing the durability of lispro mix 75/25 and glargine Buse, J. B.; Wolffenbuttel, B. H.; Herman, W. H.; Hippler, S.; Martin, S. A.; Jiang, H. H.; Shenouda, S. K.; Fahrbach, J. L.  Diabetes Care Feb 2011;34(2):249-55 | Industry Trial in a journal with IF< 30 |
| Mathieu 2015 Randomized, Double-Blind, Phase 3 Trial of Triple Therapy With Dapagliflozin Add-on to Saxagliptin Plus Metformin in Type 2 Diabetes Mathieu, C.; Ranetti, A. E.; Li, D.; Ekholm, E.; Cook, W.; Hirshberg, B.; Chen, H.; Hansen, L.; Iqbal, N.  Diabetes Care Nov 2015;38(11):2009-17 | Industry Trial in a journal with IF< 30 |
| Rosenstock 2014 Improved glucose control with weight loss, lower insulin doses, and no increased hypoglycemia with empagliflozin added to titrated multiple daily injections of insulin in obese inadequately controlled type 2 diabetes Rosenstock, J.; Jelaska, A.; Frappin, G.; Salsali, A.; Kim, G.; Woerle, H. J.; Broedl, U. C.  Diabetes Care Jul 2014;37(7):1815-23 | Industry Trial in a journal with IF< 30 |
| DeVries 2012 Sequential intensification of metformin treatment in type 2 diabetes with liraglutide followed by randomized addition of basal insulin prompted by A1C targets DeVries, J. H.; Bain, S. C.; Rodbard, H. W.; Seufert, J.; D'Alessio, D.; Thomsen, A. B.; Zychma, M.; Rosenstock, J.  Diabetes Care Jul 2012;35(7):1446-54 | Industry Trial in a journal with IF< 30 |
| DeFronzo 2015 Combination of empagliflozin and linagliptin as second-line therapy in subjects with type 2 diabetes inadequately controlled on metformin DeFronzo, R. A.; Lewin, A.; Patel, S.; Liu, D.; Kaste, R.; Woerle, H. J.; Broedl, U. C.  Diabetes Care Mar 2015;38(3):384-93 | Industry Trial in a journal with IF< 30 |
| Nauck 2009 Treatment with the human once-weekly glucagon-like peptide-1 analog taspoglutide in combination with metformin improves glycemic control and lowers body weight in patients with type 2 diabetes inadequately controlled with metformin alone: a double-blind placebo-controlled study Nauck, M. A.; Ratner, R. E.; Kapitza, C.; Berria, R.; Boldrin, M.; Balena, R.  Diabetes Care Jul 2009;32(7):1237-43 | Industry Trial in a journal with IF< 30 |
| DePaoli 2014 Can a selective PPARγ modulator improve glycemic control in patients with type 2 diabetes with fewer side effects compared with pioglitazone? DePaoli, A. M.; Higgins, L. S.; Henry, R. R.; Mantzoros, C.; Dunn, F. L.  Diabetes Care Jul 2014;37(7):1918-23 | Industry Trial in a journal with IF< 30 |
| Buse 2011 Use of twice-daily exenatide in Basal insulin-treated patients with type 2 diabetes: a randomized, controlled trial Buse, J. B.; Bergenstal, R. M.; Glass, L. C.; Heilmann, C. R.; Lewis, M. S.; Kwan, A. Y.; Hoogwerf, B. J.; Rosenstock, J.  Ann Intern Med Jan 18 2011;154(2):103-12 | Industry Trial in a journal with IF< 30 |
| Franchi 2020 Pharmacodynamic and Pharmacokinetic Effects of a Low Maintenance Dose Ticagrelor Regimen Versus Standard Dose Clopidogrel in Diabetes Mellitus Patients Without Previous Major Cardiovascular Events Undergoing Elective Percutaneous Coronary Intervention: The OPTIMUS-6 Study Franchi, F.; Rollini, F.; Been, L.; Briceno, M.; Maaliki, N.; Wali, M.; Rivas, A.; Pineda, A. M.; Suryadevara, S.; Soffer, D.; Zenni, M. M.; Bass, T. A.; Angiolillo, D. J.  Circulation Oct 13 2020;142(15):1500-1502 | Industry Trial in a journal with IF< 30 |
| Raskin 2003 Efficacy and safety of combination therapy: repaglinide plus metformin versus nateglinide plus metformin Raskin, P.; Klaff, L.; McGill, J.; South, S. A.; Hollander, P.; Khutoryansky, N.; Hale, P. M.  Diabetes Care Jul 2003;26(7):2063-8 | Industry Trial in a journal with IF< 30 |
| Fonseca 2008 Colesevelam HCl improves glycemic control and reduces LDL cholesterol in patients with inadequately controlled type 2 diabetes on sulfonylurea-based therapy Fonseca, V. A.; Rosenstock, J.; Wang, A. C.; Truitt, K. E.; Jones, M. R.  Diabetes Care Aug 2008;31(8):1479-84 | Industry Trial in a journal with IF< 30 |
| Wilding 2012 Long-term efficacy of dapagliflozin in patients with type 2 diabetes mellitus receiving high doses of insulin: a randomized trial Wilding, J. P.; Woo, V.; Soler, N. G.; Pahor, A.; Sugg, J.; Rohwedder, K.; Parikh, S.  Ann Intern Med Mar 20 2012;156(6):405-15 | Industry Trial in a journal with IF< 30 |
| Miles 2002 Effect of orlistat in overweight and obese patients with type 2 diabetes treated with metformin Miles, J. M.; Leiter, L.; Hollander, P.; Wadden, T.; Anderson, J. W.; Doyle, M.; Foreyt, J.; Aronne, L.; Klein, S.  Diabetes Care Jul 2002;25(7):1123-8 | Industry Trial in a journal with IF< 30 |
| Swinnen 2010 A 24-week, randomized, treat-to-target trial comparing initiation of insulin glargine once-daily with insulin detemir twice-daily in patients with type 2 diabetes inadequately controlled on oral glucose-lowering drugs Swinnen, S. G.; Dain, M. P.; Aronson, R.; Davies, M.; Gerstein, H. C.; Pfeiffer, A. F.; Snoek, F. J.; Devries, J. H.; Hoekstra, J. B.; Holleman, F.  Diabetes Care Jun 2010;33(6):1176-8 | Industry Trial in a journal with IF< 30 |
| Chan 2004 Renin angiotensin aldosterone system blockade and renal disease in patients with type 2 diabetes. An Asian perspective from the RENAAL Study Chan, J. C.; Wat, N. M.; So, W. Y.; Lam, K. S.; Chua, C. T.; Wong, K. S.; Morad, Z.; Dickson, T. Z.; Hille, D.; Zhang, Z.; Cooper, M. E.; Shahinfar, S.; Brenner, B. M.; Kurokawa, K.  Diabetes Care Apr 2004;27(4):874-9 | Industry Trial in a journal with IF< 30 |
| Russell-Jones 2012 Efficacy and safety of exenatide once weekly versus metformin, pioglitazone, and sitagliptin used as monotherapy in drug-naive patients with type 2 diabetes (DURATION-4): a 26-week double-blind study Russell-Jones, D.; Cuddihy, R. M.; Hanefeld, M.; Kumar, A.; González, J. G.; Chan, M.; Wolka, A. M.; Boardman, M. K.  Diabetes Care Feb 2012;35(2):252-8 | Industry Trial in a journal with IF< 30 |
| Bowering 2017 Faster Aspart Versus Insulin Aspart as Part of a Basal-Bolus Regimen in Inadequately Controlled Type 2 Diabetes: The onset 2 Trial Bowering, K.; Case, C.; Harvey, J.; Reeves, M.; Sampson, M.; Strzinek, R.; Bretler, D. M.; Bang, R. B.; Bode, B. W.  Diabetes Care Jul 2017;40(7):951-957 | Industry Trial in a journal with IF< 30 |
| Diamant 2014 Glucagon-like peptide 1 receptor agonist or bolus insulin with optimized basal insulin in type 2 diabetes Diamant, M.; Nauck, M. A.; Shaginian, R.; Malone, J. K.; Cleall, S.; Reaney, M.; de Vries, D.; Hoogwerf, B. J.; MacConell, L.; Wolffenbuttel, B. H.  Diabetes Care Oct 2014;37(10):2763-73 | Industry Trial in a journal with IF< 30 |
| Ferrannini 2010 Dapagliflozin monotherapy in type 2 diabetic patients with inadequate glycemic control by diet and exercise: a randomized, double-blind, placebo-controlled, phase 3 trial Ferrannini, E.; Ramos, S. J.; Salsali, A.; Tang, W.; List, J. F.  Diabetes Care Oct 2010;33(10):2217-24 | Industry Trial in a journal with IF< 30 |
| Charbonnel 2004 The prospective pioglitazone clinical trial in macrovascular events (PROactive): can pioglitazone reduce cardiovascular events in diabetes? Study design and baseline characteristics of 5238 patients Charbonnel, B.; Dormandy, J.; Erdmann, E.; Massi-Benedetti, M.; Skene, A.  Diabetes Care Jul 2004;27(7):1647-53 | Industry Trial in a journal with IF< 30 |
| Rosenstock 2019 Once-Weekly Efpeglenatide Dose-Range Effects on Glycemic Control and Body Weight in Patients With Type 2 Diabetes on Metformin or Drug Naive, Referenced to Liraglutide Rosenstock, J.; Sorli, C. H.; Trautmann, M. E.; Morales, C.; Wendisch, U.; Dailey, G.; Hompesch, M.; Choi, I. Y.; Kang, J.; Stewart, J.; Yoon, K. H.  Diabetes Care Sep 2019;42(9):1733-1741 | Industry Trial in a journal with IF< 30 |
| Edelman 2014 AUTONOMY: the first randomized trial comparing two patient-driven approaches to initiate and titrate prandial insulin lispro in type 2 diabetes Edelman, S. V.; Liu, R.; Johnson, J.; Glass, L. C.  Diabetes Care Aug 2014;37(8):2132-40 | Industry Trial in a journal with IF< 30 |
| Nauck 2016 A Phase 2, Randomized, Dose-Finding Study of the Novel Once-Weekly Human GLP-1 Analog, Semaglutide, Compared With Placebo and Open-Label Liraglutide in Patients With Type 2 Diabetes Nauck, M. A.; Petrie, J. R.; Sesti, G.; Mannucci, E.; Courrèges, J. P.; Lindegaard, M. L.; Jensen, C. B.; Atkin, S. L.  Diabetes Care Feb 2016;39(2):231-41 | Industry Trial in a journal with IF< 30 |
| Rosenstock 2007 Comparison of vildagliptin and rosiglitazone monotherapy in patients with type 2 diabetes: a 24-week, double-blind, randomized trial Rosenstock, J.; Baron, M. A.; Dejager, S.; Mills, D.; Schweizer, A.  Diabetes Care Feb 2007;30(2):217-23 | Industry Trial in a journal with IF< 30 |
| Hollander 2010 Effect of rimonabant on glycemic control in insulin-treated type 2 diabetes: the ARPEGGIO trial Hollander, P. A.; Amod, A.; Litwak, L. E.; Chaudhari, U.  Diabetes Care Mar 2010;33(3):605-7 | Industry Trial in a journal with IF< 30 |
| Meininger 2011 Effects of MK-0941, a novel glucokinase activator, on glycemic control in insulin-treated patients with type 2 diabetes Meininger, G. E.; Scott, R.; Alba, M.; Shentu, Y.; Luo, E.; Amin, H.; Davies, M. J.; Kaufman, K. D.; Goldstein, B. J.  Diabetes Care Dec 2011;34(12):2560-6 | Industry Trial in a journal with IF< 30 |
| Buse 2010 DURATION-1: exenatide once weekly produces sustained glycemic control and weight loss over 52 weeks Buse, J. B.; Drucker, D. J.; Taylor, K. L.; Kim, T.; Walsh, B.; Hu, H.; Wilhelm, K.; Trautmann, M.; Shen, L. Z.; Porter, L. E.  Diabetes Care Jun 2010;33(6):1255-61 | Industry Trial in a journal with IF< 30 |
| Rosenstock 2013 The fate of taspoglutide, a weekly GLP-1 receptor agonist, versus twice-daily exenatide for type 2 diabetes: the T-emerge 2 trial Rosenstock, J.; Balas, B.; Charbonnel, B.; Bolli, G. B.; Boldrin, M.; Ratner, R.; Balena, R.  Diabetes Care Mar 2013;36(3):498-504 | Industry Trial in a journal with IF< 30 |
| Zinman 2007 The effect of adding exenatide to a thiazolidinedione in suboptimally controlled type 2 diabetes: a randomized trial Zinman, B.; Hoogwerf, B. J.; Durán García, S.; Milton, D. R.; Giaconia, J. M.; Kim, D. D.; Trautmann, M. E.; Brodows, R. G.  Ann Intern Med Apr 3 2007;146(7):477-85 | Industry Trial in a journal with IF< 30 |
| Ferrannini 2013 Long-term safety and efficacy of empagliflozin, sitagliptin, and metformin: an active-controlled, parallel-group, randomized, 78-week open-label extension study in patients with type 2 diabetes Ferrannini, E.; Berk, A.; Hantel, S.; Pinnetti, S.; Hach, T.; Woerle, H. J.; Broedl, U. C.  Diabetes Care Dec 2013;36(12):4015-21 | Industry Trial in a journal with IF< 30 |
| Deeg 2007 Pioglitazone and rosiglitazone have different effects on serum lipoprotein particle concentrations and sizes in patients with type 2 diabetes and dyslipidemia Deeg, M. A.; Buse, J. B.; Goldberg, R. B.; Kendall, D. M.; Zagar, A. J.; Jacober, S. J.; Khan, M. A.; Perez, A. T.; Tan, M. H.  Diabetes Care Oct 2007;30(10):2458-64 | Industry Trial in a journal with IF< 30 |
| Meier 2015 Contrasting Effects of Lixisenatide and Liraglutide on Postprandial Glycemic Control, Gastric Emptying, and Safety Parameters in Patients With Type 2 Diabetes on Optimized Insulin Glargine With or Without Metformin: A Randomized, Open-Label Trial Meier, J. J.; Rosenstock, J.; Hincelin-Méry, A.; Roy-Duval, C.; Delfolie, A.; Coester, H. V.; Menge, B. A.; Forst, T.; Kapitza, C.  Diabetes Care Jul 2015;38(7):1263-73 | Industry Trial in a journal with IF< 30 |
| Vilsbøll 2007 Liraglutide, a long-acting human glucagon-like peptide-1 analog, given as monotherapy significantly improves glycemic control and lowers body weight without risk of hypoglycemia in patients with type 2 diabetes Vilsbøll, T.; Zdravkovic, M.; Le-Thi, T.; Krarup, T.; Schmitz, O.; Courrèges, J. P.; Verhoeven, R.; Bugánová, I.; Madsbad, S.  Diabetes Care Jun 2007;30(6):1608-10 | Industry Trial in a journal with IF< 30 |
| Goldstein 2007 Effect of initial combination therapy with sitagliptin, a dipeptidyl peptidase-4 inhibitor, and metformin on glycemic control in patients with type 2 diabetes Goldstein, B. J.; Feinglos, M. N.; Lunceford, J. K.; Johnson, J.; Williams-Herman, D. E.  Diabetes Care Aug 2007;30(8):1979-87 | Industry Trial in a journal with IF< 30 |
| Yki-Järvinen 2000 Less nocturnal hypoglycemia and better post-dinner glucose control with bedtime insulin glargine compared with bedtime NPH insulin during insulin combination therapy in type 2 diabetes. HOE 901/3002 Study Group Yki-Järvinen, H.; Dressler, A.; Ziemen, M.  Diabetes Care Aug 2000;23(8):1130-6 | Industry Trial in a journal with IF< 30 |
| Barnett 2006 An open, randomized, parallel-group study to compare the efficacy and safety profile of inhaled human insulin (Exubera) with glibenclamide as adjunctive therapy in patients with type 2 diabetes poorly controlled on metformin Barnett, A. H.; Dreyer, M.; Lange, P.; Serdarevic-Pehar, M.  Diabetes Care Aug 2006;29(8):1818-25 | Industry Trial in a journal with IF< 30 |
| Ahrén 2010 Changes in prandial glucagon levels after a 2-year treatment with vildagliptin or glimepiride in patients with type 2 diabetes inadequately controlled with metformin monotherapy Ahrén, B.; Foley, J. E.; Ferrannini, E.; Matthews, D. R.; Zinman, B.; Dejager, S.; Fonseca, V. A.  Diabetes Care Apr 2010;33(4):730-2 | Industry Trial in a journal with IF< 30 |
| Billings 2018 Efficacy and Safety of IDegLira Versus Basal-Bolus Insulin Therapy in Patients With Type 2 Diabetes Uncontrolled on Metformin and Basal Insulin: The DUAL VII Randomized Clinical Trial Billings, L. K.; Doshi, A.; Gouet, D.; Oviedo, A.; Rodbard, H. W.; Tentolouris, N.; Grøn, R.; Halladin, N.; Jodar, E.  Diabetes Care May 2018;41(5):1009-1016 | Industry Trial in a journal with IF< 30 |
| Charbonnel 2006 Efficacy and safety of the dipeptidyl peptidase-4 inhibitor sitagliptin added to ongoing metformin therapy in patients with type 2 diabetes inadequately controlled with metformin alone Charbonnel, B.; Karasik, A.; Liu, J.; Wu, M.; Meininger, G.  Diabetes Care Dec 2006;29(12):2638-43 | Industry Trial in a journal with IF< 30 |
| Standl 2005 Good glycemic control with flexibility in timing of basal insulin supply: a 24-week comparison of insulin glargine given once daily in the morning or at bedtime in combination with morning glimepiride Standl, E.; Maxeiner, S.; Raptis, S.; Karimi-Anderesi, Z.; Schweitzer, M. A.  Diabetes Care Feb 2005;28(2):419-20 | Industry Trial in a journal with IF< 30 |
| Umpierrez 2014 Efficacy and safety of dulaglutide monotherapy versus metformin in type 2 diabetes in a randomized controlled trial (AWARD-3) Umpierrez, G.; Tofé Povedano, S.; Pérez Manghi, F.; Shurzinske, L.; Pechtner, V.  Diabetes Care Aug 2014;37(8):2168-76 | Industry Trial in a journal with IF< 30 |
| Sheu 2015 Safety and Efficacy of Omarigliptin (MK-3102), a Novel Once-Weekly DPP-4 Inhibitor for the Treatment of Patients With Type 2 Diabetes Sheu, W. H.; Gantz, I.; Chen, M.; Suryawanshi, S.; Mirza, A.; Goldstein, B. J.; Kaufman, K. D.; Engel, S. S.  Diabetes Care Nov 2015;38(11):2106-14 | Industry Trial in a journal with IF< 30 |
| Rosenstock 2020 Impact of a Weekly Glucagon-Like Peptide 1 Receptor Agonist, Albiglutide, on Glycemic Control and on Reducing Prandial Insulin Use in Type 2 Diabetes Inadequately Controlled on Multiple Insulin Therapy: A Randomized Trial Rosenstock, J.; Nino, A.; Soffer, J.; Erskine, L.; Acusta, A.; Dole, J.; Carr, M. C.; Mallory, J.; Home, P.  Diabetes Care Oct 2020;43(10):2509-2518 | Industry Trial in a journal with IF< 30 |
| Johansen 2014 C-peptide levels in latent autoimmune diabetes in adults treated with linagliptin versus glimepiride: exploratory results from a 2-year double-blind, randomized, controlled study Johansen, O. E.; Boehm, B. O.; Grill, V.; Torjesen, P. A.; Bhattacharya, S.; Patel, S.; Wetzel, K.; Woerle, H. J.  Diabetes Care 2014;37(1):e11-2 | Industry Trial in a journal with IF< 30 |
| Shepherd 2006 Effect of lowering LDL cholesterol substantially below currently recommended levels in patients with coronary heart disease and diabetes: the Treating to New Targets (TNT) study Shepherd, J.; Barter, P.; Carmena, R.; Deedwania, P.; Fruchart, J. C.; Haffner, S.; Hsia, J.; Breazna, A.; LaRosa, J.; Grundy, S.; Waters, D.  Diabetes Care Jun 2006;29(6):1220-6 | Industry Trial in a journal with IF< 30 |
| Filippatos 2021 Finerenone and Cardiovascular Outcomes in Patients With Chronic Kidney Disease and Type 2 Diabetes Filippatos, G.; Anker, S. D.; Agarwal, R.; Pitt, B.; Ruilope, L. M.; Rossing, P.; Kolkhof, P.; Schloemer, P.; Tornus, I.; Joseph, A.; Bakris, G. L.  Circulation Feb 9 2021;143(6):540-552 | Industry Trial in a journal with IF< 30 |
| Rosenstock 2009 Potential of albiglutide, a long-acting GLP-1 receptor agonist, in type 2 diabetes: a randomized controlled trial exploring weekly, biweekly, and monthly dosing Rosenstock, J.; Reusch, J.; Bush, M.; Yang, F.; Stewart, M.  Diabetes Care Oct 2009;32(10):1880-6 | Industry Trial in a journal with IF< 30 |
| Zinman 2009 Efficacy and safety of the human glucagon-like peptide-1 analog liraglutide in combination with metformin and thiazolidinedione in patients with type 2 diabetes (LEAD-4 Met+TZD) Zinman, B.; Gerich, J.; Buse, J. B.; Lewin, A.; Schwartz, S.; Raskin, P.; Hale, P. M.; Zdravkovic, M.; Blonde, L.  Diabetes Care Jul 2009;32(7):1224-30 | Industry Trial in a journal with IF< 30 |
| Raskin 2000 Repaglinide/troglitazone combination therapy: improved glycemic control in type 2 diabetes Raskin, P.; Jovanovic, L.; Berger, S.; Schwartz, S.; Woo, V.; Ratner, R.  Diabetes Care Jul 2000;23(7):979-83 | Industry Trial in a journal with IF< 30 |
| Davies 2005 Improvement of glycemic control in subjects with poorly controlled type 2 diabetes: comparison of two treatment algorithms using insulin glargine Davies, M.; Storms, F.; Shutler, S.; Bianchi-Biscay, M.; Gomis, R.  Diabetes Care Jun 2005;28(6):1282-8 | Industry Trial in a journal with IF< 30 |
| Bosi 2007 Effects of vildagliptin on glucose control over 24 weeks in patients with type 2 diabetes inadequately controlled with metformin Bosi, E.; Camisasca, R. P.; Collober, C.; Rochotte, E.; Garber, A. J.  Diabetes Care Apr 2007;30(4):890-5 | Industry Trial in a journal with IF< 30 |
| Schernthaner 2013 Canagliflozin compared with sitagliptin for patients with type 2 diabetes who do not have adequate glycemic control with metformin plus sulfonylurea: a 52-week randomized trial Schernthaner, G.; Gross, J. L.; Rosenstock, J.; Guarisco, M.; Fu, M.; Yee, J.; Kawaguchi, M.; Canovatchel, W.; Meininger, G.  Diabetes Care Sep 2013;36(9):2508-15 | Industry Trial in a journal with IF< 30 |
| Tan 2005 Comparison of pioglitazone and gliclazide in sustaining glycemic control over 2 years in patients with type 2 diabetes Tan, M. H.; Baksi, A.; Krahulec, B.; Kubalski, P.; Stankiewicz, A.; Urquhart, R.; Edwards, G.; Johns, D.  Diabetes Care Mar 2005;28(3):544-50 | Industry Trial in a journal with IF< 30 |
| Aschner 2006 Effect of the dipeptidyl peptidase-4 inhibitor sitagliptin as monotherapy on glycemic control in patients with type 2 diabetes Aschner, P.; Kipnes, M. S.; Lunceford, J. K.; Sanchez, M.; Mickel, C.; Williams-Herman, D. E.  Diabetes Care Dec 2006;29(12):2632-7 | Industry Trial in a journal with IF< 30 |
| Rosenstock 2008 SERENADE: the Study Evaluating Rimonabant Efficacy in Drug-naive Diabetic Patients: effects of monotherapy with rimonabant, the first selective CB1 receptor antagonist, on glycemic control, body weight, and lipid profile in drug-naive type 2 diabetes Rosenstock, J.; Hollander, P.; Chevalier, S.; Iranmanesh, A.  Diabetes Care Nov 2008;31(11):2169-76 | Industry Trial in a journal with IF< 30 |
| Tikkanen 2015 Empagliflozin reduces blood pressure in patients with type 2 diabetes and hypertension Tikkanen, I.; Narko, K.; Zeller, C.; Green, A.; Salsali, A.; Broedl, U. C.; Woerle, H. J.  Diabetes Care Mar 2015;38(3):420-8 | Industry Trial in a journal with IF< 30 |
| Rosenstock 2016 Prandial Options to Advance Basal Insulin Glargine Therapy: Testing Lixisenatide Plus Basal Insulin Versus Insulin Glulisine Either as Basal-Plus or Basal-Bolus in Type 2 Diabetes: The GetGoal Duo-2 Trial Rosenstock, J.; Guerci, B.; Hanefeld, M.; Gentile, S.; Aronson, R.; Tinahones, F. J.; Roy-Duval, C.; Souhami, E.; Wardecki, M.; Ye, J.; Perfetti, R.; Heller, S.  Diabetes Care Aug 2016;39(8):1318-28 | Industry Trial in a journal with IF< 30 |
| Meneghini 2013 The efficacy and safety of insulin degludec given in variable once-daily dosing intervals compared with insulin glargine and insulin degludec dosed at the same time daily: a 26-week, randomized, open-label, parallel-group, treat-to-target trial in individuals with type 2 diabetes Meneghini, L.; Atkin, S. L.; Gough, S. C.; Raz, I.; Blonde, L.; Shestakova, M.; Bain, S.; Johansen, T.; Begtrup, K.; Birkeland, K. I.  Diabetes Care Apr 2013;36(4):858-64 | Industry Trial in a journal with IF< 30 |
| Vilsbøll 2019 Dapagliflozin Plus Saxagliptin Add-on Therapy Compared With Insulin in Patients With Type 2 Diabetes Poorly Controlled by Metformin With or Without Sulfonylurea Therapy: A Randomized Clinical Trial Vilsbøll, T.; Ekholm, E.; Johnsson, E.; Dronamraju, N.; Jabbour, S.; Lind, M.  Diabetes Care Aug 2019;42(8):1464-1472 | Industry Trial in a journal with IF< 30 |
| Blonde 2019 Switching to iGlarLixi Versus Continuing Daily or Weekly GLP-1 RA in Type 2 Diabetes Inadequately Controlled by GLP-1 RA and Oral Antihyperglycemic Therapy: The LixiLan-G Randomized Clinical Trial Blonde, L.; Rosenstock, J.; Del Prato, S.; Henry, R.; Shehadeh, N.; Frias, J.; Niemoeller, E.; Souhami, E.; Ji, C.; Aroda, V. R.  Diabetes Care Nov 2019;42(11):2108-2116 | Industry Trial in a journal with IF< 30 |
| Hanefeld 2000 Rapid and short-acting mealtime insulin secretion with nateglinide controls both prandial and mean glycemia Hanefeld, M.; Bouter, K. P.; Dickinson, S.; Guitard, C.  Diabetes Care Feb 2000;23(2):202-7 | Industry Trial in a journal with IF< 30 |
| Hadjadj 2016 Initial Combination of Empagliflozin and Metformin in Patients With Type 2 Diabetes Hadjadj, S.; Rosenstock, J.; Meinicke, T.; Woerle, H. J.; Broedl, U. C.  Diabetes Care Oct 2016;39(10):1718-28 | Industry Trial in a journal with IF< 30 |
| Søfteland 2017 Empagliflozin as Add-on Therapy in Patients With Type 2 Diabetes Inadequately Controlled With Linagliptin and Metformin: A 24-Week Randomized, Double-Blind, Parallel-Group Trial Søfteland, E.; Meier, J. J.; Vangen, B.; Toorawa, R.; Maldonado-Lutomirsky, M.; Broedl, U. C.  Diabetes Care Feb 2017;40(2):201-209 | Industry Trial in a journal with IF< 30 |
| Tuttle 2005 The effect of ruboxistaurin on nephropathy in type 2 diabetes Tuttle, K. R.; Bakris, G. L.; Toto, R. D.; McGill, J. B.; Hu, K.; Anderson, P. W.  Diabetes Care Nov 2005;28(11):2686-90 | Industry Trial in a journal with IF< 30 |
| Ahrén 2014 HARMONY 3: 104-week randomized, double-blind, placebo- and active-controlled trial assessing the efficacy and safety of albiglutide compared with placebo, sitagliptin, and glimepiride in patients with type 2 diabetes taking metformin Ahrén, B.; Johnson, S. L.; Stewart, M.; Cirkel, D. T.; Yang, F.; Perry, C.; Feinglos, M. N.  Diabetes Care Aug 2014;37(8):2141-8 | Industry Trial in a journal with IF< 30 |
| Nauck 2009 Efficacy and safety comparison of liraglutide, glimepiride, and placebo, all in combination with metformin, in type 2 diabetes: the LEAD (liraglutide effect and action in diabetes)-2 study Nauck, M.; Frid, A.; Hermansen, K.; Shah, N. S.; Tankova, T.; Mitha, I. H.; Zdravkovic, M.; Düring, M.; Matthews, D. R.  Diabetes Care Jan 2009;32(1):84-90 | Industry Trial in a journal with IF< 30 |
| Heise 2011 A new-generation ultra-long-acting basal insulin with a bolus boost compared with insulin glargine in insulin-naive people with type 2 diabetes: a randomized, controlled trial Heise, T.; Tack, C. J.; Cuddihy, R.; Davidson, J.; Gouet, D.; Liebl, A.; Romero, E.; Mersebach, H.; Dykiel, P.; Jorde, R.  Diabetes Care Mar 2011;34(3):669-74 | Industry Trial in a journal with IF< 30 |
| Rosenstock 2018 More Similarities Than Differences Testing Insulin Glargine 300 Units/mL Versus Insulin Degludec 100 Units/mL in Insulin-Naive Type 2 Diabetes: The Randomized Head-to-Head BRIGHT Trial Rosenstock, J.; Cheng, A.; Ritzel, R.; Bosnyak, Z.; Devisme, C.; Cali, A. M. G.; Sieber, J.; Stella, P.; Wang, X.; Frías, J. P.; Roussel, R.; Bolli, G. B.  Diabetes Care Oct 2018;41(10):2147-2154 | Industry Trial in a journal with IF< 30 |
| Davies 2016 Efficacy and Safety of Liraglutide Versus Placebo as Add-on to Glucose-Lowering Therapy in Patients With Type 2 Diabetes and Moderate Renal Impairment (LIRA-RENAL): A Randomized Clinical Trial Davies, M. J.; Bain, S. C.; Atkin, S. L.; Rossing, P.; Scott, D.; Shamkhalova, M. S.; Bosch-Traberg, H.; Syrén, A.; Umpierrez, G. E.  Diabetes Care Feb 2016;39(2):222-30 | Industry Trial in a journal with IF< 30 |
| Laakso 2015 Treatment with the dipeptidyl peptidase-4 inhibitor linagliptin or placebo followed by glimepiride in patients with type 2 diabetes with moderate to severe renal impairment: a 52-week, randomized, double-blind clinical trial Laakso, M.; Rosenstock, J.; Groop, P. H.; Barnett, A. H.; Gallwitz, B.; Hehnke, U.; Tamminen, I.; Patel, S.; von Eynatten, M.; Woerle, H. J.  Diabetes Care Feb 2015;38(2):e15-7 | Industry Trial in a journal with IF< 30 |
| Ziegler 2010 Efficacy and safety of lacosamide in painful diabetic neuropathy Ziegler, D.; Hidvégi, T.; Gurieva, I.; Bongardt, S.; Freynhagen, R.; Sen, D.; Sommerville, K.  Diabetes Care Apr 2010;33(4):839-41 | Industry Trial in a journal with IF< 30 |
| Rosenstock 2008 Two-year pulmonary safety and efficacy of inhaled human insulin (Exubera) in adult patients with type 2 diabetes Rosenstock, J.; Cefalu, W. T.; Hollander, P. A.; Belanger, A.; Eliaschewitz, F. G.; Gross, J. L.; Klioze, S. S.; St Aubin, L. B.; Foyt, H.; Ogawa, M.; Duggan, W. T.  Diabetes Care Sep 2008;31(9):1723-8 | Industry Trial in a journal with IF< 30 |
| Buse 2009 DURAbility of basal versus lispro mix 75/25 insulin efficacy (DURABLE) trial 24-week results: safety and efficacy of insulin lispro mix 75/25 versus insulin glargine added to oral antihyperglycemic drugs in patients with type 2 diabetes Buse, J. B.; Wolffenbuttel, B. H.; Herman, W. H.; Shemonsky, N. K.; Jiang, H. H.; Fahrbach, J. L.; Scism-Bacon, J. L.; Martin, S. A.  Diabetes Care Jun 2009;32(6):1007-13 | Industry Trial in a journal with IF< 30 |
| Davies 2013 Once-weekly exenatide versus once- or twice-daily insulin detemir: randomized, open-label, clinical trial of efficacy and safety in patients with type 2 diabetes treated with metformin alone or in combination with sulfonylureas Davies, M.; Heller, S.; Sreenan, S.; Sapin, H.; Adetunji, O.; Tahbaz, A.; Vora, J.  Diabetes Care May 2013;36(5):1368-76 | Industry Trial in a journal with IF< 30 |
| Rosenstock 2014 Advancing basal insulin replacement in type 2 diabetes inadequately controlled with insulin glargine plus oral agents: a comparison of adding albiglutide, a weekly GLP-1 receptor agonist, versus thrice-daily prandial insulin lispro Rosenstock, J.; Fonseca, V. A.; Gross, J. L.; Ratner, R. E.; Ahrén, B.; Chow, F. C.; Yang, F.; Miller, D.; Johnson, S. L.; Stewart, M. W.; Leiter, L. A.  Diabetes Care Aug 2014;37(8):2317-25 | Industry Trial in a journal with IF< 30 |
| Raz 2012 Efficacy and safety of taspoglutide monotherapy in drug-naive type 2 diabetic patients after 24 weeks of treatment: results of a randomized, double-blind, placebo-controlled phase 3 study (T-emerge 1) Raz, I.; Fonseca, V.; Kipnes, M.; Durrwell, L.; Hoekstra, J.; Boldrin, M.; Balena, R.  Diabetes Care Mar 2012;35(3):485-7 | Industry Trial in a journal with IF< 30 |
| #283 Glucose Variability in a 26-Week Randomized Comparison of Mealtime Treatment With Rapid-Acting Insulin Versus GLP-1 Agonist in Participants With Type 2 Diabetes at High Cardiovascular Risk Diabetes Care Jun 2016;39(6):973-81 | Industry Trial in a journal with IF< 30 |
| Rosenstock 2015 Inhaled Technosphere Insulin Versus Inhaled Technosphere Placebo in Insulin-Naïve Subjects With Type 2 Diabetes Inadequately Controlled on Oral Antidiabetes Agents Rosenstock, J.; Franco, D.; Korpachev, V.; Shumel, B.; Ma, Y.; Baughman, R.; Amin, N.; McGill, J. B.  Diabetes Care Dec 2015;38(12):2274-81 | Industry Trial in a journal with IF< 30 |
| McNulty 2003 A randomized trial of sibutramine in the management of obese type 2 diabetic patients treated with metformin McNulty, S. J.; Ur, E.; Williams, G.  Diabetes Care Jan 2003;26(1):125-31 | Industry Trial in a journal with IF< 30 |
| Figtree 2019 Effects of Canagliflozin on Heart Failure Outcomes Associated With Preserved and Reduced Ejection Fraction in Type 2 Diabetes Mellitus Figtree, G. A.; Rådholm, K.; Barrett, T. D.; Perkovic, V.; Mahaffey, K. W.; de Zeeuw, D.; Fulcher, G.; Matthews, D. R.; Shaw, W.; Neal, B.  Circulation May 28 2019;139(22):2591-2593 | Industry Trial in a journal with IF< 30 |
| Häring 2014 Empagliflozin as add-on to metformin in patients with type 2 diabetes: a 24-week, randomized, double-blind, placebo-controlled trial Häring, H. U.; Merker, L.; Seewaldt-Becker, E.; Weimer, M.; Meinicke, T.; Broedl, U. C.; Woerle, H. J.  Diabetes Care Jun 2014;37(6):1650-9 | Industry Trial in a journal with IF< 30 |
| Nauck 2016 Once-Daily Liraglutide Versus Lixisenatide as Add-on to Metformin in Type 2 Diabetes: A 26-Week Randomized Controlled Clinical Trial Nauck, M.; Rizzo, M.; Johnson, A.; Bosch-Traberg, H.; Madsen, J.; Cariou, B.  Diabetes Care Sep 2016;39(9):1501-9 | Industry Trial in a journal with IF< 30 |
| Rosenstock 2015 Dual add-on therapy in type 2 diabetes poorly controlled with metformin monotherapy: a randomized double-blind trial of saxagliptin plus dapagliflozin addition versus single addition of saxagliptin or dapagliflozin to metformin Rosenstock, J.; Hansen, L.; Zee, P.; Li, Y.; Cook, W.; Hirshberg, B.; Iqbal, N.  Diabetes Care Mar 2015;38(3):376-83 | Industry Trial in a journal with IF< 30 |
| Zinman 2012 Insulin degludec versus insulin glargine in insulin-naive patients with type 2 diabetes: a 1-year, randomized, treat-to-target trial (BEGIN Once Long) Zinman, B.; Philis-Tsimikas, A.; Cariou, B.; Handelsman, Y.; Rodbard, H. W.; Johansen, T.; Endahl, L.; Mathieu, C.  Diabetes Care Dec 2012;35(12):2464-71 | Industry Trial in a journal with IF< 30 |
| Bordeleau 2014 The association of basal insulin glargine and/or n-3 fatty acids with incident cancers in patients with dysglycemia Bordeleau, L.; Yakubovich, N.; Dagenais, G. R.; Rosenstock, J.; Probstfield, J.; Chang Yu, P.; Ryden, L. E.; Pirags, V.; Spinas, G. A.; Birkeland, K. I.; Ratner, R. E.; Marin-Neto, J. A.; Keltai, M.; Riddle, M. C.; Bosch, J.; Yusuf, S.; Gerstein, H. C.  Diabetes Care 2014;37(5):1360-6 | Industry Trial in a journal with IF< 30 |
| Rosenstock 2016 Efficacy and Safety of LixiLan, a Titratable Fixed-Ratio Combination of Lixisenatide and Insulin Glargine, Versus Insulin Glargine in Type 2 Diabetes Inadequately Controlled on Metformin Monotherapy: The LixiLan Proof-of-Concept Randomized Trial Rosenstock, J.; Diamant, M.; Aroda, V. R.; Silvestre, L.; Souhami, E.; Zhou, T.; Perfetti, R.; Fonseca, V.  Diabetes Care Sep 2016;39(9):1579-86 | Industry Trial in a journal with IF< 30 |
| Pratley 2012 Efficacy and safety of switching from the DPP-4 inhibitor sitagliptin to the human GLP-1 analog liraglutide after 52 weeks in metformin-treated patients with type 2 diabetes: a randomized, open-label trial Pratley, R. E.; Nauck, M. A.; Bailey, T.; Montanya, E.; Filetti, S.; Garber, A. J.; Thomsen, A. B.; Furber, S.; Davies, M.  Diabetes Care Oct 2012;35(10):1986-93 | Industry Trial in a journal with IF< 30 |
| Gerstein 2010 Effect of rosiglitazone on progression of coronary atherosclerosis in patients with type 2 diabetes mellitus and coronary artery disease: the assessment on the prevention of progression by rosiglitazone on atherosclerosis in diabetes patients with cardiovascular history trial Gerstein, H. C.; Ratner, R. E.; Cannon, C. P.; Serruys, P. W.; García-García, H. M.; van Es, G. A.; Kolatkar, N. S.; Kravitz, B. G.; Miller, D. M.; Huang, C.; Fitzgerald, P. J.; Nesto, R. W.  Circulation Mar 16 2010;121(10):1176-87 | Industry Trial in a journal with IF< 30 |
| Menon 2018 Fasiglifam-Induced Liver Injury in Patients With Type 2 Diabetes: Results of a Randomized Controlled Cardiovascular Outcomes Safety Trial Menon, V.; Lincoff, A. M.; Nicholls, S. J.; Jasper, S.; Wolski, K.; McGuire, D. K.; Mehta, C. R.; Rosenstock, J.; Lopez, C.; Marcinak, J.; Cao, C.; Nissen, S. E.  Diabetes Care Dec 2018;41(12):2603-2609 | Industry Trial in a journal with IF< 30 |
| Sever 2005 Reduction in cardiovascular events with atorvastatin in 2,532 patients with type 2 diabetes: Anglo-Scandinavian Cardiac Outcomes Trial--lipid-lowering arm (ASCOT-LLA) Sever, P. S.; Poulter, N. R.; Dahlöf, B.; Wedel, H.; Collins, R.; Beevers, G.; Caulfield, M.; Kjeldsen, S. E.; Kristinsson, A.; McInnes, G. T.; Mehlsen, J.; Nieminen, M.; O'Brien, E.; Ostergren, J.  Diabetes Care May 2005;28(5):1151-7 | Industry Trial in a journal with IF< 30 |
| DeFronzo 2010 Effects of exenatide plus rosiglitazone on beta-cell function and insulin sensitivity in subjects with type 2 diabetes on metformin DeFronzo, R. A.; Triplitt, C.; Qu, Y.; Lewis, M. S.; Maggs, D.; Glass, L. C.  Diabetes Care May 2010;33(5):951-7 | Industry Trial in a journal with IF< 30 |
| Fulcher 2014 Comparison of insulin degludec/insulin aspart and biphasic insulin aspart 30 in uncontrolled, insulin-treated type 2 diabetes: a phase 3a, randomized, treat-to-target trial Fulcher, G. R.; Christiansen, J. S.; Bantwal, G.; Polaszewska-Muszynska, M.; Mersebach, H.; Andersen, T. H.; Niskanen, L. K.  Diabetes Care Aug 2014;37(8):2084-90 | Industry Trial in a journal with IF< 30 |
| Fouqueray 2013 The efficacy and safety of imeglimin as add-on therapy in patients with type 2 diabetes inadequately controlled with metformin monotherapy Fouqueray, P.; Pirags, V.; Inzucchi, S. E.; Bailey, C. J.; Schernthaner, G.; Diamant, M.; Lebovitz, H. E.  Diabetes Care Mar 2013;36(3):565-8 | Industry Trial in a journal with IF< 30 |
| Barnett 2006 An open, randomized, parallel-group study to compare the efficacy and safety profile of inhaled human insulin (Exubera) with metformin as adjunctive therapy in patients with type 2 diabetes poorly controlled on a sulfonylurea Barnett, A. H.; Dreyer, M.; Lange, P.; Serdarevic-Pehar, M.  Diabetes Care Jun 2006;29(6):1282-7 | Industry Trial in a journal with IF< 30 |
| List 2009 Sodium-glucose cotransport inhibition with dapagliflozin in type 2 diabetes List, J. F.; Woo, V.; Morales, E.; Tang, W.; Fiedorek, F. T.  Diabetes Care Apr 2009;32(4):650-7 | Industry Trial in a journal with IF< 30 |
| Riddle 2003 The treat-to-target trial: randomized addition of glargine or human NPH insulin to oral therapy of type 2 diabetic patients Riddle, M. C.; Rosenstock, J.; Gerich, J.  Diabetes Care Nov 2003;26(11):3080-6 | Industry Trial in a journal with IF< 30 |
| Rosenstock 2012 Effects of dapagliflozin, an SGLT2 inhibitor, on HbA(1c), body weight, and hypoglycemia risk in patients with type 2 diabetes inadequately controlled on pioglitazone monotherapy Rosenstock, J.; Vico, M.; Wei, L.; Salsali, A.; List, J. F.  Diabetes Care Jul 2012;35(7):1473-8 | Industry Trial in a journal with IF< 30 |
| ArjonaFerreira 2013 Efficacy and safety of sitagliptin versus glipizide in patients with type 2 diabetes and moderate-to-severe chronic renal insufficiency Arjona Ferreira, J. C.; Marre, M.; Barzilai, N.; Guo, H.; Golm, G. T.; Sisk, C. M.; Kaufman, K. D.; Goldstein, B. J.  Diabetes Care May 2013;36(5):1067-73 | Industry Trial in a journal with IF< 30 |
| Lane 2020 A Randomized Trial Evaluating the Efficacy and Safety of Fast-Acting Insulin Aspart Compared With Insulin Aspart, Both in Combination With Insulin Degludec With or Without Metformin, in Adults With Type 2 Diabetes (ONSET 9) Lane, W. S.; Favaro, E.; Rathor, N.; Jang, H. C.; Kjærsgaard, M. I. S.; Oviedo, A.; Rose, L.; Senior, P.; Sesti, G.; Soto Gonzalez, A.; Franek, E.  Diabetes Care Aug 2020;43(8):1710-1716 | Industry Trial in a journal with IF< 30 |
| Meneilly 2017 Lixisenatide Therapy in Older Patients With Type 2 Diabetes Inadequately Controlled on Their Current Antidiabetic Treatment: The GetGoal-O Randomized Trial Meneilly, G. S.; Roy-Duval, C.; Alawi, H.; Dailey, G.; Bellido, D.; Trescoli, C.; Manrique Hurtado, H.; Guo, H.; Pilorget, V.; Perfetti, R.; Simpson, H.  Diabetes Care Apr 2017;40(4):485-493 | Industry Trial in a journal with IF< 30 |
| Fonseca 2012 Efficacy and safety of the once-daily GLP-1 receptor agonist lixisenatide in monotherapy: a randomized, double-blind, placebo-controlled trial in patients with type 2 diabetes (GetGoal-Mono) Fonseca, V. A.; Alvarado-Ruiz, R.; Raccah, D.; Boka, G.; Miossec, P.; Gerich, J. E.  Diabetes Care Jun 2012;35(6):1225-31 | Industry Trial in a journal with IF< 30 |
| Ahmann 2018 Efficacy and Safety of Once-Weekly Semaglutide Versus Exenatide ER in Subjects With Type 2 Diabetes (SUSTAIN 3): A 56-Week, Open-Label, Randomized Clinical Trial Ahmann, A. J.; Capehorn, M.; Charpentier, G.; Dotta, F.; Henkel, E.; Lingvay, I.; Holst, A. G.; Annett, M. P.; Aroda, V. R.  Diabetes Care Feb 2018;41(2):258-266 | Industry Trial in a journal with IF< 30 |
| Bergenstal 2012 A randomized, controlled study of once-daily LY2605541, a novel long-acting basal insulin, versus insulin glargine in basal insulin-treated patients with type 2 diabetes Bergenstal, R. M.; Rosenstock, J.; Arakaki, R. F.; Prince, M. J.; Qu, Y.; Sinha, V. P.; Howey, D. C.; Jacober, S. J.  Diabetes Care Nov 2012;35(11):2140-7 | Industry Trial in a journal with IF< 30 |
| Rosenstock 2013 Efficacy and safety of lixisenatide once daily versus exenatide twice daily in type 2 diabetes inadequately controlled on metformin: a 24-week, randomized, open-label, active-controlled study (GetGoal-X) Rosenstock, J.; Raccah, D.; Korányi, L.; Maffei, L.; Boka, G.; Miossec, P.; Gerich, J. E.  Diabetes Care Oct 2013;36(10):2945-51 | Industry Trial in a journal with IF< 30 |
| Hollander 2004 Efficacy and safety of inhaled insulin (exubera) compared with subcutaneous insulin therapy in patients with type 2 diabetes: results of a 6-month, randomized, comparative trial Hollander, P. A.; Blonde, L.; Rowe, R.; Mehta, A. E.; Milburn, J. L.; Hershon, K. S.; Chiasson, J. L.; Levin, S. R.  Diabetes Care Oct 2004;27(10):2356-62 | Industry Trial in a journal with IF< 30 |
| Nauck 2011 Dapagliflozin versus glipizide as add-on therapy in patients with type 2 diabetes who have inadequate glycemic control with metformin: a randomized, 52-week, double-blind, active-controlled noninferiority trial Nauck, M. A.; Del Prato, S.; Meier, J. J.; Durán-García, S.; Rohwedder, K.; Elze, M.; Parikh, S. J.  Diabetes Care Sep 2011;34(9):2015-22 | Industry Trial in a journal with IF< 30 |
| Giugliano 2014 Initiation and gradual intensification of premixed insulin lispro therapy versus Basal {+/-} mealtime insulin in patients with type 2 diabetes eating light breakfasts Giugliano, D.; Tracz, M.; Shah, S.; Calle-Pascual, A.; Mistodie, C.; Duarte, R.; Sari, R.; Woo, V.; Jiletcovici, A. O.; Deinhard, J.; Wille, S. A.; Kiljanski, J.  Diabetes Care Feb 2014;37(2):372-80 | Industry Trial in a journal with IF< 30 |
| Janka 2005 Comparison of basal insulin added to oral agents versus twice-daily premixed insulin as initial insulin therapy for type 2 diabetes Janka, H. U.; Plewe, G.; Riddle, M. C.; Kliebe-Frisch, C.; Schweitzer, M. A.; Yki-Järvinen, H.  Diabetes Care Feb 2005;28(2):254-9 | Industry Trial in a journal with IF< 30 |
| Andersen 2003 Kidney function during and after withdrawal of long-term irbesartan treatment in patients with type 2 diabetes and microalbuminuria Andersen, S.; Bröchner-Mortensen, J.; Parving, H. H.  Diabetes Care Dec 2003;26(12):3296-302 | Industry Trial in a journal with IF< 30 |
| Blevins 2020 Randomized Double-Blind Clinical Trial Comparing Ultra Rapid Lispro With Lispro in a Basal-Bolus Regimen in Patients With Type 2 Diabetes: PRONTO-T2D Blevins, T.; Zhang, Q.; Frias, J. P.; Jinnouchi, H.; Chang, A. M.  Diabetes Care Dec 2020;43(12):2991-2998 | Industry Trial in a journal with IF< 30 |
| Matthaei 2015 Randomized, Double-Blind Trial of Triple Therapy With Saxagliptin Add-on to Dapagliflozin Plus Metformin in Patients With Type 2 Diabetes Matthaei, S.; Catrinoiu, D.; Celiński, A.; Ekholm, E.; Cook, W.; Hirshberg, B.; Chen, H.; Iqbal, N.; Hansen, L.  Diabetes Care Nov 2015;38(11):2018-24 | Industry Trial in a journal with IF< 30 |
| Rosenstock 2016 Benefits of LixiLan, a Titratable Fixed-Ratio Combination of Insulin Glargine Plus Lixisenatide, Versus Insulin Glargine and Lixisenatide Monocomponents in Type 2 Diabetes Inadequately Controlled on Oral Agents: The LixiLan-O Randomized Trial Rosenstock, J.; Aronson, R.; Grunberger, G.; Hanefeld, M.; Piatti, P.; Serusclat, P.; Cheng, X.; Zhou, T.; Niemoeller, E.; Souhami, E.; Davies, M.  Diabetes Care Nov 2016;39(11):2026-2035 | Industry Trial in a journal with IF< 30 |
| Zinman 2019 Efficacy, Safety, and Tolerability of Oral Semaglutide Versus Placebo Added to Insulin With or Without Metformin in Patients With Type 2 Diabetes: The PIONEER 8 Trial Zinman, B.; Aroda, V. R.; Buse, J. B.; Cariou, B.; Harris, S. B.; Hoff, S. T.; Pedersen, K. B.; Tarp-Johansen, M. J.; Araki, E.  Diabetes Care Dec 2019;42(12):2262-2271 | Industry Trial in a journal with IF< 30 |
| Aroda 2016 Efficacy and Safety of LixiLan, a Titratable Fixed-Ratio Combination of Insulin Glargine Plus Lixisenatide in Type 2 Diabetes Inadequately Controlled on Basal Insulin and Metformin: The LixiLan-L Randomized Trial Aroda, V. R.; Rosenstock, J.; Wysham, C.; Unger, J.; Bellido, D.; González-Gálvez, G.; Takami, A.; Guo, H.; Niemoeller, E.; Souhami, E.; Bergenstal, R. M.  Diabetes Care Nov 2016;39(11):1972-1980 | Industry Trial in a journal with IF< 30 |
| Raz 2009 Effects of prandial versus fasting glycemia on cardiovascular outcomes in type 2 diabetes: the HEART2D trial Raz, I.; Wilson, P. W.; Strojek, K.; Kowalska, I.; Bozikov, V.; Gitt, A. K.; Jermendy, G.; Campaigne, B. N.; Kerr, L.; Milicevic, Z.; Jacober, S. J.  Diabetes Care Mar 2009;32(3):381-6 | Industry Trial in a journal with IF< 30 |
| Lingvay 2018 A 26-Week Randomized Controlled Trial of Semaglutide Once Daily Versus Liraglutide and Placebo in Patients With Type 2 Diabetes Suboptimally Controlled on Diet and Exercise With or Without Metformin Lingvay, I.; Desouza, C. V.; Lalic, K. S.; Rose, L.; Hansen, T.; Zacho, J.; Pieber, T. R.  Diabetes Care Sep 2018;41(9):1926-1937 | Industry Trial in a journal with IF< 30 |
| Moses 2001 Flexible meal-related dosing with repaglinide facilitates glycemic control in therapy-naive type 2 diabetes Moses, R. G.; Gomis, R.; Frandsen, K. B.; Schlienger, J. L.; Dedov, I.  Diabetes Care Jan 2001;24(1):11-5 | Industry Trial in a journal with IF< 30 |
| Knopp 2006 Efficacy and safety of atorvastatin in the prevention of cardiovascular end points in subjects with type 2 diabetes: the Atorvastatin Study for Prevention of Coronary Heart Disease Endpoints in non-insulin-dependent diabetes mellitus (ASPEN) Knopp, R. H.; d'Emden, M.; Smilde, J. G.; Pocock, S. J.  Diabetes Care Jul 2006;29(7):1478-85 | Industry Trial in a journal with IF< 30 |
| Buse 2014 Contribution of liraglutide in the fixed-ratio combination of insulin degludec and liraglutide (IDegLira) Buse, J. B.; Vilsbøll, T.; Thurman, J.; Blevins, T. C.; Langbakke, I. H.; Bøttcher, S. G.; Rodbard, H. W.  Diabetes Care Nov 2014;37(11):2926-33 | Industry Trial in a journal with IF< 30 |
| Yki-Järvinen 2007 Initiate Insulin by Aggressive Titration and Education (INITIATE): a randomized study to compare initiation of insulin combination therapy in type 2 diabetic patients individually and in groups Yki-Järvinen, H.; Juurinen, L.; Alvarsson, M.; Bystedt, T.; Caldwell, I.; Davies, M.; Lahdenperä, S.; Nijpels, G.; Vähätalo, M.  Diabetes Care Jun 2007;30(6):1364-9 | Industry Trial in a journal with IF< 30 |
| Jabbour 2014 Dapagliflozin is effective as add-on therapy to sitagliptin with or without metformin: a 24-week, multicenter, randomized, double-blind, placebo-controlled study Jabbour, S. A.; Hardy, E.; Sugg, J.; Parikh, S.  Diabetes Care 2014;37(3):740-50 | Industry Trial in a journal with IF< 30 |
| Riddle 2013 Adding once-daily lixisenatide for type 2 diabetes inadequately controlled with newly initiated and continuously titrated basal insulin glargine: a 24-week, randomized, placebo-controlled study (GetGoal-Duo 1) Riddle, M. C.; Forst, T.; Aronson, R.; Sauque-Reyna, L.; Souhami, E.; Silvestre, L.; Ping, L.; Rosenstock, J.  Diabetes Care Sep 2013;36(9):2497-503 | Industry Trial in a journal with IF< 30 |
| Gerich 2005 PRESERVE-beta: two-year efficacy and safety of initial combination therapy with nateglinide or glyburide plus metformin Gerich, J.; Raskin, P.; Jean-Louis, L.; Purkayastha, D.; Baron, M. A.  Diabetes Care Sep 2005;28(9):2093-9 | Industry Trial in a journal with IF< 30 |
| Madsbad 2004 Improved glycemic control with no weight increase in patients with type 2 diabetes after once-daily treatment with the long-acting glucagon-like peptide 1 analog liraglutide (NN2211): a 12-week, double-blind, randomized, controlled trial Madsbad, S.; Schmitz, O.; Ranstam, J.; Jakobsen, G.; Matthews, D. R.  Diabetes Care Jun 2004;27(6):1335-42 | Industry Trial in a journal with IF< 30 |
| Gerstein 2018 Effect of Basal Insulin Glargine on First and Recurrent Episodes of Heart Failure Hospitalization: The ORIGIN Trial (Outcome Reduction With Initial Glargine Intervention) Gerstein, H. C.; Jung, H.; Rydén, L.; Diaz, R.; Gilbert, R. E.; Yusuf, S.  Circulation Jan 2 2018;137(1):88-90 | Industry Trial in a journal with IF< 30 |
| #3 Predictors of nonsevere and severe hypoglycemia during glucose-lowering treatment with insulin glargine or standard drugs in the ORIGIN trial Diabetes Care Jan 2015;38(1):22-8 | Industry Trial in a journal with IF< 30 |
| Jabbour 2018 Safety and Efficacy of Exenatide Once Weekly Plus Dapagliflozin Once Daily Versus Exenatide or Dapagliflozin Alone in Patients With Type 2 Diabetes Inadequately Controlled With Metformin Monotherapy: 52-Week Results of the DURATION-8 Randomized Controlled Trial Jabbour, S. A.; Frías, J. P.; Hardy, E.; Ahmed, A.; Wang, H.; Öhman, P.; Guja, C.  Diabetes Care Oct 2018;41(10):2136-2146 | Industry Trial in a journal with IF< 30 |
| Kahn 2008 Rosiglitazone-associated fractures in type 2 diabetes: an Analysis from A Diabetes Outcome Progression Trial (ADOPT) Kahn, S. E.; Zinman, B.; Lachin, J. M.; Haffner, S. M.; Herman, W. H.; Holman, R. R.; Kravitz, B. G.; Yu, D.; Heise, M. A.; Aftring, R. P.; Viberti, G.  Diabetes Care May 2008;31(5):845-51 | Industry Trial in a journal with IF< 30 |
| Bhatt 2020 Role of Combination Antiplatelet and Anticoagulation Therapy in Diabetes Mellitus and Cardiovascular Disease: Insights From the COMPASS Trial Bhatt, D. L.; Eikelboom, J. W.; Connolly, S. J.; Steg, P. G.; Anand, S. S.; Verma, S.; Branch, K. R. H.; Probstfield, J.; Bosch, J.; Shestakovska, O.; Szarek, M.; Maggioni, A. P.; Widimský, P.; Avezum, A.; Diaz, R.; Lewis, B. S.; Berkowitz, S. D.; Fox, K. A. A.; Ryden, L.; Yusuf, S.  Circulation Jun 9 2020;141(23):1841-1854 | Industry Trial in a journal with IF< 30 |
| Jabbour 2020 Efficacy and Safety Over 2 Years of Exenatide Plus Dapagliflozin in the DURATION-8 Study: A Multicenter, Double-Blind, Phase 3, Randomized Controlled Trial Jabbour, S. A.; Frías, J. P.; Ahmed, A.; Hardy, E.; Choi, J.; Sjöström, C. D.; Guja, C.  Diabetes Care Oct 2020;43(10):2528-2536 | Industry Trial in a journal with IF< 30 |
| Riddle 2014 New insulin glargine 300 units/mL versus glargine 100 units/mL in people with type 2 diabetes using basal and mealtime insulin: glucose control and hypoglycemia in a 6-month randomized controlled trial (EDITION 1) Riddle, M. C.; Bolli, G. B.; Ziemen, M.; Muehlen-Bartmer, I.; Bizet, F.; Home, P. D.  Diabetes Care Oct 2014;37(10):2755-62 | Industry Trial in a journal with IF< 30 |
| Ahrén 2013 Efficacy and safety of lixisenatide once-daily morning or evening injections in type 2 diabetes inadequately controlled on metformin (GetGoal-M) Ahrén, B.; Leguizamo Dimas, A.; Miossec, P.; Saubadu, S.; Aronson, R.  Diabetes Care Sep 2013;36(9):2543-50 | Industry Trial in a journal with IF< 30 |
| Häring 2013 Empagliflozin as add-on to metformin plus sulfonylurea in patients with type 2 diabetes: a 24-week, randomized, double-blind, placebo-controlled trial Häring, H. U.; Merker, L.; Seewaldt-Becker, E.; Weimer, M.; Meinicke, T.; Woerle, H. J.; Broedl, U. C.  Diabetes Care Nov 2013;36(11):3396-404 | Industry Trial in a journal with IF< 30 |
| McGill 2013 Long-term efficacy and safety of linagliptin in patients with type 2 diabetes and severe renal impairment: a 1-year, randomized, double-blind, placebo-controlled study McGill, J. B.; Sloan, L.; Newman, J.; Patel, S.; Sauce, C.; von Eynatten, M.; Woerle, H. J.  Diabetes Care Feb 2013;36(2):237-44 | Industry Trial in a journal with IF< 30 |
| Yki-Järvinen 2013 Effects of adding linagliptin to basal insulin regimen for inadequately controlled type 2 diabetes: a ≥52-week randomized, double-blind study Yki-Järvinen, H.; Rosenstock, J.; Durán-Garcia, S.; Pinnetti, S.; Bhattacharya, S.; Thiemann, S.; Patel, S.; Woerle, H. J.  Diabetes Care Dec 2013;36(12):3875-81 | Industry Trial in a journal with IF< 30 |
| Hanefeld 2004 One-year glycemic control with a sulfonylurea plus pioglitazone versus a sulfonylurea plus metformin in patients with type 2 diabetes Hanefeld, M.; Brunetti, P.; Schernthaner, G. H.; Matthews, D. R.; Charbonnel, B. H.  Diabetes Care Jan 2004;27(1):141-7 | Industry Trial in a journal with IF< 30 |
| DeFronzo 2008 Efficacy and safety of the dipeptidyl peptidase-4 inhibitor alogliptin in patients with type 2 diabetes and inadequate glycemic control: a randomized, double-blind, placebo-controlled study DeFronzo, R. A.; Fleck, P. R.; Wilson, C. A.; Mekki, Q.  Diabetes Care Dec 2008;31(12):2315-7 | Industry Trial in a journal with IF< 30 |
| Lewin 2015 Initial combination of empagliflozin and linagliptin in subjects with type 2 diabetes Lewin, A.; DeFronzo, R. A.; Patel, S.; Liu, D.; Kaste, R.; Woerle, H. J.; Broedl, U. C.  Diabetes Care Mar 2015;38(3):394-402 | Industry Trial in a journal with IF< 30 |
| Frykberg 2020 A Multinational, Multicenter, Randomized, Double-Blinded, Placebo-Controlled Trial to Evaluate the Efficacy of Cyclical Topical Wound Oxygen (TWO2) Therapy in the Treatment of Chronic Diabetic Foot Ulcers: The TWO2 Study Frykberg, R. G.; Franks, P. J.; Edmonds, M.; Brantley, J. N.; Téot, L.; Wild, T.; Garoufalis, M. G.; Lee, A. M.; Thompson, J. A.; Reach, G.; Dove, C. R.; Lachgar, K.; Grotemeyer, D.; Renton, S. C.  Diabetes Care Mar 2020;43(3):616-624 | Industry Trial in a journal with IF< 30 |
| Moulin 2006 Efficacy of benfluorex in combination with sulfonylurea in type 2 diabetic patients: an 18-week, randomized, double-blind study Moulin, P.; Andre, M.; Alawi, H.; dos Santos, L. C.; Khalid, A. K.; Koev, D.; Moore, R.; Serban, V.; Picandet, B.; Francillard, M.  Diabetes Care Mar 2006;29(3):515-20 | Industry Trial in a journal with IF< 30 |
| Ritzel 2018 A Randomized Controlled Trial Comparing Efficacy and Safety of Insulin Glargine 300 Units/mL Versus 100 Units/mL in Older People With Type 2 Diabetes: Results From the SENIOR Study Ritzel, R.; Harris, S. B.; Baron, H.; Florez, H.; Roussel, R.; Espinasse, M.; Muehlen-Bartmer, I.; Zhang, N.; Bertolini, M.; Brulle-Wohlhueter, C.; Munshi, M.; Bolli, G. B.  Diabetes Care Aug 2018;41(8):1672-1680 | Industry Trial in a journal with IF< 30 |
| Kahn 2010 Rosiglitazone decreases C-reactive protein to a greater extent relative to glyburide and metformin over 4 years despite greater weight gain: observations from a Diabetes Outcome Progression Trial (ADOPT) Kahn, S. E.; Haffner, S. M.; Viberti, G.; Herman, W. H.; Lachin, J. M.; Kravitz, B. G.; Yu, D.; Paul, G.; Holman, R. R.; Zinman, B.  Diabetes Care Jan 2010;33(1):177-83 | Industry Trial in a journal with IF< 30 |
| Panelo 2005 Repaglinide/bedtime NPH insulin is comparable to twice-daily NPH insulin Panelo, A.; Wing, J. R.  Diabetes Care Jul 2005;28(7):1789-90 | Industry Trial in a journal with IF< 30 |
| Riddle 2013 Adding once-daily lixisenatide for type 2 diabetes inadequately controlled by established basal insulin: a 24-week, randomized, placebo-controlled comparison (GetGoal-L) Riddle, M. C.; Aronson, R.; Home, P.; Marre, M.; Niemoeller, E.; Miossec, P.; Ping, L.; Ye, J.; Rosenstock, J.  Diabetes Care Sep 2013;36(9):2489-96 | Industry Trial in a journal with IF< 30 |
| Hermansen 2006 A 26-week, randomized, parallel, treat-to-target trial comparing insulin detemir with NPH insulin as add-on therapy to oral glucose-lowering drugs in insulin-naive people with type 2 diabetes Hermansen, K.; Davies, M.; Derezinski, T.; Martinez Ravn, G.; Clauson, P.; Home, P.  Diabetes Care Jun 2006;29(6):1269-74 | Industry Trial in a journal with IF< 30 |
| Heine 2005 Exenatide versus insulin glargine in patients with suboptimally controlled type 2 diabetes: a randomized trial Heine, R. J.; Van Gaal, L. F.; Johns, D.; Mihm, M. J.; Widel, M. H.; Brodows, R. G.  Ann Intern Med Oct 18 2005;143(8):559-69 | Industry Trial in a journal with IF< 30 |
| Giorgino 2015 Efficacy and Safety of Once-Weekly Dulaglutide Versus Insulin Glargine in Patients With Type 2 Diabetes on Metformin and Glimepiride (AWARD-2) Giorgino, F.; Benroubi, M.; Sun, J. H.; Zimmermann, A. G.; Pechtner, V.  Diabetes Care Dec 2015;38(12):2241-9 | Industry Trial in a journal with IF< 30 |
| Nauck 2014 Efficacy and safety of dulaglutide versus sitagliptin after 52 weeks in type 2 diabetes in a randomized controlled trial (AWARD-5) Nauck, M.; Weinstock, R. S.; Umpierrez, G. E.; Guerci, B.; Skrivanek, Z.; Milicevic, Z.  Diabetes Care Aug 2014;37(8):2149-58 | Industry Trial in a journal with IF< 30 |
| Rosenstock 2005 Inhaled insulin improves glycemic control when substituted for or added to oral combination therapy in type 2 diabetes: a randomized, controlled trial Rosenstock, J.; Zinman, B.; Murphy, L. J.; Clement, S. C.; Moore, P.; Bowering, C. K.; Hendler, R.; Lan, S. P.; Cefalu, W. T.  Ann Intern Med Oct 18 2005;143(8):549-58 | Industry Trial in a journal with IF< 30 |
| Ferdinand 2019 Antihyperglycemic and Blood Pressure Effects of Empagliflozin in Black Patients With Type 2 Diabetes Mellitus and Hypertension Ferdinand, K. C.; Izzo, J. L.; Lee, J.; Meng, L.; George, J.; Salsali, A.; Seman, L.  Circulation Apr 30 2019;139(18):2098-2109 | Industry Trial in a journal with IF< 30 |
| Bilous 2009 Effect of candesartan on microalbuminuria and albumin excretion rate in diabetes: three randomized trials Bilous, R.; Chaturvedi, N.; Sjølie, A. K.; Fuller, J.; Klein, R.; Orchard, T.; Porta, M.; Parving, H. H.  Ann Intern Med Jul 7 2009;151(1):11-20, w3-4 | Industry Trial in a journal with IF< 30 |
| Fonseca 2003 Addition of nateglinide to rosiglitazone monotherapy suppresses mealtime hyperglycemia and improves overall glycemic control Fonseca, V.; Grunberger, G.; Gupta, S.; Shen, S.; Foley, J. E.  Diabetes Care Jun 2003;26(6):1685-90 | Industry Trial in a journal with IF< 30 |
| Viberti 2002 Microalbuminuria reduction with valsartan in patients with type 2 diabetes mellitus: a blood pressure-independent effect Viberti, G.; Wheeldon, N. M.  Circulation Aug 6 2002;106(6):672-8 | Industry Trial in a journal with IF< 30 |
| Rosenstock 2010 Initial combination therapy with alogliptin and pioglitazone in drug-naïve patients with type 2 diabetes Rosenstock, J.; Inzucchi, S. E.; Seufert, J.; Fleck, P. R.; Wilson, C. A.; Mekki, Q.  Diabetes Care Nov 2010;33(11):2406-8 | Industry Trial in a journal with IF< 30 |
| Matthaei 2015 Dapagliflozin improves glycemic control and reduces body weight as add-on therapy to metformin plus sulfonylurea: a 24-week randomized, double-blind clinical trial Matthaei, S.; Bowering, K.; Rohwedder, K.; Grohl, A.; Parikh, S.  Diabetes Care Mar 2015;38(3):365-72 | Industry Trial in a journal with IF< 30 |
| Berl 2003 Cardiovascular outcomes in the Irbesartan Diabetic Nephropathy Trial of patients with type 2 diabetes and overt nephropathy Berl, T.; Hunsicker, L. G.; Lewis, J. B.; Pfeffer, M. A.; Porush, J. G.; Rouleau, J. L.; Drury, P. L.; Esmatjes, E.; Hricik, D.; Parikh, C. R.; Raz, I.; Vanhille, P.; Wiegmann, T. B.; Wolfe, B. M.; Locatelli, F.; Goldhaber, S. Z.; Lewis, E. J.  Ann Intern Med Apr 1 2003;138(7):542-9 | Industry Trial in a journal with IF< 30 |
| Rosenstock 2012 Dose-ranging effects of canagliflozin, a sodium-glucose cotransporter 2 inhibitor, as add-on to metformin in subjects with type 2 diabetes Rosenstock, J.; Aggarwal, N.; Polidori, D.; Zhao, Y.; Arbit, D.; Usiskin, K.; Capuano, G.; Canovatchel, W.  Diabetes Care Jun 2012;35(6):1232-8 | Industry Trial in a journal with IF< 30 |
| Fouqueray 2014 The efficacy and safety of imeglimin as add-on therapy in patients with type 2 diabetes inadequately controlled with sitagliptin monotherapy Fouqueray, P.; Pirags, V.; Diamant, M.; Schernthaner, G.; Lebovitz, H. E.; Inzucchi, S. E.; Bailey, C. J.  Diabetes Care Jul 2014;37(7):1924-30 | Industry Trial in a journal with IF< 30 |
| Diamant 2012 Safety and efficacy of once-weekly exenatide compared with insulin glargine titrated to target in patients with type 2 diabetes over 84 weeks Diamant, M.; Van Gaal, L.; Stranks, S.; Guerci, B.; MacConell, L.; Haber, H.; Scism-Bacon, J.; Trautmann, M.  Diabetes Care Apr 2012;35(4):683-9 | Industry Trial in a journal with IF< 30 |
| Rodbard 2019 Oral Semaglutide Versus Empagliflozin in Patients With Type 2 Diabetes Uncontrolled on Metformin: The PIONEER 2 Trial Rodbard, H. W.; Rosenstock, J.; Canani, L. H.; Deerochanawong, C.; Gumprecht, J.; Lindberg, SØ; Lingvay, I.; Søndergaard, A. L.; Treppendahl, M. B.; Montanya, E.  Diabetes Care Dec 2019;42(12):2272-2281 | Industry Trial in a journal with IF< 30 |
| Erdmann 2007 Pioglitazone use and heart failure in patients with type 2 diabetes and preexisting cardiovascular disease: data from the PROactive study (PROactive 08) Erdmann, E.; Charbonnel, B.; Wilcox, R. G.; Skene, A. M.; Massi-Benedetti, M.; Yates, J.; Tan, M.; Spanheimer, R.; Standl, E.; Dormandy, J. A.  Diabetes Care Nov 2007;30(11):2773-8 | Industry Trial in a journal with IF< 30 |
| Cefalu 2015 Dapagliflozin's Effects on Glycemia and Cardiovascular Risk Factors in High-Risk Patients With Type 2 Diabetes: A 24-Week, Multicenter, Randomized, Double-Blind, Placebo-Controlled Study With a 28-Week Extension Cefalu, W. T.; Leiter, L. A.; de Bruin, T. W.; Gause-Nilsson, I.; Sugg, J.; Parikh, S. J.  Diabetes Care Jul 2015;38(7):1218-27 | Industry Trial in a journal with IF< 30 |
| Striepe 2017 Effects of the Selective Sodium-Glucose Cotransporter 2 Inhibitor Empagliflozin on Vascular Function and Central Hemodynamics in Patients With Type 2 Diabetes Mellitus Striepe, K.; Jumar, A.; Ott, C.; Karg, M. V.; Schneider, M. P.; Kannenkeril, D.; Schmieder, R. E.  Circulation Sep 19 2017;136(12):1167-1169 | Industry Trial in a journal with IF< 30 |
| Haffner 2002 Effect of rosiglitazone treatment on nontraditional markers of cardiovascular disease in patients with type 2 diabetes mellitus Haffner, S. M.; Greenberg, A. S.; Weston, W. M.; Chen, H.; Williams, K.; Freed, M. I.  Circulation Aug 6 2002;106(6):679-84 | Industry Trial in a journal with IF< 30 |
| Yki-Järvinen 2014 New insulin glargine 300 units/mL versus glargine 100 units/mL in people with type 2 diabetes using oral agents and basal insulin: glucose control and hypoglycemia in a 6-month randomized controlled trial (EDITION 2) Yki-Järvinen, H.; Bergenstal, R.; Ziemen, M.; Wardecki, M.; Muehlen-Bartmer, I.; Boelle, E.; Riddle, M. C.  Diabetes Care Dec 2014;37(12):3235-43 | Industry Trial in a journal with IF< 30 |
| vonEynatten 2005 Adipocytokines as a novel target for the anti-inflammatory effect of atorvastatin in patients with type 2 diabetes von Eynatten, M.; Schneider, J. G.; Hadziselimovic, S.; Hamann, A.; Bierhaus, A.; Nawroth, P. P.; Dugi, K. A.  Diabetes Care Mar 2005;28(3):754-5 | Industry Trial in a journal with IF< 30 |
| #99 Cardiovascular and Other Outcomes Postintervention With Insulin Glargine and Omega-3 Fatty Acids (ORIGINALE) Diabetes Care May 2016;39(5):709-16 | Industry Trial in a journal with IF< 30 |
| Gottschalk 2007 Glimepiride versus metformin as monotherapy in pediatric patients with type 2 diabetes: a randomized, single-blind comparative study Gottschalk, M.; Danne, T.; Vlajnic, A.; Cara, J. F.  Diabetes Care Apr 2007;30(4):790-4 | Industry Trial in a journal with IF< 30 |
| Best 2011 Weight-related quality of life, health utility, psychological well-being, and satisfaction with exenatide once weekly compared with sitagliptin or pioglitazone after 26 weeks of treatment Best, J. H.; Rubin, R. R.; Peyrot, M.; Li, Y.; Yan, P.; Malloy, J.; Garrison, L. P.  Diabetes Care Feb 2011;34(2):314-9 | Industry Trial in a journal with IF< 30 |
| Siegelaar 2011 A decrease in glucose variability does not reduce cardiovascular event rates in type 2 diabetic patients after acute myocardial infarction: a reanalysis of the HEART2D study Siegelaar, S. E.; Kerr, L.; Jacober, S. J.; Devries, J. H.  Diabetes Care Apr 2011;34(4):855-7 | Industry Trial in a journal with IF< 30 |
| Hollander 2003 Pramlintide as an adjunct to insulin therapy improves long-term glycemic and weight control in patients with type 2 diabetes: a 1-year randomized controlled trial Hollander, P. A.; Levy, P.; Fineman, M. S.; Maggs, D. G.; Shen, L. Z.; Strobel, S. A.; Weyer, C.; Kolterman, O. G.  Diabetes Care Mar 2003;26(3):784-90 | Industry Trial in a journal with IF< 30 |
| Gaziano 2010 Randomized clinical trial of quick-release bromocriptine among patients with type 2 diabetes on overall safety and cardiovascular outcomes Gaziano, J. M.; Cincotta, A. H.; O'Connor, C. M.; Ezrokhi, M.; Rutty, D.; Ma, Z. J.; Scranton, R. E.  Diabetes Care Jul 2010;33(7):1503-8 | Industry Trial in a journal with IF< 30 |
| Estacio 2000 Effect of blood pressure control on diabetic microvascular complications in patients with hypertension and type 2 diabetes Estacio, R. O.; Jeffers, B. W.; Gifford, N.; Schrier, R. W.  Diabetes Care Apr 2000;23 Suppl 2():B54-64  Upload full text Abstract 3 Notes History Duplicate Move study to Full text review | Industry Trial in a journal with IF< 30 |
| Dailey 2004 Insulin glulisine provides improved glycemic control in patients with type 2 diabetes Dailey, G.; Rosenstock, J.; Moses, R. G.; Ways, K.  Diabetes Care Oct 2004;27(10):2363-8 | Industry Trial in a journal with IF< 30 |
| Fritsche 2003 Glimepiride combined with morning insulin glargine, bedtime neutral protamine hagedorn insulin, or bedtime insulin glargine in patients with type 2 diabetes. A randomized, controlled trial Fritsche, A.; Schweitzer, M. A.; Häring, H. U.  Ann Intern Med Jun 17 2003;138(12):952-9 | Industry Trial in a journal with IF< 30 |
| Langenfeld 2005 Pioglitazone decreases carotid intima-media thickness independently of glycemic control in patients with type 2 diabetes mellitus: results from a controlled randomized study Langenfeld, M. R.; Forst, T.; Hohberg, C.; Kann, P.; Lübben, G.; Konrad, T.; Füllert, S. D.; Sachara, C.; Pfützner, A.  Circulation May 17 2005;111(19):2525-31 | Industry Trial in a journal with IF< 30 |
| Hodis 2006 Effect of peroxisome proliferator-activated receptor gamma agonist treatment on subclinical atherosclerosis in patients with insulin-requiring type 2 diabetes Hodis, H. N.; Mack, W. J.; Zheng, L.; Li, Y.; Torres, M.; Sevilla, D.; Stewart, Y.; Hollen, B.; Garcia, K.; Alaupovic, P.; Buchanan, T. A.  Diabetes Care Jul 2006;29(7):1545-53 | Industry Trial in a journal with IF< 30 |
| Umpierrez 2013 Randomized study comparing a Basal-bolus with a basal plus correction insulin regimen for the hospital management of medical and surgical patients with type 2 diabetes: basal plus trial Umpierrez, G. E.; Smiley, D.; Hermayer, K.; Khan, A.; Olson, D. E.; Newton, C.; Jacobs, S.; Rizzo, M.; Peng, L.; Reyes, D.; Pinzon, I.; Fereira, M. E.; Hunt, V.; Gore, A.; Toyoshima, M. T.; Fonseca, V. A.  Diabetes Care Aug 2013;36(8):2169-74 | Industry Trial in a journal with IF< 30 |
| Rosenstock 2001 Basal insulin therapy in type 2 diabetes: 28-week comparison of insulin glargine (HOE 901) and NPH insulin Rosenstock, J.; Schwartz, S. L.; Clark, C. M., Jr.; Park, G. D.; Donley, D. W.; Edwards, M. B.  Diabetes Care Apr 2001;24(4):631-6 | Industry Trial in a journal with IF< 30 |
| StJohnSutton 2002 A comparison of the effects of rosiglitazone and glyburide on cardiovascular function and glycemic control in patients with type 2 diabetes St John Sutton, M.; Rendell, M.; Dandona, P.; Dole, J. F.; Murphy, K.; Patwardhan, R.; Patel, J.; Freed, M.  Diabetes Care Nov 2002;25(11):2058-64 | Industry Trial in a journal with IF< 30 |
| Mita 2016 Sitagliptin Attenuates the Progression of Carotid Intima-Media Thickening in Insulin-Treated Patients With Type 2 Diabetes: The Sitagliptin Preventive Study of Intima-Media Thickness Evaluation (SPIKE): A Randomized Controlled Trial Mita, T.; Katakami, N.; Shiraiwa, T.; Yoshii, H.; Onuma, T.; Kuribayashi, N.; Osonoi, T.; Kaneto, H.; Kosugi, K.; Umayahara, Y.; Yamamoto, T.; Matsumoto, K.; Yokoyama, H.; Tsugawa, M.; Gosho, M.; Shimomura, I.; Watada, H.  Diabetes Care Mar 2016;39(3):455-64 | Industry Trial in a journal with IF< 30 |
| Phillips 2001 Once- and twice-daily dosing with rosiglitazone improves glycemic control in patients with type 2 diabetes Phillips, L. S.; Grunberger, G.; Miller, E.; Patwardhan, R.; Rappaport, E. B.; Salzman, A.  Diabetes Care Feb 2001;24(2):308-15 | Industry Trial in a journal with IF< 30 |
| Saad 2004 Ragaglitazar improves glycemic control and lipid profile in type 2 diabetic subjects: a 12-week, double-blind, placebo-controlled dose-ranging study with an open pioglitazone arm Saad, M. F.; Greco, S.; Osei, K.; Lewin, A. J.; Edwards, C.; Nunez, M.; Reinhardt, R. R.  Diabetes Care Jun 2004;27(6):1324-9 | Industry Trial in a journal with IF< 30 |
| Raskin 2001 A randomized trial of rosiglitazone therapy in patients with inadequately controlled insulin-treated type 2 diabetes Raskin, P.; Rendell, M.; Riddle, M. C.; Dole, J. F.; Freed, M. I.; Rosenstock, J.  Diabetes Care Jul 2001;24(7):1226-32 | Industry Trial in a journal with IF< 30 |
| Ryan 2006 Improving metabolic control leads to better working memory in adults with type 2 diabetes Ryan, C. M.; Freed, M. I.; Rood, J. A.; Cobitz, A. R.; Waterhouse, B. R.; Strachan, M. W.  Diabetes Care Feb 2006;29(2):345-51 | Industry Trial in a journal with IF< 30 |
| Yale 2001 The effect of a thiazolidinedione drug, troglitazone, on glycemia in patients with type 2 diabetes mellitus poorly controlled with sulfonylurea and metformin. A multicenter, randomized, double-blind, placebo-controlled trial Yale, J. F.; Valiquett, T. R.; Ghazzi, M. N.; Owens-Grillo, J. K.; Whitcomb, R. W.; Foyt, H. L.  Ann Intern Med May 1 2001;134(9 Pt 1):737-45 | Industry Trial in a journal with IF< 30 |
| Sandercock 2009 Gabapentin extended release for the treatment of painful diabetic peripheral neuropathy: efficacy and tolerability in a double-blind, randomized, controlled clinical trial Sandercock, D.; Cramer, M.; Wu, J.; Chiang, Y. K.; Biton, V.; Heritier, M.  Diabetes Care Feb 2009;32(2):e20 | Industry Trial in a journal with IF< 30 |
| Raskin 2003 Continuous subcutaneous insulin infusion and multiple daily injection therapy are equally effective in type 2 diabetes: a randomized, parallel-group, 24-week study Raskin, P.; Bode, B. W.; Marks, J. B.; Hirsch, I. B.; Weinstein, R. L.; McGill, J. B.; Peterson, G. E.; Mudaliar, S. R.; Reinhardt, R. R.  Diabetes Care Sep 2003;26(9):2598-603 | Industry Trial in a journal with IF< 30 |
| Buse 2004 Effects of exenatide (exendin-4) on glycemic control over 30 weeks in sulfonylurea-treated patients with type 2 diabetes Buse, J. B.; Henry, R. R.; Han, J.; Kim, D. D.; Fineman, M. S.; Baron, A. D.  Diabetes Care Nov 2004;27(11):2628-35 | Industry Trial in a journal with IF< 30 |
| Zandbergen 2003 Effect of losartan on microalbuminuria in normotensive patients with type 2 diabetes mellitus. A randomized clinical trial Zandbergen, A. A.; Baggen, M. G.; Lamberts, S. W.; Bootsma, A. H.; de Zeeuw, D.; Ouwendijk, R. J.  Ann Intern Med Jul 15 2003;139(2):90-6 | Industry Trial in a journal with IF< 30 |
| Beishuizen 2005 No effect of statin therapy on silent myocardial ischemia in patients with type 2 diabetes without manifest cardiovascular disease Beishuizen, E. D.; Jukema, J. W.; Tamsma, J. T.; van de Ree, M. A.; van der Vijver, J. C.; Putter, H.; Maan, A. C.; Meinders, A. E.; Huisman, M. V.  Diabetes Care Jul 2005;28(7):1675-9 | Industry Trial in a journal with IF< 30 |
| Sloan-Lancaster 2013 Double-blind, randomized study evaluating the glycemic and anti-inflammatory effects of subcutaneous LY2189102, a neutralizing IL-1β antibody, in patients with type 2 diabetes Sloan-Lancaster, J.; Abu-Raddad, E.; Polzer, J.; Miller, J. W.; Scherer, J. C.; De Gaetano, A.; Berg, J. K.; Landschulz, W. H.  Diabetes Care Aug 2013;36(8):2239-46 | Industry Trial in a journal with IF< 30 |
| Wulffelé 2002 Combination of insulin and metformin in the treatment of type 2 diabetes Wulffelé, M. G.; Kooy, A.; Lehert, P.; Bets, D.; Ogterop, J. C.; Borger van der Burg, B.; Donker, A. J.; Stehouwer, C. D.  Diabetes Care Dec 2002;25(12):2133-40 | Industry Trial in a journal with IF< 30 |
| Chiasson 2001 The synergistic effect of miglitol plus metformin combination therapy in the treatment of type 2 diabetes Chiasson, J. L.; Naditch, L.  Diabetes Care Jun 2001;24(6):989-94 | Industry Trial in a journal with IF< 30 |
| Wysham 2016 Efficacy and Safety of Multiple Doses of Exenatide Once-Monthly Suspension in Patients With Type 2 Diabetes: A Phase II Randomized Clinical Trial Wysham, C. H.; MacConell, L.; Hardy, E.  Diabetes Care Oct 2016;39(10):1768-76 | Industry Trial in a journal with IF< 30 |
| Yang 2008 Biphasic insulin aspart 30 three times daily is more effective than a twice-daily regimen, without increasing hypoglycemia, in Chinese subjects with type 2 diabetes inadequately controlled on oral antidiabetes drugs Yang, W.; Ji, Q.; Zhu, D.; Yang, J.; Chen, L.; Liu, Z.; Yu, D.; Yan, L.  Diabetes Care May 2008;31(5):852-6 | Industry Trial in a journal with IF< 30 |
| Cleveringa 2008 Combined task delegation, computerized decision support, and feedback improve cardiovascular risk for type 2 diabetic patients: a cluster randomized trial in primary care Cleveringa, F. G.; Gorter, K. J.; van den Donk, M.; Rutten, G. E.  Diabetes Care Dec 2008;31(12):2273-5 | Industry Trial in a journal with IF< 30 |
| Young 2005 Pro-active call center treatment support (PACCTS) to improve glucose control in type 2 diabetes: a randomized controlled trial Young, R. J.; Taylor, J.; Friede, T.; Hollis, S.; Mason, J. M.; Lee, P.; Burns, E.; Long, A. F.; Gambling, T.; New, J. P.; Gibson, J. M.  Diabetes Care Feb 2005;28(2):278-82 | Industry Trial in a journal with IF< 30 |
| Gallwitz 2011 Exenatide twice daily versus premixed insulin aspart 70/30 in metformin-treated patients with type 2 diabetes: a randomized 26-week study on glycemic control and hypoglycemia Gallwitz, B.; Böhmer, M.; Segiet, T.; Mölle, A.; Milek, K.; Becker, B.; Helsberg, K.; Petto, H.; Peters, N.; Bachmann, O.  Diabetes Care Mar 2011;34(3):604-6 | Industry Trial in a journal with IF< 30 |
| Rosenstock 2004 Repaglinide versus nateglinide monotherapy: a randomized, multicenter study Rosenstock, J.; Hassman, D. R.; Madder, R. D.; Brazinsky, S. A.; Farrell, J.; Khutoryansky, N.; Hale, P. M.  Diabetes Care Jun 2004;27(6):1265-70 | Industry Trial in a journal with IF< 30 |
| Kipnes 2003 Control of postprandial plasma glucose by an oral insulin product (HIM2) in patients with type 2 diabetes Kipnes, M.; Dandona, P.; Tripathy, D.; Still, J. G.; Kosutic, G.  Diabetes Care Feb 2003;26(2):421-6 | Industry Trial in a journal with IF< 30 |
| Rosenstock 2015 Greater dose-ranging effects on A1C levels than on glucosuria with LX4211, a dual inhibitor of SGLT1 and SGLT2, in patients with type 2 diabetes on metformin monotherapy Rosenstock, J.; Cefalu, W. T.; Lapuerta, P.; Zambrowicz, B.; Ogbaa, I.; Banks, P.; Sands, A.  Diabetes Care Mar 2015;38(3):431-8 | Industry Trial in a journal with IF< 30 |
| Mita 2016 Alogliptin, a Dipeptidyl Peptidase 4 Inhibitor, Prevents the Progression of Carotid Atherosclerosis in Patients With Type 2 Diabetes: The Study of Preventive Effects of Alogliptin on Diabetic Atherosclerosis (SPEAD-A) Mita, T.; Katakami, N.; Yoshii, H.; Onuma, T.; Kaneto, H.; Osonoi, T.; Shiraiwa, T.; Kosugi, K.; Umayahara, Y.; Yamamoto, T.; Yokoyama, H.; Kuribayashi, N.; Jinnouchi, H.; Gosho, M.; Shimomura, I.; Watada, H.  Diabetes Care Jan 2016;39(1):139-48 | Industry Trial in a journal with IF< 30 |
| Kelley 2002 Clinical efficacy of orlistat therapy in overweight and obese patients with insulin-treated type 2 diabetes: A 1-year randomized controlled trial Kelley, D. E.; Bray, G. A.; Pi-Sunyer, F. X.; Klein, S.; Hill, J.; Miles, J.; Hollander, P.  Diabetes Care Jun 2002;25(6):1033-41 | Industry Trial in a journal with IF< 30 |
| delaPeña 2011 Pharmacokinetics and pharmacodynamics of high-dose human regular U-500 insulin versus human regular U-100 insulin in healthy obese subjects de la Peña, A.; Riddle, M.; Morrow, L. A.; Jiang, H. H.; Linnebjerg, H.; Scott, A.; Win, K. M.; Hompesch, M.; Mace, K. F.; Jacobson, J. G.; Jackson, J. A.  Diabetes Care Dec 2011;34(12):2496-501 | Industry Trial in a journal with IF< 30 |
| Kendall 2005 Effects of exenatide (exendin-4) on glycemic control over 30 weeks in patients with type 2 diabetes treated with metformin and a sulfonylurea Kendall, D. M.; Riddle, M. C.; Rosenstock, J.; Zhuang, D.; Kim, D. D.; Fineman, M. S.; Baron, A. D.  Diabetes Care May 2005;28(5):1083-91 | Industry Trial in a journal with IF< 30 |
| Rosenstock 2006 Triple therapy in type 2 diabetes: insulin glargine or rosiglitazone added to combination therapy of sulfonylurea plus metformin in insulin-naive patients Rosenstock, J.; Sugimoto, D.; Strange, P.; Stewart, J. A.; Soltes-Rak, E.; Dailey, G.  Diabetes Care Mar 2006;29(3):554-9 | Industry Trial in a journal with IF< 30 |
| Henry 2013 Randomized trial of continuous subcutaneous delivery of exenatide by ITCA 650 versus twice-daily exenatide injections in metformin-treated type 2 diabetes Henry, R. R.; Rosenstock, J.; Logan, D. K.; Alessi, T. R.; Luskey, K.; Baron, M. A.  Diabetes Care Sep 2013;36(9):2559-65 | Industry Trial in a journal with IF< 30 |
| Idorn 2016 Safety and Efficacy of Liraglutide in Patients With Type 2 Diabetes and End-Stage Renal Disease: An Investigator-Initiated, Placebo-Controlled, Double-Blind, Parallel-Group, Randomized Trial Idorn, T.; Knop, F. K.; Jørgensen, M. B.; Jensen, T.; Resuli, M.; Hansen, P. M.; Christensen, K. B.; Holst, J. J.; Hornum, M.; Feldt-Rasmussen, B.  Diabetes Care Feb 2016;39(2):206-13 | Industry Trial in a journal with IF< 30 |
| Sfikakis 2010 Infliximab for diabetic macular edema refractory to laser photocoagulation: a randomized, double-blind, placebo-controlled, crossover, 32-week study Sfikakis, P. P.; Grigoropoulos, V.; Emfietzoglou, I.; Theodossiadis, G.; Tentolouris, N.; Delicha, E.; Katsiari, C.; Alexiadou, K.; Hatziagelaki, E.; Theodossiadis, P. G.  Diabetes Care Jul 2010;33(7):1523-8 | Industry Trial in a journal with IF< 30 |
| Umpierrez 2007 Randomized study of basal-bolus insulin therapy in the inpatient management of patients with type 2 diabetes (RABBIT 2 trial) Umpierrez, G. E.; Smiley, D.; Zisman, A.; Prieto, L. M.; Palacio, A.; Ceron, M.; Puig, A.; Mejia, R.  Diabetes Care Sep 2007;30(9):2181-6 | Industry Trial in a journal with IF< 30 |
| Beishuizen 2005 The effect of statin therapy on endothelial function in type 2 diabetes without manifest cardiovascular disease Beishuizen, E. D.; Tamsma, J. T.; Jukema, J. W.; van de Ree, M. A.; van der Vijver, J. C.; Meinders, A. E.; Huisman, M. V.  Diabetes Care Jul 2005;28(7):1668-74 | Industry Trial in a journal with IF< 30 |
| Fineman 2003 Effect on glycemic control of exenatide (synthetic exendin-4) additive to existing metformin and/or sulfonylurea treatment in patients with type 2 diabetes Fineman, M. S.; Bicsak, T. A.; Shen, L. Z.; Taylor, K.; Gaines, E.; Varns, A.; Kim, D.; Baron, A. D.  Diabetes Care Aug 2003;26(8):2370-7 | Industry Trial in a journal with IF< 30 |
| vanVenrooij 2002 Aggressive lipid lowering does not improve endothelial function in type 2 diabetes: the Diabetes Atorvastatin Lipid Intervention (DALI) Study: a randomized, double-blind, placebo-controlled trial van Venrooij, F. V.; van de Ree, M. A.; Bots, M. L.; Stolk, R. P.; Huisman, M. V.; Banga, J. D.  Diabetes Care Jul 2002;25(7):1211-6 | Industry Trial in a journal with IF< 30 |
| vanVenrooij 2003 Common cholesteryl ester transfer protein gene polymorphisms and the effect of atorvastatin therapy in type 2 diabetes van Venrooij, F. V.; Stolk, R. P.; Banga, J. D.; Sijmonsma, T. P.; van Tol, A.; Erkelens, D. W.; Dallinga-Thie, G. M.  Diabetes Care Apr 2003;26(4):1216-23 | Industry Trial in a journal with IF< 30 |
| Umpierrez 2011 Randomized study of basal-bolus insulin therapy in the inpatient management of patients with type 2 diabetes undergoing general surgery (RABBIT 2 surgery) Umpierrez, G. E.; Smiley, D.; Jacobs, S.; Peng, L.; Temponi, A.; Mulligan, P.; Umpierrez, D.; Newton, C.; Olson, D.; Rizzo, M.  Diabetes Care Feb 2011;34(2):256-61 | Industry Trial in a journal with IF< 30 |
| Riddle 2009 Randomized comparison of pramlintide or mealtime insulin added to basal insulin treatment for patients with type 2 diabetes Riddle, M.; Pencek, R.; Charenkavanich, S.; Lutz, K.; Wilhelm, K.; Porter, L.  Diabetes Care Sep 2009;32(9):1577-82 | Industry Trial in a journal with IF< 30 |
| Pettus 2020 Efficacy and Safety of the Glucagon Receptor Antagonist RVT-1502 in Type 2 Diabetes Uncontrolled on Metformin Monotherapy: A 12-Week Dose-Ranging Study Pettus, J. H.; D'Alessio, D.; Frias, J. P.; Vajda, E. G.; Pipkin, J. D.; Rosenstock, J.; Williamson, G.; Zangmeister, M. A.; Zhi, L.; Marschke, K. B.  Diabetes Care Jan 2020;43(1):161-168 | Industry Trial in a journal with IF< 30 |
| Bretzel 2004 A direct efficacy and safety comparison of insulin aspart, human soluble insulin, and human premix insulin (70/30) in patients with type 2 diabetes Bretzel, R. G.; Arnolds, S.; Medding, J.; Linn, T.  Diabetes Care May 2004;27(5):1023-7 | Industry Trial in a journal with IF< 30 |
| Bril 2004 Aldose reductase inhibition by AS-3201 in sural nerve from patients with diabetic sensorimotor polyneuropathy Bril, V.; Buchanan, R. A.  Diabetes Care Oct 2004;27(10):2369-75 | Industry Trial in a journal with IF< 30 |
| Vigersky 2012 Short- and long-term effects of real-time continuous glucose monitoring in patients with type 2 diabetes Vigersky, R. A.; Fonda, S. J.; Chellappa, M.; Walker, M. S.; Ehrhardt, N. M.  Diabetes Care Jan 2012;35(1):32-8 | Industry Trial in a journal with IF< 30 |
| #1 The effect of aggressive versus standard lipid lowering by atorvastatin on diabetic dyslipidemia: the DALI study: a double-blind, randomized, placebo-controlled trial in patients with type 2 diabetes and diabetic dyslipidemia Diabetes Care Aug 2001;24(8):1335-41 | Industry Trial in a journal with IF< 30 |
| Ruggenenti 2008 Preventing left ventricular hypertrophy by ACE inhibition in hypertensive patients with type 2 diabetes: a prespecified analysis of the Bergamo Nephrologic Diabetes Complications Trial (BENEDICT) Ruggenenti, P.; Iliev, I.; Costa, G. M.; Parvanova, A.; Perna, A.; Giuliano, G. A.; Motterlini, N.; Ene-Iordache, B.; Remuzzi, G.  Diabetes Care Aug 2008;31(8):1629-34 | Industry Trial in a journal with IF< 30 |
| Goldberg 2005 A comparison of lipid and glycemic effects of pioglitazone and rosiglitazone in patients with type 2 diabetes and dyslipidemia Goldberg, R. B.; Kendall, D. M.; Deeg, M. A.; Buse, J. B.; Zagar, A. J.; Pinaire, J. A.; Tan, M. H.; Khan, M. A.; Perez, A. T.; Jacober, S. J.  Diabetes Care Jul 2005;28(7):1547-54 | Industry Trial in a journal with IF< 30 |
| Bae 2015 Improvement of Nonalcoholic Fatty Liver Disease With Carnitine-Orotate Complex in Type 2 Diabetes (CORONA): A Randomized Controlled Trial Bae, J. C.; Lee, W. Y.; Yoon, K. H.; Park, J. Y.; Son, H. S.; Han, K. A.; Lee, K. W.; Woo, J. T.; Ju, Y. C.; Lee, W. J.; Cho, Y. Y.; Lee, M. K.  Diabetes Care Jul 2015;38(7):1245-52 | Industry Trial in a journal with IF< 30 |
| Vinik 2007 Adding insulin glargine versus rosiglitazone: health-related quality-of-life impact in type 2 diabetes Vinik, A. I.; Zhang, Q.  Diabetes Care Apr 2007;30(4):795-800 | Industry Trial in a journal with IF< 30 |
| Aronoff 2000 Pioglitazone hydrochloride monotherapy improves glycemic control in the treatment of patients with type 2 diabetes: a 6-month randomized placebo-controlled dose-response study. The Pioglitazone 001 Study Group Aronoff, S.; Rosenblatt, S.; Braithwaite, S.; Egan, J. W.; Mathisen, A. L.; Schneider, R. L.  Diabetes Care Nov 2000;23(11):1605-11 | Industry Trial in a journal with IF< 30 |
| Watada 2020 Efficacy and Safety of 1:1 Fixed-Ratio Combination of Insulin Glargine and Lixisenatide Versus Lixisenatide in Japanese Patients With Type 2 Diabetes Inadequately Controlled on Oral Antidiabetic Drugs: The LixiLan JP-O1 Randomized Clinical Trial Watada, H.; Takami, A.; Spranger, R.; Amano, A.; Hashimoto, Y.; Niemoeller, E.  Diabetes Care Jun 2020;43(6):1249-1257 | Industry Trial in a journal with IF< 30 |
| Kennedy 2006 Impact of active versus usual algorithmic titration of basal insulin and point-of-care versus laboratory measurement of HbA1c on glycemic control in patients with type 2 diabetes: the Glycemic Optimization with Algorithms and Labs at Point of Care (GOAL A1C) trial Kennedy, L.; Herman, W. H.; Strange, P.; Harris, A.  Diabetes Care Jan 2006;29(1):1-8 | Industry Trial in a journal with IF< 30 |
| Gerstein 2020 Impact of Acarbose on Incident Diabetes and Regression to Normoglycemia in People With Coronary Heart Disease and Impaired Glucose Tolerance: Insights From the ACE Trial Gerstein, H. C.; Coleman, R. L.; Scott, C. A. B.; Xu, S.; Tuomilehto, J.; Rydén, L.; Holman, R. R.  Diabetes Care Sep 2020;43(9):2242-2247 | Industry Trial in a journal with IF< 30 |
| Rosenstock 2007 A randomized, double-blind, placebo-controlled, multicenter study to assess the efficacy and safety of topiramate controlled release in the treatment of obese type 2 diabetic patients Rosenstock, J.; Hollander, P.; Gadde, K. M.; Sun, X.; Strauss, R.; Leung, A.  Diabetes Care Jun 2007;30(6):1480-6 | Industry Trial in a journal with IF< 30 |
| Itoh 2018 Intensive Treat-to-Target Statin Therapy in High-Risk Japanese Patients With Hypercholesterolemia and Diabetic Retinopathy: Report of a Randomized Study Itoh, H.; Komuro, I.; Takeuchi, M.; Akasaka, T.; Daida, H.; Egashira, Y.; Fujita, H.; Higaki, J.; Hirata, K. I.; Ishibashi, S.; Isshiki, T.; Ito, S.; Kashiwagi, A.; Kato, S.; Kitagawa, K.; Kitakaze, M.; Kitazono, T.; Kurabayashi, M.; Miyauchi, K.; Murakami, T.; Murohara, T.; Node, K.; Ogawa, S.; Saito, Y.; Seino, Y.; Shigeeda, T.; Shindo, S.; Sugawara, M.; Sugiyama, S.; Terauchi, Y.; Tsutsui, H.; Ueshima, K.; Utsunomiya, K.; Yamagishi, M.; Yamazaki, T.; Yo, S.; Yokote, K.; Yoshida, K.; Yoshimura, M.; Yoshimura, N.; Nakao, K.; Nagai, R.  Diabetes Care Jun 2018;41(6):1275-1284 | Industry Trial in a journal with IF< 30 |
| Kendall 2006 Improvement of glycemic control, triglycerides, and HDL cholesterol levels with muraglitazar, a dual (alpha/gamma) peroxisome proliferator-activated receptor activator, in patients with type 2 diabetes inadequately controlled with metformin monotherapy: A double-blind, randomized, pioglitazone-comparative study Kendall, D. M.; Rubin, C. J.; Mohideen, P.; Ledeine, J. M.; Belder, R.; Gross, J.; Norwood, P.; O'Mahony, M.; Sall, K.; Sloan, G.; Roberts, A.; Fiedorek, F. T.; DeFronzo, R. A.  Diabetes Care May 2006;29(5):1016-23 | Industry Trial in a journal with IF< 30 |
| Schwartz 2006 Efficacy, tolerability, and safety of a novel once-daily extended-release metformin in patients with type 2 diabetes Schwartz, S.; Fonseca, V.; Berner, B.; Cramer, M.; Chiang, Y. K.; Lewin, A.  Diabetes Care Apr 2006;29(4):759-64 | Industry Trial in a journal with IF< 30 |
| Schwartz 2003 Insulin 70/30 mix plus metformin versus triple oral therapy in the treatment of type 2 diabetes after failure of two oral drugs: efficacy, safety, and cost analysis Schwartz, S.; Sievers, R.; Strange, P.; Lyness, W. H.; Hollander, P.  Diabetes Care Aug 2003;26(8):2238-43 | Industry Trial in a journal with IF< 30 |
| Raskin 2005 Initiating insulin therapy in type 2 Diabetes: a comparison of biphasic and basal insulin analogs Raskin, P.; Allen, E.; Hollander, P.; Lewin, A.; Gabbay, R. A.; Hu, P.; Bode, B.; Garber, A.  Diabetes Care Feb 2005;28(2):260-5 | Industry Trial in a journal with IF< 30 |
| Sacco 2003 Primary prevention of cardiovascular events with low-dose aspirin and vitamin E in type 2 diabetic patients: results of the Primary Prevention Project (PPP) trial Sacco, M.; Pellegrini, F.; Roncaglioni, M. C.; Avanzini, F.; Tognoni, G.; Nicolucci, A.  Diabetes Care Dec 2003;26(12):3264-72 | Industry Trial in a journal with IF< 30 |
| Henry 2014 Basal insulin peglispro demonstrates preferential hepatic versus peripheral action relative to insulin glargine in healthy subjects Henry, R. R.; Mudaliar, S.; Ciaraldi, T. P.; Armstrong, D. A.; Burke, P.; Pettus, J.; Garhyan, P.; Choi, S. L.; Jacober, S. J.; Knadler, M. P.; Lam, E. C.; Prince, M. J.; Bose, N.; Porksen, N.; Sinha, V. P.; Linnebjerg, H.  Diabetes Care Sep 2014;37(9):2609-15 | Industry Trial in a journal with IF< 30 |
| Rosenstock 2018 Efficacy and Safety of ITCA 650, a Novel Drug-Device GLP-1 Receptor Agonist, in Type 2 Diabetes Uncontrolled With Oral Antidiabetes Drugs: The FREEDOM-1 Trial Rosenstock, J.; Buse, J. B.; Azeem, R.; Prabhakar, P.; Kjems, L.; Huang, H.; Baron, M. A.  Diabetes Care Feb 2018;41(2):333-340 | Industry Trial in a journal with IF< 30 |
| Araki 2018 Effects of Pemafibrate, a Novel Selective PPARα Modulator, on Lipid and Glucose Metabolism in Patients With Type 2 Diabetes and Hypertriglyceridemia: A Randomized, Double-Blind, Placebo-Controlled, Phase 3 Trial Araki, E.; Yamashita, S.; Arai, H.; Yokote, K.; Satoh, J.; Inoguchi, T.; Nakamura, J.; Maegawa, H.; Yoshioka, N.; Tanizawa, Y.; Watada, H.; Suganami, H.; Ishibashi, S.  Diabetes Care Mar 2018;41(3):538-546 | Industry Trial in a journal with IF< 30 |
| Baldwin 2012 A randomized trial of two weight-based doses of insulin glargine and glulisine in hospitalized subjects with type 2 diabetes and renal insufficiency Baldwin, D.; Zander, J.; Munoz, C.; Raghu, P.; DeLange-Hudec, S.; Lee, H.; Emanuele, M. A.; Glossop, V.; Smallwood, K.; Molitch, M.  Diabetes Care Oct 2012;35(10):1970-4 | Industry Trial in a journal with IF< 30 |
| Cryer 2005 Comparative outcomes study of metformin intervention versus conventional approach the COSMIC Approach Study Cryer, D. R.; Nicholas, S. P.; Henry, D. H.; Mills, D. J.; Stadel, B. V.  Diabetes Care Mar 2005;28(3):539-43 | Industry Trial in a journal with IF< 30 |
| Rosenstock 2002 Combination therapy with nateglinide and a thiazolidinedione improves glycemic control in type 2 diabetes Rosenstock, J.; Shen, S. G.; Gatlin, M. R.; Foley, J. E.  Diabetes Care Sep 2002;25(9):1529-33 | Industry Trial in a journal with IF< 30 |
| Ruggenenti 2010 Effects of combined ezetimibe and simvastatin therapy as compared with simvastatin alone in patients with type 2 diabetes: a prospective randomized double-blind clinical trial Ruggenenti, P.; Cattaneo, D.; Rota, S.; Iliev, I.; Parvanova, A.; Diadei, O.; Ene-Iordache, B.; Ferrari, S.; Bossi, A. C.; Trevisan, R.; Belviso, A.; Remuzzi, G.  Diabetes Care Sep 2010;33(9):1954-6 | Industry Trial in a journal with IF< 30 |
| Rosenstock 2008 Advancing insulin therapy in type 2 diabetes previously treated with glargine plus oral agents: prandial premixed (insulin lispro protamine suspension/lispro) versus basal/bolus (glargine/lispro) therapy Rosenstock, J.; Ahmann, A. J.; Colon, G.; Scism-Bacon, J.; Jiang, H.; Martin, S.  Diabetes Care Jan 2008;31(1):20-5 | Industry Trial in a journal with IF< 30 |
| Horton 2000 Nateglinide alone and in combination with metformin improves glycemic control by reducing mealtime glucose levels in type 2 diabetes Horton, E. S.; Clinkingbeard, C.; Gatlin, M.; Foley, J.; Mallows, S.; Shen, S.  Diabetes Care Nov 2000;23(11):1660-5 | Industry Trial in a journal with IF< 30 |
| Berk 2003 Atorvastatin dose-dependently decreases hepatic lipase activity in type 2 diabetes: effect of sex and the LIPC promoter variant Berk, Planken, II; Hoogerbrugge, N.; Stolk, R. P.; Bootsma, A. H.; Jansen, H.  Diabetes Care Feb 2003;26(2):427-32 | Industry Trial in a journal with IF< 30 |
| deJager 2010 Long term treatment with metformin in patients with type 2 diabetes and risk of vitamin B-12 deficiency: randomised placebo controlled trial de Jager, J.; Kooy, A.; Lehert, P.; Wulffelé, M. G.; van der Kolk, J.; Bets, D.; Verburg, J.; Donker, A. J.; Stehouwer, C. D.  Bmj May 20 2010;340():c2181 | Industry Trial in a journal with IF< 30 |
| Hollander 2001 Importance of early insulin secretion: comparison of nateglinide and glyburide in previously diet-treated patients with type 2 diabetes Hollander, P. A.; Schwartz, S. L.; Gatlin, M. R.; Haas, S. J.; Zheng, H.; Foley, J. E.; Dunning, B. E.  Diabetes Care Jun 2001;24(6):983-8 | Industry Trial in a journal with IF< 30 |
| Riddle 2007 Pramlintide improved glycemic control and reduced weight in patients with type 2 diabetes using basal insulin Riddle, M.; Frias, J.; Zhang, B.; Maier, H.; Brown, C.; Lutz, K.; Kolterman, O.  Diabetes Care Nov 2007;30(11):2794-9 | Industry Trial in a journal with IF< 30 |
| Pasquel 2020 A Randomized Controlled Trial Comparing Glargine U300 and Glargine U100 for the Inpatient Management of Medicine and Surgery Patients With Type 2 Diabetes: Glargine U300 Hospital Trial Pasquel, F. J.; Lansang, M. C.; Khowaja, A.; Urrutia, M. A.; Cardona, S.; Albury, B.; Galindo, R. J.; Fayfman, M.; Davis, G.; Migdal, A.; Vellanki, P.; Peng, L.; Umpierrez, G. E.  Diabetes Care Jun 2020;43(6):1242-1248 | Industry Trial in a journal with IF< 30 |
| Beishuizen 2004 Two-year statin therapy does not alter the progression of intima-media thickness in patients with type 2 diabetes without manifest cardiovascular disease Beishuizen, E. D.; van de Ree, M. A.; Jukema, J. W.; Tamsma, J. T.; van der Vijver, J. C.; Meinders, A. E.; Putter, H.; Huisman, M. V.  Diabetes Care Dec 2004;27(12):2887-92 | Industry Trial in a journal with IF< 30 |
| DeFronzo 2005 Effects of exenatide (exendin-4) on glycemic control and weight over 30 weeks in metformin-treated patients with type 2 diabetes DeFronzo, R. A.; Ratner, R. E.; Han, J.; Kim, D. D.; Fineman, M. S.; Baron, A. D.  Diabetes Care May 2005;28(5):1092-100 | Industry Trial in a journal with IF< 30 |
| DeFronzo 2005 Efficacy of inhaled insulin in patients with type 2 diabetes not controlled with diet and exercise: a 12-week, randomized, comparative trial DeFronzo, R. A.; Bergenstal, R. M.; Cefalu, W. T.; Pullman, J.; Lerman, S.; Bode, B. W.; Phillips, L. S.  Diabetes Care Aug 2005;28(8):1922-8 | Industry Trial in a journal with IF< 30 |
| Bastyr 2000 Therapy focused on lowering postprandial glucose, not fasting glucose, may be superior for lowering HbA1c. IOEZ Study Group Bastyr, E. J., 3rd; Stuart, C. A.; Brodows, R. G.; Schwartz, S.; Graf, C. J.; Zagar, A.; Robertson, K. E.  Diabetes Care Sep 2000;23(9):1236-41 | Industry Trial in a journal with IF< 30 |
| Kaku 2013 Randomized, double-blind, dose-ranging study of TAK-875, a novel GPR40 agonist, in Japanese patients with inadequately controlled type 2 diabetes Kaku, K.; Araki, T.; Yoshinaka, R.  Diabetes Care Feb 2013;36(2):245-50 | Industry Trial in a journal with IF< 30 |
| Not all patients had T2DM | 13 |
| Gerstein 2012 Basal insulin and cardiovascular and other outcomes in dysglycemia Gerstein, H. C.; Bosch, J.; Dagenais, G. R.; Díaz, R.; Jung, H.; Maggioni, A. P.; Pogue, J.; Probstfield, J.; Ramachandran, A.; Riddle, M. C.; Rydén, L. E.; Yusuf, S.  N Engl J Med Jul 26 2012;367(4):319-28 | Not all patients had T2DM |
| Gerstein 2006 Effect of rosiglitazone on the frequency of diabetes in patients with impaired glucose tolerance or impaired fasting glucose: a randomised controlled trial Gerstein, H. C.; Yusuf, S.; Bosch, J.; Pogue, J.; Sheridan, P.; Dinccag, N.; Hanefeld, M.; Hoogwerf, B.; Laakso, M.; Mohan, V.; Shaw, J.; Zinman, B.; Holman, R. R.  Lancet Sep 23 2006;368(9541):1096-105 | Not all patients had T2DM |
| Heerspink 2020 Dapagliflozin in Patients with Chronic Kidney Disease Heerspink, H. J. L.; Stefánsson, B. V.; Correa-Rotter, R.; Chertow, G. M.; Greene, T.; Hou, F. F.; Mann, J. F. E.; McMurray, J. J. V.; Lindberg, M.; Rossing, P.; Sjöström, C. D.; Toto, R. D.; Langkilde, A. M.; Wheeler, D. C.  N Engl J Med Oct 8 2020;383(15):1436-1446 | Not all patients had T2DM |
| Bosch 2012 n-3 fatty acids and cardiovascular outcomes in patients with dysglycemia Bosch, J.; Gerstein, H. C.; Dagenais, G. R.; Díaz, R.; Dyal, L.; Jung, H.; Maggiono, A. P.; Probstfield, J.; Ramachandran, A.; Riddle, M. C.; Rydén, L. E.; Yusuf, S.  N Engl J Med Jul 26 2012;367(4):309-18 | Not all patients had T2DM |
| Lee 2021 Effect of Empagliflozin on Left Ventricular Volumes in Patients With Type 2 Diabetes, or Prediabetes, and Heart Failure With Reduced Ejection Fraction (SUGAR-DM-HF) Lee, M. M. Y.; Brooksbank, K. J. M.; Wetherall, K.; Mangion, K.; Roditi, G.; Campbell, R. T.; Berry, C.; Chong, V.; Coyle, L.; Docherty, K. F.; Dreisbach, J. G.; Labinjoh, C.; Lang, N. N.; Lennie, V.; McConnachie, A.; Murphy, C. L.; Petrie, C. J.; Petrie, J. R.; Speirits, I. A.; Sourbron, S.; Welsh, P.; Woodward, R.; Radjenovic, A.; Mark, P. B.; McMurray, J. J. V.; Jhund, P. S.; Petrie, M. C.; Sattar, N.  Circulation Feb 9 2021;143(6):516-525 | Not all patients had T2DM |
| Lindholm 2002 Cardiovascular morbidity and mortality in patients with diabetes in the Losartan Intervention For Endpoint reduction in hypertension study (LIFE): a randomised trial against atenolol Lindholm, L. H.; Ibsen, H.; Dahlöf, B.; Devereux, R. B.; Beevers, G.; de Faire, U.; Fyhrquist, F.; Julius, S.; Kjeldsen, S. E.; Kristiansson, K.; Lederballe-Pedersen, O.; Nieminen, M. S.; Omvik, P.; Oparil, S.; Wedel, H.; Aurup, P.; Edelman, J.; Snapinn, S.  Lancet Mar 23 2002;359(9311):1004-10 | Not all patients had T2DM |
| #295 The Diabetes Prevention Program: baseline characteristics of the randomized cohort. The Diabetes Prevention Program Research Group Diabetes Care Nov 2000;23(11):1619-29 | Not all patients had T2DM |
| DeFronzo 2011 Pioglitazone for diabetes prevention in impaired glucose tolerance DeFronzo, R. A.; Tripathy, D.; Schwenke, D. C.; Banerji, M.; Bray, G. A.; Buchanan, T. A.; Clement, S. C.; Henry, R. R.; Hodis, H. N.; Kitabchi, A. E.; Mack, W. J.; Mudaliar, S.; Ratner, R. E.; Williams, K.; Stentz, F. B.; Musi, N.; Reaven, P. D.  N Engl J Med Mar 24 2011;364(12):1104-15 | Not all patients had T2DM |
| #1 Effects of ramipril on cardiovascular and microvascular outcomes in people with diabetes mellitus: results of the HOPE study and MICRO-HOPE substudy. Heart Outcomes Prevention Evaluation Study Investigators Lancet Jan 22 2000;355(9200):253-9  Upload full text Abstract 2 Notes History Duplicate Move study to Full text review | Not all patients had T2DM |
| Kirkman 2006 Treating postprandial hyperglycemia does not appear to delay progression of early type 2 diabetes: the Early Diabetes Intervention Program Kirkman, M. S.; Shankar, R. R.; Shankar, S.; Shen, C.; Brizendine, E.; Baron, A.; McGill, J.  Diabetes Care Sep 2006;29(9):2095-101 | Not all patients have T2DM |
| Gross 2015 Panretinal Photocoagulation vs Intravitreous Ranibizumab for Proliferative Diabetic Retinopathy: A Randomized Clinical Trial Gross, J. G.; Glassman, A. R.; Jampol, L. M.; Inusah, S.; Aiello, L. P.; Antoszyk, A. N.; Baker, C. W.; Berger, B. B.; Bressler, N. M.; Browning, D.; Elman, M. J.; Ferris, F. L., 3rd; Friedman, S. M.; Marcus, D. M.; Melia, M.; Stockdale, C. R.; Sun, J. K.; Beck, R. W.  Jama Nov 24 2015;314(20):2137-2146 | Not all patients have T2DM |
| Petrak 2015 Cognitive Behavioral Therapy Versus Sertraline in Patients With Depression and Poorly Controlled Diabetes: The Diabetes and Depression (DAD) Study: A Randomized Controlled Multicenter Trial Petrak, F.; Herpertz, S.; Albus, C.; Hermanns, N.; Hiemke, C.; Hiller, W.; Kronfeld, K.; Kruse, J.; Kulzer, B.; Ruckes, C.; Zahn, D.; Müller, M. J.  Diabetes Care May 2015;38(5):767-75 | Not all patients have T2DM |
| Echeverry 2009 Effect of pharmacological treatment of depression on A1C and quality of life in low-income Hispanics and African Americans with diabetes: a randomized, double-blind, placebo-controlled trial Echeverry, D.; Duran, P.; Bonds, C.; Lee, M.; Davidson, M. B.  Diabetes Care Dec 2009;32(12):2156-60 | Not all patients have T2DM |
| Not government nor industry-funded | 7 |
| Uzu 2007 Reduction of microalbuminuria in patients with type 2 diabetes: the Shiga Microalbuminuria Reduction Trial (SMART) Uzu, T.; Sawaguchi, M.; Maegawa, H.; Kashiwagi, A.  Diabetes Care Jun 2007;30(6):1581-3 | Not government nor industry-funded |
| Sasso 2002 Irbesartan reduces the albumin excretion rate in microalbuminuric type 2 diabetic patients independently of hypertension: a randomized double-blind placebo-controlled crossover study Sasso, F. C.; Carbonara, O.; Persico, M.; Iafusco, D.; Salvatore, T.; D'Ambrosio, R.; Torella, R.; Cozzolino, D.  Diabetes Care Nov 2002;25(11):1909-13 | Not government nor industry-funded |
| Athyros 2002 Atorvastatin and micronized fenofibrate alone and in combination in type 2 diabetes with combined hyperlipidemia Athyros, V. G.; Papageorgiou, A. A.; Athyrou, V. V.; Demitriadis, D. S.; Kontopoulos, A. G.  Diabetes Care Jul 2002;25(7):1198-202 | Not government nor industry-funded |
| Pruski 2009 Pleiotropic action of short-term metformin and fenofibrate treatment, combined with lifestyle intervention, in type 2 diabetic patients with mixed dyslipidemia Pruski, M.; Krysiak, R.; Okopien, B.  Diabetes Care Aug 2009;32(8):1421-4 | Not government nor industry-funded |
| Khan 2002 A prospective, randomized comparison of the metabolic effects of pioglitazone or rosiglitazone in patients with type 2 diabetes who were previously treated with troglitazone Khan, M. A.; St Peter, J. V.; Xue, J. L.  Diabetes Care Apr 2002;25(4):708-11 | Not government nor industry-funded |
| Meyer 2010 Glulisine versus human regular insulin in combination with glargine in noncritically ill hospitalized patients with type 2 diabetes: a randomized double-blind study Meyer, C.; Boron, A.; Plummer, E.; Voltchenok, M.; Vedda, R.  Diabetes Care Dec 2010;33(12):2496-501 | Not government nor industry-funded |
| Esposito 2008 Addition of neutral protamine lispro insulin or insulin glargine to oral type 2 diabetes regimens for patients with suboptimal glycemic control: a randomized trial Esposito, K.; Ciotola, M.; Maiorino, M. I.; Gualdiero, R.; Schisano, B.; Ceriello, A.; Beneduce, F.; Feola, G.; Giugliano, D.  Ann Intern Med Oct 21 2008;149(8):531-9 | Not government nor industry-funded |
| Not RCT of a T2DM pharmacotherapy | 54 |
| Middelkoop 2001 Effectiveness of culture-specific diabetes care for Surinam South Asian patients in the Hague: a randomized controlled trial/controlled before-and-after study Middelkoop, B. J.; Geelhoed-Duijvestijn, P. H.; van der Wal, G.  Diabetes Care Nov 2001;24(11):1997-8 | Not RCT of a T2DM pharmacotherapy |
| García-Patterson 2001 Evaluation of light exercise in the treatment of gestational diabetes García-Patterson, A.; Martín, E.; Ubeda, J.; María, M. A.; de Leiva, A.; Corcoy, R.  Diabetes Care Nov 2001;24(11):2006-7 | Not RCT of a T2DM pharmacotherapy |
| Schauer 2014 Bariatric surgery versus intensive medical therapy for diabetes--3-year outcomes Schauer, P. R.; Bhatt, D. L.; Kirwan, J. P.; Wolski, K.; Brethauer, S. A.; Navaneethan, S. D.; Aminian, A.; Pothier, C. E.; Kim, E. S.; Nissen, S. E.; Kashyap, S. R.  N Engl J Med May 22 2014;370(21):2002-13 | Not RCT of a T2DM pharmacotherapy |
| Fukui 2003 Glycyrrhizin and serum testosterone concentrations in male patients with type 2 diabetes Fukui, M.; Kitagawa, Y.; Nakamura, N.; Yoshikawa, T.  Diabetes Care Oct 2003;26(10):2962 | Not RCT of a T2DM pharmacotherapy |
| Marre 2000 Determinants of elevated urinary albumin in the 4,937 type 2 diabetic subjects recruited for the DIABHYCAR Study in Western Europe and North Africa Marre, M.; Lièvre, M.; Vasmant, D.; Gallois, Y.; Hadjadj, S.; Reglier, J. C.; Chatellier, G.; Mann, J.; Viberti, G. C.; Passa, P.  Diabetes Care Apr 2000;23 Suppl 2():B40-8  Upload full text Abstract Note History Duplicate Move study to Full text review | Not RCT of a T2DM pharmacotherapy |
| 5-Year Outcomes Schauer, P. R.; Bhatt, D. L.; Kirwan, J. P.; Wolski, K.; Aminian, A.; Brethauer, S. A.; Navaneethan, S. D.; Singh, R. P.; Pothier, C. E.; Nissen, S. E.; Kashyap, S. R.  N Engl J Med Feb 16 2017;376(7):641-651 | Not RCT of a T2DM pharmacotherapy |
| Sabatine 2017 Evolocumab and Clinical Outcomes in Patients with Cardiovascular Disease Sabatine, M. S.; Giugliano, R. P.; Keech, A. C.; Honarpour, N.; Wiviott, S. D.; Murphy, S. A.; Kuder, J. F.; Wang, H.; Liu, T.; Wasserman, S. M.; Sever, P. S.; Pedersen, T. R.  N Engl J Med May 4 2017;376(18):1713-1722 | Not RCT of a T2DM pharmacotherapy |
| McMurray 2019 Dapagliflozin in Patients with Heart Failure and Reduced Ejection Fraction McMurray, J. J. V.; Solomon, S. D.; Inzucchi, S. E.; Køber, L.; Kosiborod, M. N.; Martinez, F. A.; Ponikowski, P.; Sabatine, M. S.; Anand, I. S.; Bělohlávek, J.; Böhm, M.; Chiang, C. E.; Chopra, V. K.; de Boer, R. A.; Desai, A. S.; Diez, M.; Drozdz, J.; Dukát, A.; Ge, J.; Howlett, J. G.; Katova, T.; Kitakaze, M.; Ljungman, C. E. A.; Merkely, B.; Nicolau, J. C.; O'Meara, E.; Petrie, M. C.; Vinh, P. N.; Schou, M.; Tereshchenko, S.; Verma, S.; Held, C.; DeMets, D. L.; Docherty, K. F.; Jhund, P. S.; Bengtsson, O.; Sjöstrand, M.; Langkilde, A. M.  N Engl J Med Nov 21 2019;381(21):1995-2008 | Not RCT of a T2DM pharmacotherapy |
| Holman 2008 Long-term follow-up after tight control of blood pressure in type 2 diabetes Holman, R. R.; Paul, S. K.; Bethel, M. A.; Neil, H. A.; Matthews, D. R.  N Engl J Med Oct 9 2008;359(15):1565-76 | Not RCT of a T2DM pharmacotherapy |
| Antoszyk 2020 Effect of Intravitreous Aflibercept vs Vitrectomy With Panretinal Photocoagulation on Visual Acuity in Patients With Vitreous Hemorrhage From Proliferative Diabetic Retinopathy: A Randomized Clinical Trial Antoszyk, A. N.; Glassman, A. R.; Beaulieu, W. T.; Jampol, L. M.; Jhaveri, C. D.; Punjabi, O. S.; Salehi-Had, H.; Wells, J. A., 3rd; Maguire, M. G.; Stockdale, C. R.; Martin, D. F.; Sun, J. K.  Jama Dec 15 2020;324(23):2383-2395 | Not RCT of a T2DM pharmacotherapy |
| Ikramuddin 2013 Roux-en-Y gastric bypass vs intensive medical management for the control of type 2 diabetes, hypertension, and hyperlipidemia: the Diabetes Surgery Study randomized clinical trial Ikramuddin, S.; Korner, J.; Lee, W. J.; Connett, J. E.; Inabnet, W. B.; Billington, C. J.; Thomas, A. J.; Leslie, D. B.; Chong, K.; Jeffery, R. W.; Ahmed, L.; Vella, A.; Chuang, L. M.; Bessler, M.; Sarr, M. G.; Swain, J. M.; Laqua, P.; Jensen, M. D.; Bantle, J. P.  Jama Jun 5 2013;309(21):2240-9 | Not RCT of a T2DM pharmacotherapy |
| Cosman 2016 Romosozumab Treatment in Postmenopausal Women with Osteoporosis Cosman, F.; Crittenden, D. B.; Adachi, J. D.; Binkley, N.; Czerwinski, E.; Ferrari, S.; Hofbauer, L. C.; Lau, E.; Lewiecki, E. M.; Miyauchi, A.; Zerbini, C. A.; Milmont, C. E.; Chen, L.; Maddox, J.; Meisner, P. D.; Libanati, C.; Grauer, A.  N Engl J Med Oct 20 2016;375(16):1532-1543 | Not RCT of a T2DM pharmacotherapy |
| Simpson 2011 Effect of adding pharmacists to primary care teams on blood pressure control in patients with type 2 diabetes: a randomized controlled trial Simpson, S. H.; Majumdar, S. R.; Tsuyuki, R. T.; Lewanczuk, R. Z.; Spooner, R.; Johnson, J. A.  Diabetes Care Jan 2011;34(1):20-6 | Not RCT of a T2DM pharmacotherapy |
| Stub 2015 Air Versus Oxygen in ST-Segment-Elevation Myocardial Infarction Stub, D.; Smith, K.; Bernard, S.; Nehme, Z.; Stephenson, M.; Bray, J. E.; Cameron, P.; Barger, B.; Ellims, A. H.; Taylor, A. J.; Meredith, I. T.; Kaye, D. M.  Circulation Jun 16 2015;131(24):2143-50 | Not RCT of a T2DM pharmacotherapy |
| TenKulve 2016 Liraglutide Reduces CNS Activation in Response to Visual Food Cues Only After Short-term Treatment in Patients With Type 2 Diabetes Ten Kulve, J. S.; Veltman, D. J.; van Bloemendaal, L.; Barkhof, F.; Drent, M. L.; Diamant, M.; I. Jzerman RG  Diabetes Care Feb 2016;39(2):214-21 | Not RCT of a T2DM pharmacotherapy |
| Wu 2004 The effect of diabetes on B-type natriuretic peptide concentrations in patients with acute dyspnea: an analysis from the Breathing Not Properly Multinational Study Wu, A. H.; Omland, T.; Duc, P.; McCord, J.; Nowak, R. M.; Hollander, J. E.; Herrmann, H. C.; Steg, P. G.; Wold Knudsen, C.; Storrow, A. B.; Abraham, W. T.; Perez, A.; Kamin, R.; Clopton, P.; Maisel, A. S.; McCullough, P. A.  Diabetes Care Oct 2004;27(10):2398-404 | Not RCT of a T2DM pharmacotherapy |
| Xie 2020 Comparative Effectiveness of SGLT2 Inhibitors, GLP-1 Receptor Agonists, DPP-4 Inhibitors, and Sulfonylureas on Risk of Kidney Outcomes: Emulation of a Target Trial Using Health Care Databases Xie, Y.; Bowe, B.; Gibson, A. K.; McGill, J. B.; Maddukuri, G.; Yan, Y.; Al-Aly, Z.  Diabetes Care Nov 2020;43(11):2859-2869 | Not RCT of a T2DM pharmacotherapy |
| Mehler 2003 Intensive blood pressure control reduces the risk of cardiovascular events in patients with peripheral arterial disease and type 2 diabetes Mehler, P. S.; Coll, J. R.; Estacio, R.; Esler, A.; Schrier, R. W.; Hiatt, W. R.  Circulation Feb 11 2003;107(5):753-6 | Not RCT of a T2DM pharmacotherapy |
| Yang 2013 Primary prevention of macroangiopathy in patients with short-duration type 2 diabetes by intensified multifactorial intervention: seven-year follow-up of diabetes complications in Chinese Yang, Y.; Yao, J. J.; Du, J. L.; Bai, R.; Sun, L. P.; Sun, G. H.; Song, G. R.; Cao, S. M.; Shi, C. H.; Ba, Y.; Xing, Q.; Zhang, X. Y.  Diabetes Care Apr 2013;36(4):978-84 | Not RCT of T2DM Pharmacotherapy |
| Dungan 2013 Prandial insulin dosing using the carbohydrate counting technique in hospitalized patients with type 2 diabetes Dungan, K. M.; Sagrilla, C.; Abdel-Rasoul, M.; Osei, K.  Diabetes Care Nov 2013;36(11):3476-82 | Not RCT of T2DM Pharmacotherapy |
| Linnebjerg 2015 Comparison of the Pharmacokinetics and Pharmacodynamics of LY2963016 Insulin Glargine and EU- and US-Approved Versions of Lantus Insulin Glargine in Healthy Subjects: Three Randomized Euglycemic Clamp Studies Linnebjerg, H.; Lam, E. C.; Seger, M. E.; Coutant, D.; Chua, L.; Chong, C. L.; Ferreira, M. M.; Soon, D.; Zhang, X.  Diabetes Care Dec 2015;38(12):2226-33 | Not RCT of T2DM Pharmacotherapy |
| Clifford 2005 Effect of a pharmaceutical care program on vascular risk factors in type 2 diabetes: the Fremantle Diabetes Study Clifford, R. M.; Davis, W. A.; Batty, K. T.; Davis, T. M.  Diabetes Care Apr 2005;28(4):771-6 | Not RCT of T2DM Pharmacotherapy |
| Collins 2011 Effects of a home-based walking intervention on mobility and quality of life in people with diabetes and peripheral arterial disease: a randomized controlled trial Collins, T. C.; Lunos, S.; Carlson, T.; Henderson, K.; Lightbourne, M.; Nelson, B.; Hodges, J. S.  Diabetes Care Oct 2011;34(10):2174-9 | Not RCT of T2DM Pharmacotherapy |
| Chen 2009 Metabolic syndrome and salt sensitivity of blood pressure in non-diabetic people in China: a dietary intervention study Chen, J.; Gu, D.; Huang, J.; Rao, D. C.; Jaquish, C. E.; Hixson, J. E.; Chen, C. S.; Chen, J.; Lu, F.; Hu, D.; Rice, T.; Kelly, T. N.; Hamm, L. L.; Whelton, P. K.; He, J.  Lancet Mar 7 2009;373(9666):829-35 | Not RCT of T2DM Pharmacotherapy |
| Majumdar 2003 Controlled trial of a multifaceted intervention for improving quality of care for rural patients with type 2 diabetes Majumdar, S. R.; Guirguis, L. M.; Toth, E. L.; Lewanczuk, R. Z.; Lee, T. K.; Johnson, J. A.  Diabetes Care Nov 2003;26(11):3061-6 | Not RCT of T2DM Pharmacotherapy |
| Schillinger 2009 Effects of self-management support on structure, process, and outcomes among vulnerable patients with diabetes: a three-arm practical clinical trial Schillinger, D.; Handley, M.; Wang, F.; Hammer, H.  Diabetes Care Apr 2009;32(4):559-66 | Not RCT of T2DM Pharmacotherapy |
| Keyserling 2002 A randomized trial of an intervention to improve self-care behaviors of African-American women with type 2 diabetes: impact on physical activity Keyserling, T. C.; Samuel-Hodge, C. D.; Ammerman, A. S.; Ainsworth, B. E.; Henríquez-Roldán, C. F.; Elasy, T. A.; Skelly, A. H.; Johnston, L. F.; Bangdiwala, S. I.  Diabetes Care Sep 2002;25(9):1576-83 | Not RCT of T2DM Pharmacotherapy |
| DePue 2013 Nurse-community health worker team improves diabetes care in American Samoa: results of a randomized controlled trial DePue, J. D.; Dunsiger, S.; Seiden, A. D.; Blume, J.; Rosen, R. K.; Goldstein, M. G.; Nu'usolia, O.; Tuitele, J.; McGarvey, S. T.  Diabetes Care Jul 2013;36(7):1947-53 | Not RCT of T2DM Pharmacotherapy |
| Hermida 2011 Influence of time of day of blood pressure-lowering treatment on cardiovascular risk in hypertensive patients with type 2 diabetes Hermida, R. C.; Ayala, D. E.; Mojón, A.; Fernández, J. R.  Diabetes Care Jun 2011;34(6):1270-6 | Not RCT of T2DM Pharmacotherapy |
| Heisler 2010 Diabetes control with reciprocal peer support versus nurse care management: a randomized trial Heisler, M.; Vijan, S.; Makki, F.; Piette, J. D.  Ann Intern Med Oct 19 2010;153(8):507-15 | Not RCT of T2DM Pharmacotherapy |
| Polonsky 2011 Structured self-monitoring of blood glucose significantly reduces A1C levels in poorly controlled, noninsulin-treated type 2 diabetes: results from the Structured Testing Program study Polonsky, W. H.; Fisher, L.; Schikman, C. H.; Hinnen, D. A.; Parkin, C. G.; Jelsovsky, Z.; Petersen, B.; Schweitzer, M.; Wagner, R. S.  Diabetes Care Feb 2011;34(2):262-7 | Not RCT of T2DM Pharmacotherapy |
| Salas-Salvadó 2011 Reduction in the incidence of type 2 diabetes with the Mediterranean diet: results of the PREDIMED-Reus nutrition intervention randomized trial Salas-Salvadó, J.; Bulló, M.; Babio, N.; Martínez-González, MÁ; Ibarrola-Jurado, N.; Basora, J.; Estruch, R.; Covas, M. I.; Corella, D.; Arós, F.; Ruiz-Gutiérrez, V.; Ros, E.  Diabetes Care Jan 2011;34(1):14-9 | Not RCT of T2DM Pharmacotherapy |
| Ulbrecht 2014 Prevention of recurrent foot ulcers with plantar pressure-based in-shoe orthoses: the CareFUL prevention multicenter randomized controlled trial Ulbrecht, J. S.; Hurley, T.; Mauger, D. T.; Cavanagh, P. R.  Diabetes Care Jul 2014;37(7):1982-9 | Not RCT of T2DM Pharmacotherapy |
| Ali 2020 Effect of a Collaborative Care Model on Depressive Symptoms and Glycated Hemoglobin, Blood Pressure, and Serum Cholesterol Among Patients With Depression and Diabetes in India: The INDEPENDENT Randomized Clinical Trial Ali, M. K.; Chwastiak, L.; Poongothai, S.; Emmert-Fees, K. M. F.; Patel, S. A.; Anjana, R. M.; Sagar, R.; Shankar, R.; Sridhar, G. R.; Kosuri, M.; Sosale, A. R.; Sosale, B.; Rao, D.; Tandon, N.; Narayan, K. M. V.; Mohan, V.  Jama Aug 18 2020;324(7):651-662 | Not RCT of T2DM Pharmacotherapy |
| Legrand 2004 Three-year outcome after coronary stenting versus bypass surgery for the treatment of multivessel disease Legrand, V. M.; Serruys, P. W.; Unger, F.; van Hout, B. A.; Vrolix, M. C.; Fransen, G. M.; Nielsen, T. T.; Paulsen, P. K.; Gomes, R. S.; de Queiroz e Melo, J. M.; Neves, J. P.; Lindeboom, W.; Backx, B.  Circulation Mar 9 2004;109(9):1114-20 | Not RCT of T2DM Pharmacotherapy |
| Orr 2006 Mobility impairment in type 2 diabetes: association with muscle power and effect of Tai Chi intervention Orr, R.; Tsang, T.; Lam, P.; Comino, E.; Singh, M. F.  Diabetes Care Sep 2006;29(9):2120-2 | Not RCT of T2DM Pharmacotherapy |
| Thoolen 2006 Psychological outcomes of patients with screen-detected type 2 diabetes: the influence of time since diagnosis and treatment intensity Thoolen, B. J.; de Ridder, D. T.; Bensing, J. M.; Gorter, K. J.; Rutten, G. E.  Diabetes Care Oct 2006;29(10):2257-62 | Not RCT of T2DM Pharmacotherapy |
| Keegan 2004 Effect of alendronate on bone mineral density and biochemical markers of bone turnover in type 2 diabetic women: the fracture intervention trial Keegan, T. H.; Schwartz, A. V.; Bauer, D. C.; Sellmeyer, D. E.; Kelsey, J. L.  Diabetes Care Jul 2004;27(7):1547-53 | Not RCT of T2DM Pharmacotherapy |
| Freund 2016 Medical Assistant-Based Care Management for High-Risk Patients in Small Primary Care Practices: A Cluster Randomized Clinical Trial Freund, T.; Peters-Klimm, F.; Boyd, C. M.; Mahler, C.; Gensichen, J.; Erler, A.; Beyer, M.; Gondan, M.; Rochon, J.; Gerlach, F. M.; Szecsenyi, J.  Ann Intern Med Mar 1 2016;164(5):323-30 | Not RCT of T2DM Pharmacotherapy |
| Li 2004 Induction of long-term glycemic control in newly diagnosed type 2 diabetic patients is associated with improvement of beta-cell function Li, Y.; Xu, W.; Liao, Z.; Yao, B.; Chen, X.; Huang, Z.; Hu, G.; Weng, J.  Diabetes Care Nov 2004;27(11):2597-602 | Not RCT of T2DM Pharmacotherapy |
| Pandey 2020 Association of Intensive Lifestyle Intervention, Fitness, and Body Mass Index With Risk of Heart Failure in Overweight or Obese Adults With Type 2 Diabetes Mellitus: An Analysis From the Look AHEAD Trial Pandey, A.; Patel, K. V.; Bahnson, J. L.; Gaussoin, S. A.; Martin, C. K.; Balasubramanyam, A.; Johnson, K. C.; McGuire, D. K.; Bertoni, A. G.; Kitzman, D.; Berry, J. D.  Circulation Apr 21 2020;141(16):1295-1306 | Not RCT of T2DM Pharmacotherapy |
| #2 Closing the gap: effect of diabetes case management on glycemic control among low-income ethnic minority populations: the California Medi-Cal type 2 diabetes study Diabetes Care Jan 2004;27(1):95-103 | Not RCT of T2DM Pharmacotherapy |
| Canga 2000 Intervention study for smoking cessation in diabetic patients: a randomized controlled trial in both clinical and primary care settings Canga, N.; De Irala, J.; Vara, E.; Duaso, M. J.; Ferrer, A.; Martínez-González, M. A.  Diabetes Care Oct 2000;23(10):1455-60 | Not RCT of T2DM Pharmacotherapy |
| Howard 2008 Effect of lower targets for blood pressure and LDL cholesterol on atherosclerosis in diabetes: the SANDS randomized trial Howard, B. V.; Roman, M. J.; Devereux, R. B.; Fleg, J. L.; Galloway, J. M.; Henderson, J. A.; Howard, W. J.; Lee, E. T.; Mete, M.; Poolaw, B.; Ratner, R. E.; Russell, M.; Silverman, A.; Stylianou, M.; Umans, J. G.; Wang, W.; Weir, M. R.; Weissman, N. J.; Wilson, C.; Yeh, F.; Zhu, J.  Jama Apr 9 2008;299(14):1678-89 | Not RCT of T2DM Pharmacotherapy |
| House 2010 Effect of B-vitamin therapy on progression of diabetic nephropathy: a randomized controlled trial House, A. A.; Eliasziw, M.; Cattran, D. C.; Churchill, D. N.; Oliver, M. J.; Fine, A.; Dresser, G. K.; Spence, J. D.  Jama Apr 28 2010;303(16):1603-9 | Not RCT of T2DM Pharmacotherapy |
| Müller 2013 Randomized crossover study to examine the necessity of an injection-to-meal interval in patients with type 2 diabetes and human insulin Müller, N.; Frank, T.; Kloos, C.; Lehmann, T.; Wolf, G.; Müller, U. A.  Diabetes Care Jul 2013;36(7):1865-9 | Not RCT of T2DM Pharmacotherapy |
| Bergenstal 2019 Automated insulin dosing guidance to optimise insulin management in patients with type 2 diabetes: a multicentre, randomised controlled trial Bergenstal, R. M.; Johnson, M.; Passi, R.; Bhargava, A.; Young, N.; Kruger, D. F.; Bashan, E.; Bisgaier, S. G.; Isaman, D. J. M.; Hodish, I.  Lancet Mar 16 2019;393(10176):1138-1148 | Not RCT of T2DM Pharmacotherapy |
| deBoer 2019 Effect of Vitamin D and Omega-3 Fatty Acid Supplementation on Kidney Function in Patients With Type 2 Diabetes: A Randomized Clinical Trial de Boer, I. H.; Zelnick, L. R.; Ruzinski, J.; Friedenberg, G.; Duszlak, J.; Bubes, V. Y.; Hoofnagle, A. N.; Thadhani, R.; Glynn, R. J.; Buring, J. E.; Sesso, H. D.; Manson, J. E.  Jama Nov 19 2019;322(19):1899-1909 | Not RCT of T2DM Pharmacotherapy |
| Verdecchia 2009 Usual versus tight control of systolic blood pressure in non-diabetic patients with hypertension (Cardio-Sis): an open-label randomised trial Verdecchia, P.; Staessen, J. A.; Angeli, F.; de Simone, G.; Achilli, A.; Ganau, A.; Mureddu, G.; Pede, S.; Maggioni, A. P.; Lucci, D.; Reboldi, G.  Lancet Aug 15 2009;374(9689):525-33 | Not RCT of T2DM Pharmacotherapy |
| Nathan 2013 Rationale and design of the glycemia reduction approaches in diabetes: a comparative effectiveness study (GRADE) Nathan, D. M.; Buse, J. B.; Kahn, S. E.; Krause-Steinrauf, H.; Larkin, M. E.; Staten, M.; Wexler, D.; Lachin, J. M.  Diabetes Care Aug 2013;36(8):2254-61 | Not RCT of T2DM Pharmacotherapy |
| deZeeuw 2010 Selective vitamin D receptor activation with paricalcitol for reduction of albuminuria in patients with type 2 diabetes (VITAL study): a randomised controlled trial de Zeeuw, D.; Agarwal, R.; Amdahl, M.; Audhya, P.; Coyne, D.; Garimella, T.; Parving, H. H.; Pritchett, Y.; Remuzzi, G.; Ritz, E.; Andress, D.  Lancet Nov 6 2010;376(9752):1543-51 | Not RCT of T2DM Pharmacotherapy |
| Denver 2003 Management of uncontrolled hypertension in a nurse-led clinic compared with conventional care for patients with type 2 diabetes Denver, E. A.; Barnard, M.; Woolfson, R. G.; Earle, K. A.  Diabetes Care Aug 2003;26(8):2256-60 | Not RCT of T2DM Pharmacotherapy |
| Taniguchi 2000 Effect of physical training on insulin sensitivity in Japanese type 2 diabetic patients: role of serum triglyceride levels Taniguchi, A.; Fukushima, M.; Sakai, M.; Nagasaka, S.; Doi, K.; Nagata, I.; Matsushita, K.; Ooyama, Y.; Kawamoto, A.; Nakasone, M.; Tokuyama, K.; Nakai, Y.  Diabetes Care Jun 2000;23(6):857-8 | Not RCT of T2DM Pharmacotherapy |
| Gaede 2003 Multifactorial intervention and cardiovascular disease in patients with type 2 diabetes Gaede, P.; Vedel, P.; Larsen, N.; Jensen, G. V.; Parving, H. H.; Pedersen, O.  N Engl J Med Jan 30 2003;348(5):383-93 | Not RCT of T2DM Pharmacotherapy |
| Substudy | 1 |
| Mari 2005 Beta-cell function in mild type 2 diabetic patients: effects of 6-month glucose lowering with nateglinide Mari, A.; Gastaldelli, A.; Foley, J. E.; Pratley, R. E.; Ferrannini, E.  Diabetes Care May 2005;28(5):1132-8 | Substudy |
| One Country for Industry Trial | 11 |
| Mazzone 2006 Effect of pioglitazone compared with glimepiride on carotid intima-media thickness in type 2 diabetes: a randomized trial Mazzone, T.; Meyer, P. M.; Feinstein, S. B.; Davidson, M. H.; Kondos, G. T.; D'Agostino, R. B., Sr.; Perez, A.; Provost, J. C.; Haffner, S. M.  Jama Dec 6 2006;296(21):2572-81 | One Country for Industry Trial |
| Fonseca 2000 Effect of metformin and rosiglitazone combination therapy in patients with type 2 diabetes mellitus: a randomized controlled trial Fonseca, V.; Rosenstock, J.; Patwardhan, R.; Salzman, A.  Jama Apr 5 2000;283(13):1695-702 | One Country for Industry Trial |
| Wysham 2017 Effect of Insulin Degludec vs Insulin Glargine U100 on Hypoglycemia in Patients With Type 2 Diabetes: The SWITCH 2 Randomized Clinical Trial Wysham, C.; Bhargava, A.; Chaykin, L.; de la Rosa, R.; Handelsman, Y.; Troelsen, L. N.; Kvist, K.; Norwood, P.  Jama Jul 4 2017;318(1):45-56 | One Country for Industry Trial |
| Bakris 2004 Metabolic effects of carvedilol vs metoprolol in patients with type 2 diabetes mellitus and hypertension: a randomized controlled trial Bakris, G. L.; Fonseca, V.; Katholi, R. E.; McGill, J. B.; Messerli, F. H.; Phillips, R. A.; Raskin, P.; Wright, J. T., Jr.; Oakes, R.; Lukas, M. A.; Anderson, K. M.; Bell, D. S.  Jama Nov 10 2004;292(18):2227-36 | One Country for Industry Trial |
| Lind 2015 Liraglutide in people treated for type 2 diabetes with multiple daily insulin injections: randomised clinical trial (MDI Liraglutide trial) Lind, M.; Hirsch, I. B.; Tuomilehto, J.; Dahlqvist, S.; Ahrén, B.; Torffvit, O.; Attvall, S.; Ekelund, M.; Filipsson, K.; Tengmark, B. O.; Sjöberg, S.; Pehrsson, N. G.  Bmj Oct 28 2015;351():h5364 | One Country for Industry Trial |
| Pradhan 2009 Effects of initiating insulin and metformin on glycemic control and inflammatory biomarkers among patients with type 2 diabetes: the LANCET randomized trial Pradhan, A. D.; Everett, B. M.; Cook, N. R.; Rifai, N.; Ridker, P. M.  Jama Sep 16 2009;302(11):1186-94 | One Country for Industry Trial |
| Zinman 2010 Low-dose combination therapy with rosiglitazone and metformin to prevent type 2 diabetes mellitus (CANOE trial): a double-blind randomised controlled study Zinman, B.; Harris, S. B.; Neuman, J.; Gerstein, H. C.; Retnakaran, R. R.; Raboud, J.; Qi, Y.; Hanley, A. J.  Lancet Jul 10 2010;376(9735):103-11 | One Country for Industry Trial |
| Wanner 2005 Atorvastatin in patients with type 2 diabetes mellitus undergoing hemodialysis Wanner, C.; Krane, V.; März, W.; Olschewski, M.; Mann, J. F.; Ruf, G.; Ritz, E.  N Engl J Med Jul 21 2005;353(3):238-48 | One Country for Industry Trial |
| Pergola 2011 Bardoxolone methyl and kidney function in CKD with type 2 diabetes Pergola, P. E.; Raskin, P.; Toto, R. D.; Meyer, C. J.; Huff, J. W.; Grossman, E. B.; Krauth, M.; Ruiz, S.; Audhya, P.; Christ-Schmidt, H.; Wittes, J.; Warnock, D. G.  N Engl J Med Jul 28 2011;365(4):327-36 | One Country for Industry Trial |
| Ruggenenti 2004 Preventing microalbuminuria in type 2 diabetes Ruggenenti, P.; Fassi, A.; Ilieva, A. P.; Bruno, S.; Iliev, I. P.; Brusegan, V.; Rubis, N.; Gherardi, G.; Arnoldi, F.; Ganeva, M.; Ene-Iordache, B.; Gaspari, F.; Perna, A.; Bossi, A.; Trevisan, R.; Dodesini, A. R.; Remuzzi, G.  N Engl J Med Nov 4 2004;351(19):1941-51 | One Country for Industry Trial |
| Ambery 2018 MEDI0382, a GLP-1 and glucagon receptor dual agonist, in obese or overweight patients with type 2 diabetes: a randomised, controlled, double-blind, ascending dose and phase 2a study Ambery, P.; Parker, V. E.; Stumvoll, M.; Posch, M. G.; Heise, T.; Plum-Moerschel, L.; Tsai, L. F.; Robertson, D.; Jain, M.; Petrone, M.; Rondinone, C.; Hirshberg, B.; Jermutus, L.  Lancet Jun 30 2018;391(10140):2607-2618 | One Country for Industry Trial |
| No participant breakdown by ethnicity/race | 20 |
| Skov 2014 Metformin, but Not Rosiglitazone, Attenuates the Increasing Plasma Levels of a New Cardiovascular Marker, Fibulin-1, in Patients With Type 2 Diabetes Skov, V.; Cangemi, C.; Gram, J.; Christensen, M.M.; Grodum, E.; Sorenson, D.; Argraves, W.S.; Henriksen, J.E.; Rasmussen, L.M. Diabetes Care Feb 11 2014;37(3):760-766 | No participant breakdown by ethnicity/race |
| Pitale 2000 Two years of intensive glycemic control and left ventricular function in the Veterans Affairs Cooperative Study in Type 2 Diabetes Mellitus (VA CSDM) Pitale, S.U.; Abraira, C.; Emanuele, N.V.; McCarren, M.; Henderson, W.G.; Pacold, I.; Bushnell, D.; Colwell, J.A,; Nuttall, F.Q.; Levin, S.R.; Sawin, C.T.; Comstock, J.P.; Silbert, C.K.; Diabetes Care Sep 23 2000;(9):1316-20. | No participant breakdown by ethnicity/race |
| Hong 2013 Effects of Metformin Versus Glipizide on Cardiovascular Outcomes in Patients With Type 2 Diabetes and Coronary Artery Disease Hong, J.; Zhang, Y.; Lai, S.; Lv, A.; Su, Q.; Dong, Y.; Zhou, Z.; Tang, W.; Zhao, J.; Cui, L.; Zou, D.; Wang, D.; Li, H.; Liu, C.; Qu, G.; Shen, J.; Zhu, D.; Wang, W.; Shen, W.; Ning, G. Diabetes Care Apr 13 2013;36(5):1304-1311. | No participant breakdown by ethnicity/race |
| Gram 2011 Pharmacological Treatment of the Pathogenetic Defects in Type 2 Diabetes: The randomized multicenter South Danish Diabetes Study Gram, J.; Henriksen, J.E.; Grodum, E.; Juhl, H.; Hansen, T.B.; Christiansen, C.; Yderstraede, K., Gjessing, H.; Hansen, H.M.; Vestergaard, V.; Hangaard, J.; Beck-Nielsen, H. Diabetes Care Oct 7 2010;34(1):27-33 | No participant breakdown by ethnicity/race |
| Abdul-Ghani 2017 Combination Therapy With Exenatide Plus Pioglitazone Versus Basal/Bolus Insulin in Patients With Poorly Controlled Type 2 Diabetes on Sulfonylurea Plus Metformin: The Qatar Study Abdul-Ghani, M.; Migahid, O.; Megahed, A.; Adams, J.; Triplitt, C.; DeFronzo, R.A.; Zirie, M.; Jayyousi, A. Diabetes Care Mar 2017;40(3):325-331 | No participant breakdown by ethnicity/race |
| Weng 2008 Effect of intensive insulin therapy on β-cell function and glycaemic control in patients with newly diagnosed type 2 diabetes: a multicentre randomised parallel-group trial Weng, J.; Li, Y.; Xu, W.; Shi, L.; Zhang, Q.; Zhu, D.; Hu, Y.; Zhou, Z.; Yan, X.; Tian, H., Ran, X.; Luo, Z.; Xian, J.; Yan, L.; Li, F.; Zeng, L.; Chen, Y.; Yang, L.; Yan, S.; Liu, J.; Li, M.; Fu, Z.; Cheng, H. Lancet 2008;371(9626):1753-1760 | No participant breakdown by ethnicity/race |
| Rosenstock 2020 Once-Weekly Insulin for Type 2 Diabetes without Previous Insulin Treatment Rosenstock, J.; Bajaj, H.S.; Janez, A.; Silver, R. NEJM Nov 26 2020;383:2107-2116 | No participant breakdown by ethnicity/race |
| Reznik 2014 Insulin pump treatment compared with multiple daily injections for treatment of type 2 diabetes (OpT2mise): a randomised open-label controlled trial Reznik, Y.; Cohen, O.; Aronson, R.; Conget. I.; Runzis, S.; Castaneda, J.; Lee, S.W. Lancet Oct 2014;384(9950):1265-1272 | No participant breakdown by ethnicity/race |
| Patel 2007 Effects of a fixed combination of perindopril and indapamide on macrovascular and microvascular outcomes in patients with type 2 diabetes mellitus (the ADVANCE trial): a randomised controlled trial Patel, A. Lancet Sept 2007;370(9590):829-840 | No participant breakdown by ethnicity/race |
| Ogawa 2008 Low-Dose Aspirin for Primary Prevention of Atherosclerotic Events in Patients With Type 2 Diabetes  A Randomized Controlled Trial Ogawa, H.; Nakayama, M.; Morimoto, T. JAMA Nov 12 2008;300(18):2134-2141 | No participant breakdown by ethnicity/race |
| Mogensen 2000 Randomised controlled trial of dual blockade of renin-angiotensin system in patients with hypertension, microalbuminuria, and non-insulin dependent diabetes: the candesartan and lisinopril microalbuminuria (CALM) study Mogensen, C.E.; Neldam, S.; Tikkanen, I.; Oren, S.; Viskoper, R.; Watts, R.W., Cooper, M.E. BMJ Dec 9 2000;321:1440-1444 | No participant breakdown by ethnicity/race |
| Marre 2004 Effects of low dose ramipril on cardiovascular and renal outcomes in patients with type 2 diabetes and raised excretion of urinary albumin: randomised, double blind, placebo controlled trial (the DIABHYCAR study) Marre, M.; Lievre, M.; Chatellier, G.; Mann, J.F.E.; Passa, P.; Menard, J. BMJ Mar 18 2004;328:495 | No participant breakdown by ethnicity/race |
| Katakami 2010 The Phosphodiesterase Inhibitor Cilostazol Induces Regression of Carotid Atherosclerosis in Subjects With Type 2 Diabetes Mellitus Katakami, N.; Kim, Y.; Kawamori, R.; Yamasaki, Y. Circulation Jun 1 2010;121:2584-2591 | No participant breakdown by ethnicity/race |
| Henry 2009 Effect of the dual peroxisome proliferator-activated receptor-α/γ agonist aleglitazar on risk of cardiovascular disease in patients with type 2 diabetes (SYNCHRONY): a phase II, randomised, dose-ranging study Henry, R.R.; Lincoff, M.; Mudaliar, S.; Rabbia, M.; Chognot, C.; Herz, M. Lancet Jul 2009;374(9684):126-135 | No participant breakdown by ethnicity/race |
| Haritoglou 2009 Effect of calcium dobesilate on occurrence of diabetic macular oedema (CALDIRET study): randomised, double-blind, placebo-controlled, multicentre trial Haritoglou, C.; Gerss, J.; Sauerland, C.; Kampik, A.; Ulbig, M.W. Lancet Apr 2009;373(9672):1364-1371 | No participant breakdown by ethnicity/race |
| Esposito 2004 Regression of Carotid Atherosclerosis by Control of Postprandial Hyperglycemia in Type 2 Diabetes Mellitus Esposito, K.; Giugliano, D.; Nappo, F.; Marfella, R. Circulation Jun 14 2004;110:214-219 | No participant breakdown by ethnicity/race |
| Bretzel 2008 Once-daily basal insulin glargine versus thrice-daily prandial insulin lispro in people with type 2 diabetes on oral hypoglycaemic agents (APOLLO): an open randomised controlled trial Bretzel, R.G.; Nuber, U.; Landgraf, W.; Owens, D.R.; Bradley, C.; Linn, T. Lancet April 2008;371(9618):1047-1048 | No participant breakdown by ethnicity/race |
| Balley 2018 Closed-Loop Insulin for Glycemic Control in Noncritical Care NEJM Nov 15 2018;379:1970-1971 | No participant breakdown by ethnicity/race |
| Aschner 2012 Insulin glargine versus sitagliptin in insulin-naive patients with type 2 diabetes mellitus uncontrolled on metformin (EASIE): a multicentre, randomised open-label trial Aschner, P.; Chan, J.; Owens, D.R.; Picard, S.; Wang, E.; Dain, M. Lancet Jun 16 2012;379(9833):2262-2269 | No participant breakdown by ethnicity/race |
| Ahren 2004 Twelve- and 52-Week Efficacy of the Dipeptidyl Peptidase IV Inhibitor LAF237 in Metformin-Treated Patients With Type 2 Diabetes Ahren, B.; Mills, D.; Gomis, R.; Schweizer, A.; Standl, E. Diabetes Care 2004;27:2874-2880. | No participant breakdown by ethnicity/race |
