## Supplementary material for "Assessing non-white ethnic participation in type 2 diabetes mellitus randomized clinical trials: A Meta-Analysis": eTable 4

**eTable 4. Estimates used for PPR Calculations**

| **Variable** | **Estimate (%) of Population** | **Source** |
| --- | --- | --- |
| T2DM Prevalence in US | White: 7.5  Non-white: 12 | Centers for Disease Control and Prevention. National Diabetes Statistics Report, 2020. Atlanta, GA: Centers for Disease Control and Prevention, U.S. Dept of Health and Human Services; 2020. |
| Prevalence in UK | White: 5.0  Non-white: 6.8 | Pham TM, Carpenter JR, Morris TP, Sharma M, Petersen I. Ethnic Differences in the Prevalence of Type 2 Diabetes Diagnoses in the UK: Cross-Sectional Analysis of the Health Improvement Network Primary Care Database. *Clin Epidemiol*. 2019;11:1081-1088. doi:10.2147/CLEP.S227621 |
| Prevalence in Australia | White: 8.3  Non-white: 15.6 | Saeedi et al. (2019) – used estimates for Western Pacific |
| Prevalence in Japan | White: 8.3  Non-white: 15.6 | Saeedi et al. (2019) – used estimates for Western Pacific |
| Prevalence in Argentina | White: 6.7  Non-white: 11.3 | Saeedi et al. (2019) – used estimates for South America |
| Global Prevalence | White: 6.2  Non-white: 11.8 | Saeedi et al. (2019) |
| North America Prevalence | White: 9.0  Non-white: 14.5 | Saeedi et al. (2019) |
| Europe Prevalence | White: 4.9  Non-white: 9.2 | Saeedi et al. (2019) |
| Proportion of the population (US) | White: 60.1  Non-white: 39.9 | US Census Bureau. (2019). https://www.census.gov/quickfacts/fact/table/US/PST045219 |
| Proportion of the population (UK) | White: 86  Non-white: 14 | UK Census data, Government of the United Kingdom. (2011). https://www.ons.gov.uk/census/2011census |
| Proportion of the population (Australia) | White: 90.2  Non-white: 9.8 | People of Australia. Britannica. (2007). https://www.britannica.com/place/Australia |
| Proportion of the population (Argentina) | White: 85  Non-white: 15 | Fernandez FL. Composición Étnica de las Tres Áreas Culturales del Continente Americano al Comienzo del Siglo XXI" (PDF) (in Spanish). Centro de Investigación en Ciencias Sociales y Humanidades, UAEM. 2005. |
| Proportion of the population (Global) | White: 14.8  Non-white: 85.2 | Proportions were estimated by assuming that all people in Europe, Canada, the United States, and Australia are white  Population in millions (approximate):   - Europe: 746.4 - Canada: 37.59 - USA: 328.2 - Australia: 25.36 - Global: 7674   Source: United Nations World Population Prospects |
| Proportion of the population (North America) | White: 66.5  Non-white: 33.5 | Average of the percentage of white people in Canada, 72.9 (Statistics Canada 2016), and in the US, 60.1 (US Census Bureau 2019) |
| Proportion of the population (Europe) | White: 86  Non-white: 14 | UK census data estimates were also used for Europe estimates |
