## Supplementary material for "Assessing non-white ethnic participation in type 2 diabetes mellitus randomized clinical trials: A Meta-Analysis": eFigure 1

**eFigure 1. Consort flow diagram of the study selection process**

Records identified from*:

Databases (n = 3060)

Annals of Internal Medicine (n = 68)

Bmj (n= 64)

Circulation (n = 305)

Diabetes Care (n =2082)

JAMA (n = 144)

Lancet (n = 180)

NEJM (n= 219)

Records removed *before screening*:

Duplicate records removed (n = 0)

Records marked as ineligible by automation tools (n = 0)

Records removed for other reasons (n = 0)

Records screened

(n = 3060)

Records excluded** (n = 2487)

*Not RCT (n=318)*

*No pharmaceutical intervention (n=516)*

*Participants did not all have type 2 diabetes (n=1280)*

*Cell or animal study (n=4)*

*RCT with <100 participants (n=356)*

*RCT that enrolled children with type 2 diabetes (n=13)*

Reports sought for retrieval

(n = 573)

Reports not retrieved (n=0)

Reports assessed for eligibility

(n = 573)

Reports excluded:

*RCT with <100 participants (n=15)*

*Ancillary publication (n=105)*

*Industry trial in a journal with IF<30 (n=263)*

*Participants did not all have type 2 diabetes (n=13)*

*Not RCT of a T2DM pharmacotherapy (n=54)*

*Substudy (n=1)*

*One country for industry trial (n=11)*

*No participant breakdown by ethnicity/race (n=20)*

Studies included in review

(n =82)

- Industry-funded (n=68)
- Government-funded (n=14)

**Identification**

**Screening**

**Included**
