## Appendix A for "Assessing non-white ethnic participation in type 2 diabetes mellitus randomized clinical trials: A Meta-Analysis"

**Appendix A: PPR Calculation Process**

When a study recruited predominantly from the United States (US) (32) or United Kingdom (UK) (2), country-specific prevalence and demographic data was used to determine the denominator. Trials with Argentina (1) and Australia (1) as the country with the greatest participant recruitment also used country-specific demographic data.

To determine the expected number of diabetes cases for white participants in a study, prevalence and demographic data was used. For studies that recruited the most participants from the US, an estimate for white diabetes prevalence for the US from the Centre of Disease Control and Prevention website was used. This value was multiplied by the percentage of the US that is comprised of white people (retrieved from the US Census website). Then, the white prevalence percentage was multiplied by the total number of white people in the US to get an estimate of how many white people have diabetes in the US. The expected number of white cases was determined by dividing the number of white people with diabetes in the US by the total number of people with diabetes in the US (white and non-white).

Similarly, to determine the expected number of non-white diabetes cases in the US, the non-white diabetes prevalence value was multiplied by the percentage of the US that is comprised of non-white people. Then, the non-white prevalence percentage was multiplied by the total number of non-white people in the US to get an estimate of how many non-white people have diabetes in the US. The expected number of non-white cases was found by dividing the number of non-white people with diabetes in the US by the total number of people with diabetes in the US (non-white and white).

For trials that specified the *region* of greatest recruitment instead of a country, regional prevalence estimates were used. Diabetes prevalence data for the remaining trials was estimated using Table 1 from Saeedi et al.^94^ The study defines confidence intervals for the prevalence of diabetes in IDF regions. For the White and Non-White diabetes prevalence estimates, the lower bound of the confidence interval was taken to represent the White prevalence, and the upper bound of the confidence interval represented the Non-White prevalence. This was a reasonable approach for estimation given that the greatest prevalence of T2DM is observed in non-white ethnic groups. For trials with no indication of how many participants were recruited from each region, worldwide prevalence estimates from Saeedi et al. were used. Demographic data about ethnic breakdown in specific countries (US, UK, Argentina, Australia) was retrieved from online sources (eTable 4 in the Supplement)

When demographic data about ethnicity was unavailable for regions (Europe, North America, Worldwide), it was estimated. For trials that recruited primarily from Europe (16), demographic data from the UK was used. For North American trials (5), ethnicity data from the US and Canada was combined and averaged. For trials with no participant recruitment data by region/country (19), the proportion of white and non-white people worldwide was estimated by assuming that all people in Europe, Canada, the US, and Australia are white, resulting in an estimate of 14.8% white people and 85.2% non-white people.

For trials that recruited only from North America and Europe (3) and South America and North America (1), it was assumed that 50% of the recruitment was from each respective region. As such, the prevalence values for each region from Saeedi et al were averaged and the total population of each region was added to calculate the PPR. For trials that had 0 white participants or 0 non-white participants, a 0.5 correction was used in place of “0” for calculation purposes.
