## Appendix B for "Assessing non-white ethnic participation in type 2 diabetes mellitus randomized clinical trials: A Meta-Analysis"

**Appendix B: Sensitivity analysis with varying worldwide population proportions**

Variation 1: Worldwide population proportion is 90% non-white and 10% white.

**eFigure 2a: Sensitivity Analysis 90/10 non-white/white comparison – White PPR Industry Trials**

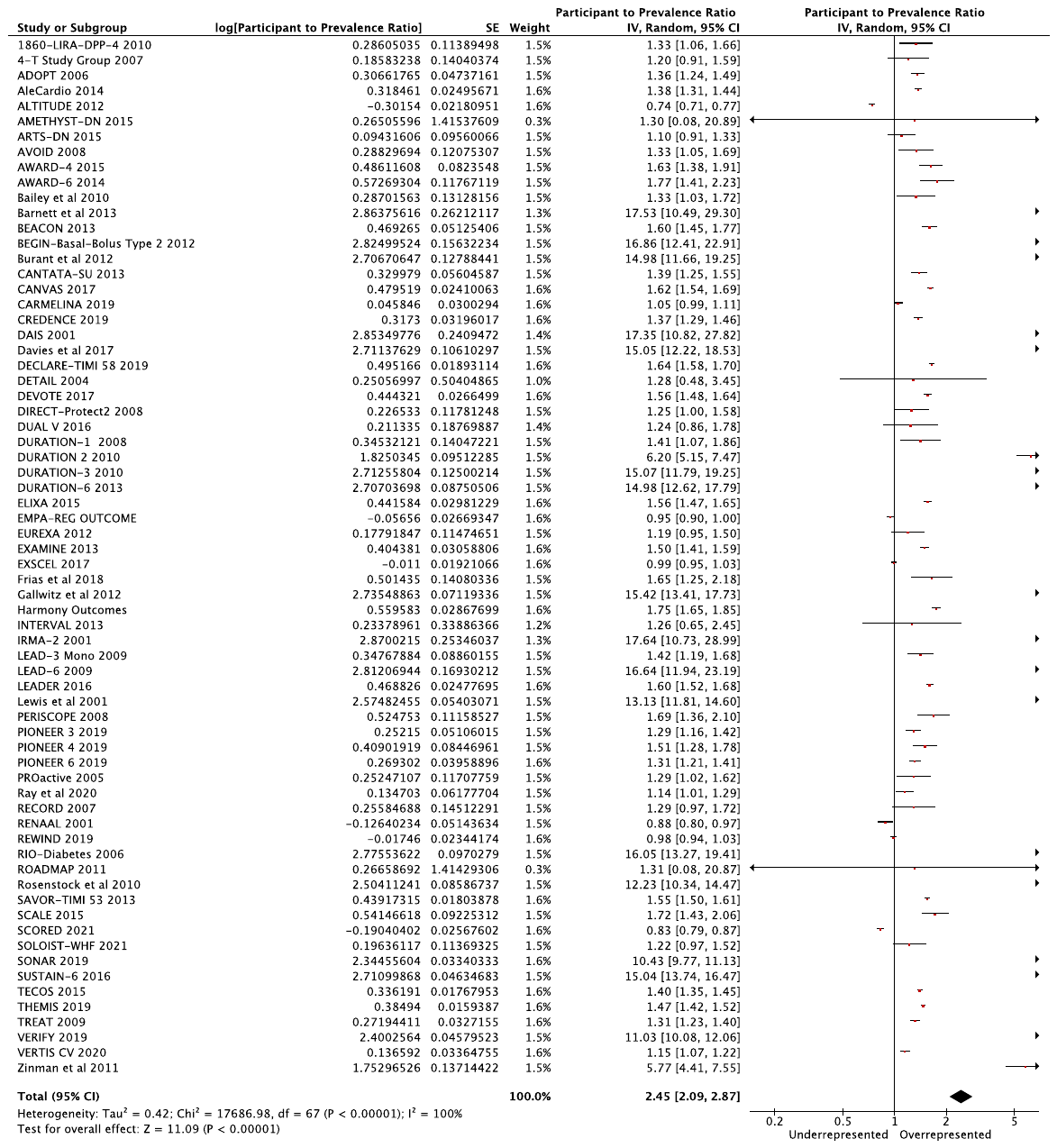

**eFigure 2b: Sensitivity Analysis 90/10 non-white/white – Non-white PPR Industry Trials**

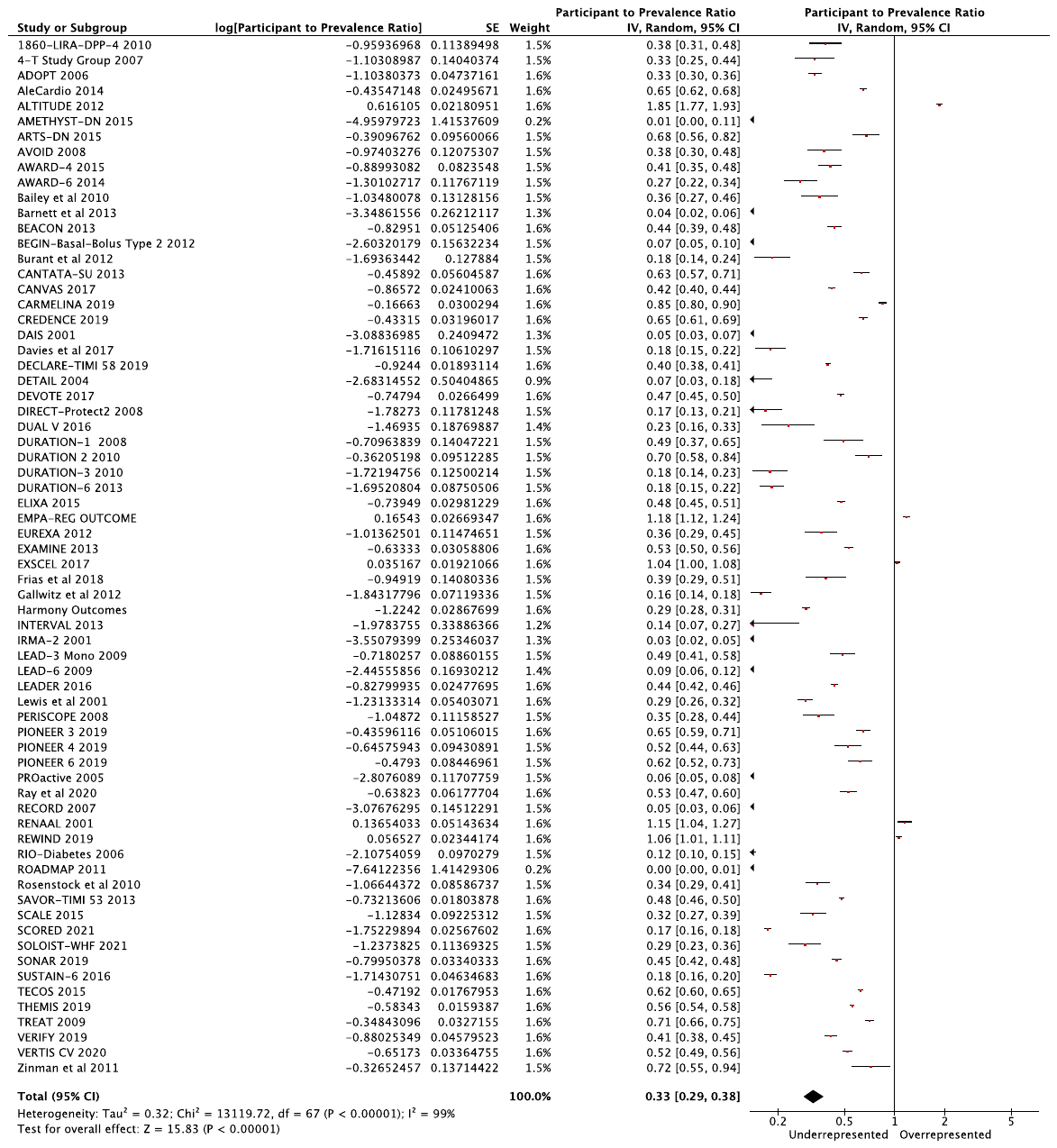

Variation 2: Worldwide population proportion is 87.5% non-white, 12.5% white

**eFigure 3a: Sensitivity Analysis 87.5/12.5 non-white/white – White PPR Industry Trials**

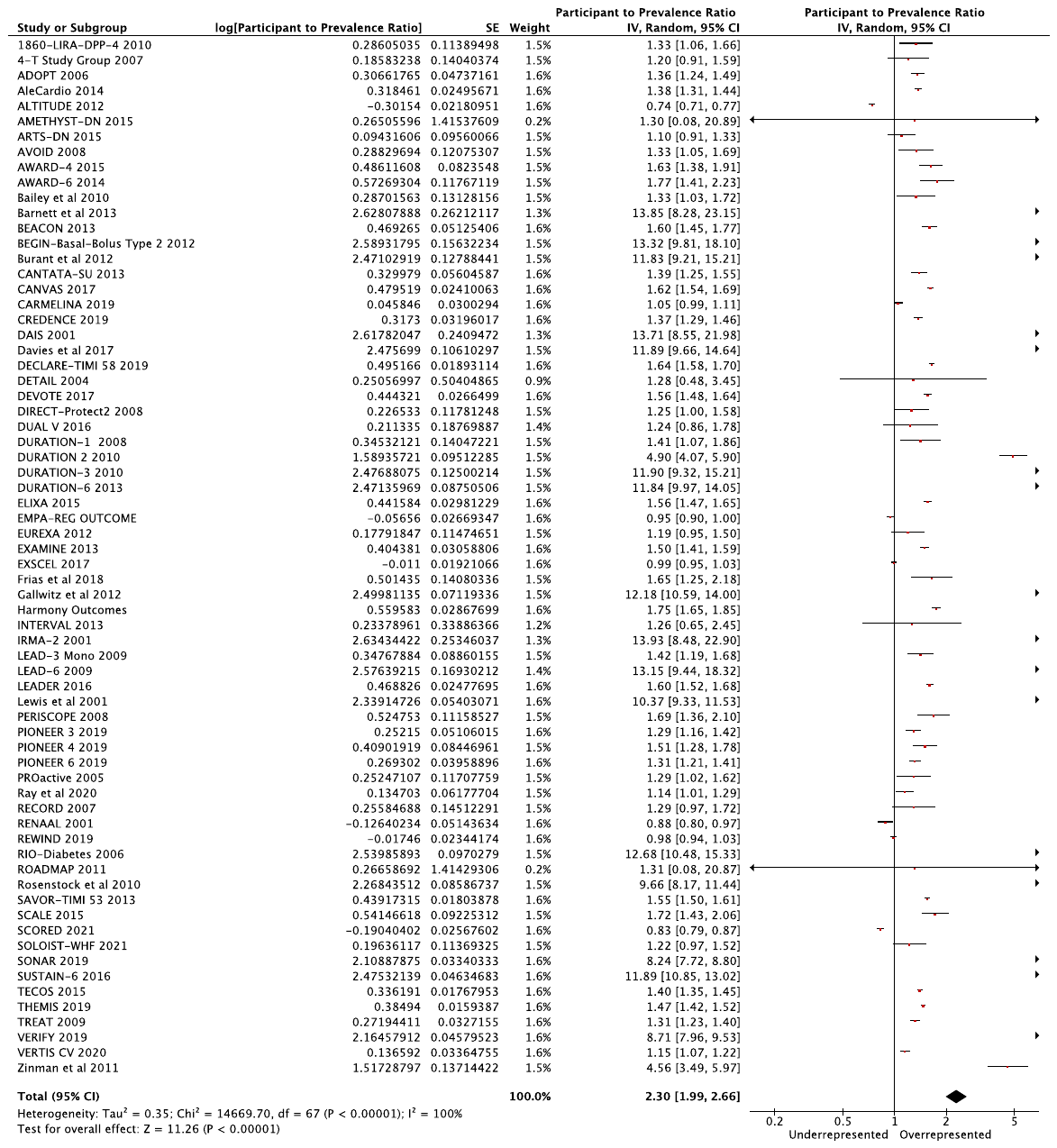

**eFigure 3b: Sensitivity Analysis 87.5/12.5 non-white/white – Non-white PPR Industry Trials**

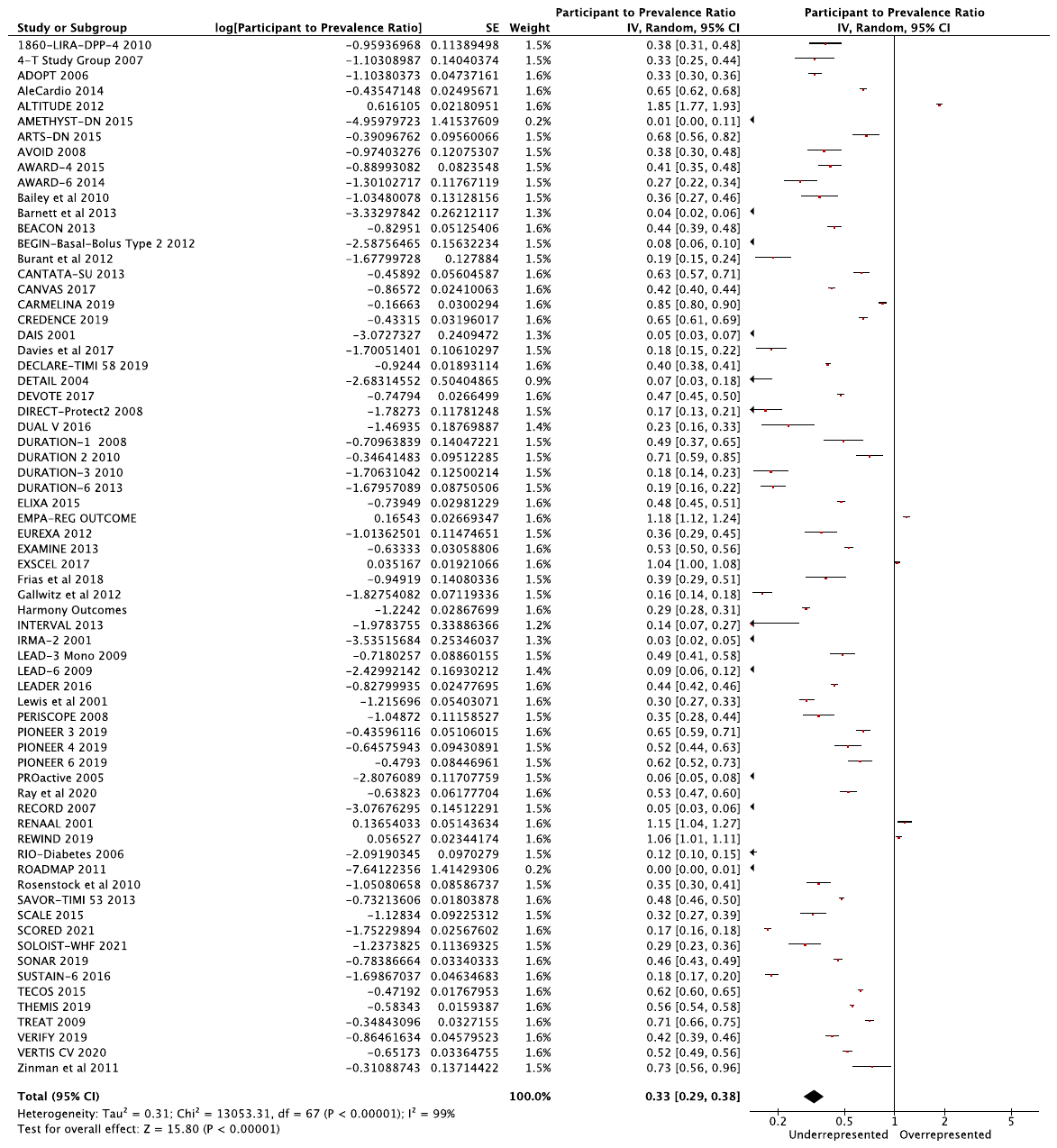

Variation 3: Worldwide population proportion is 85% non-white, 15% white

**eFigure 4a: Sensitivity Analysis 85/15 non-white/white – White PPR Industry Trials**

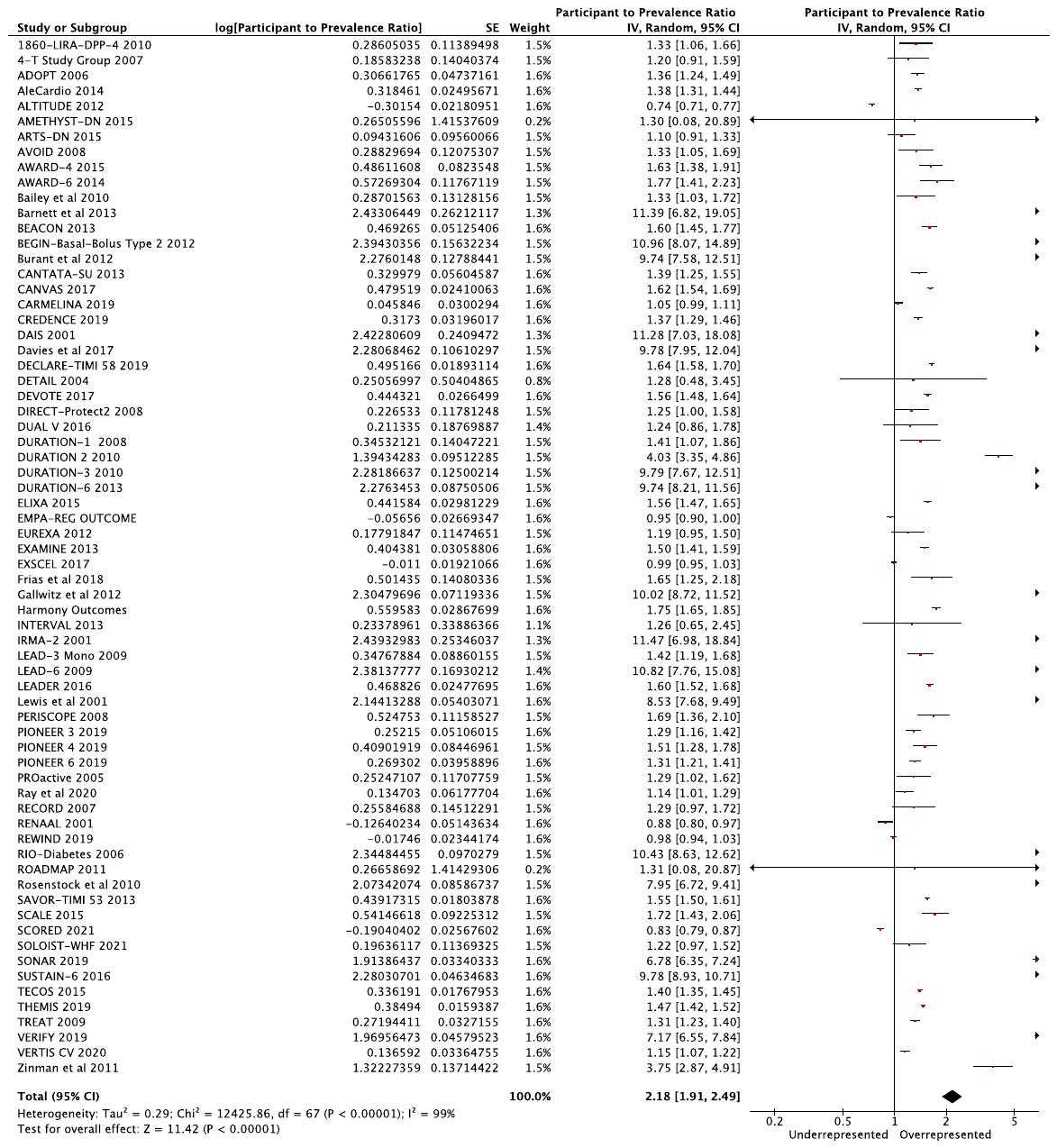

**eFigure 4b: Sensitivity Analysis 85/15 non-white/white – Non-white PPR Industry Trials**

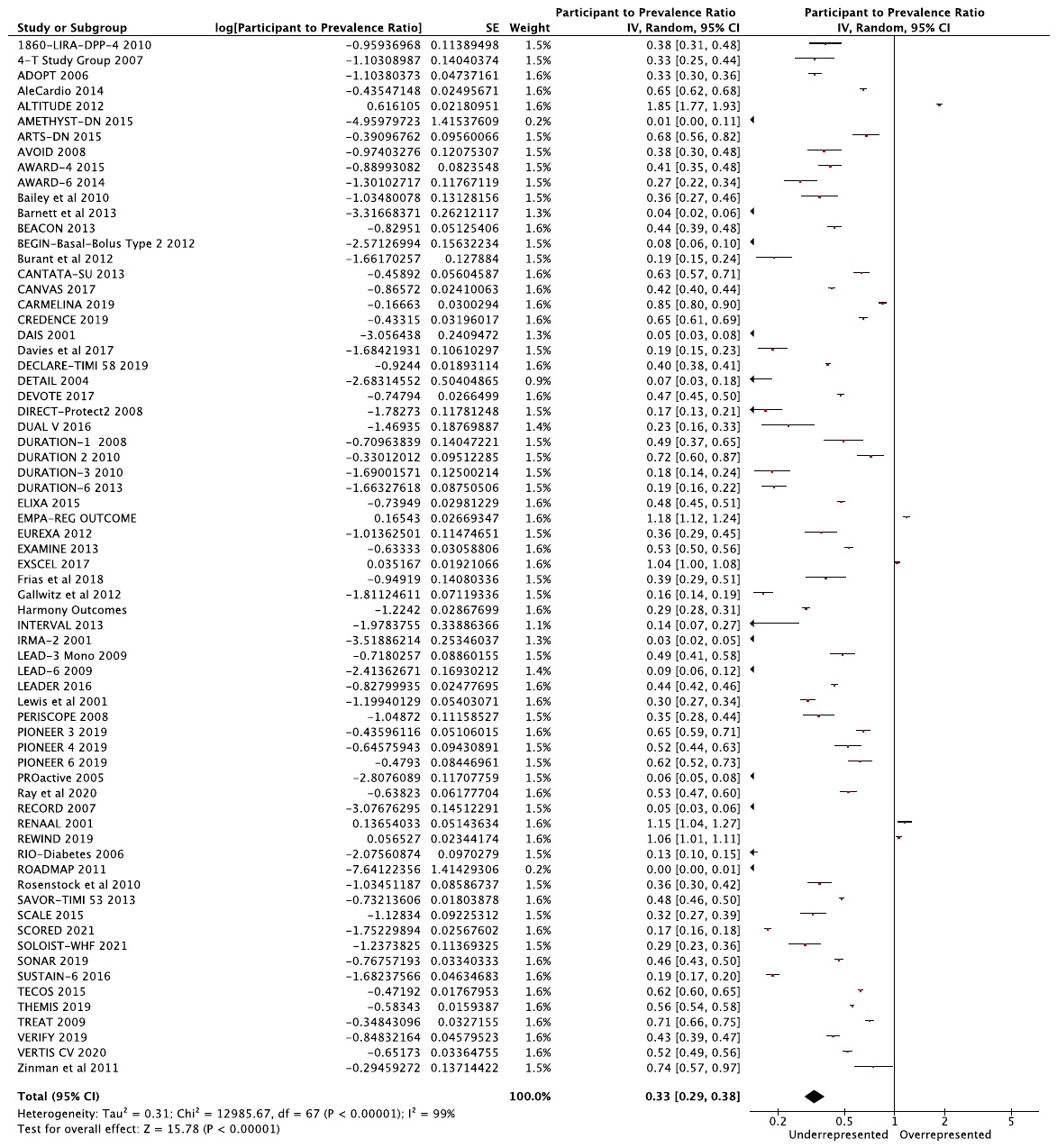

Variation 4: Worldwide population proportion is 80% non-white, 20% white

**eFigure 5a: Sensitivity Analysis 80/20 non-white/white – White PPR Industry Trials**

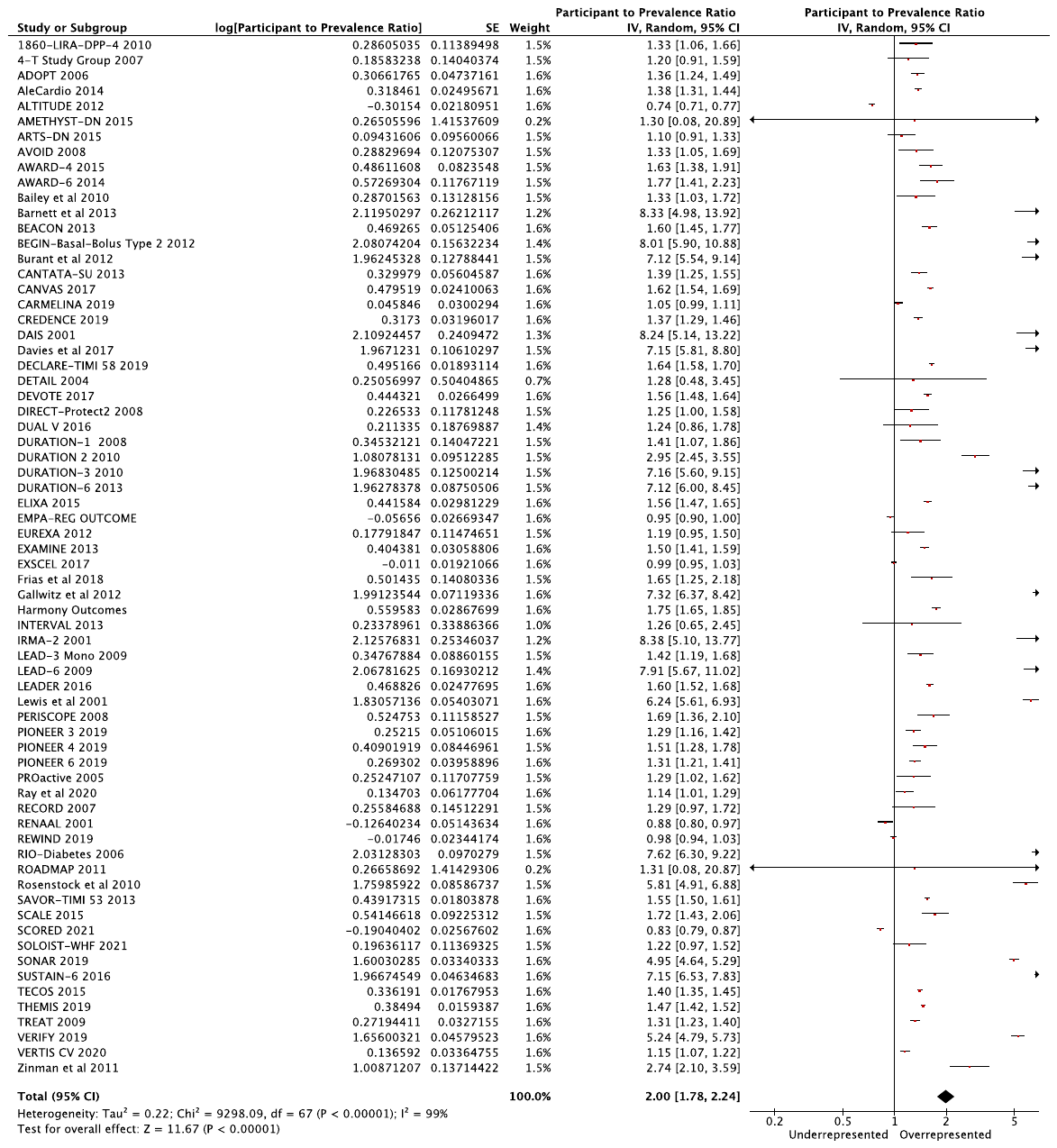

**eFigure 5b: Sensitivity Analysis 80/20 non-white/white – Non-white PPR Industry Trials**

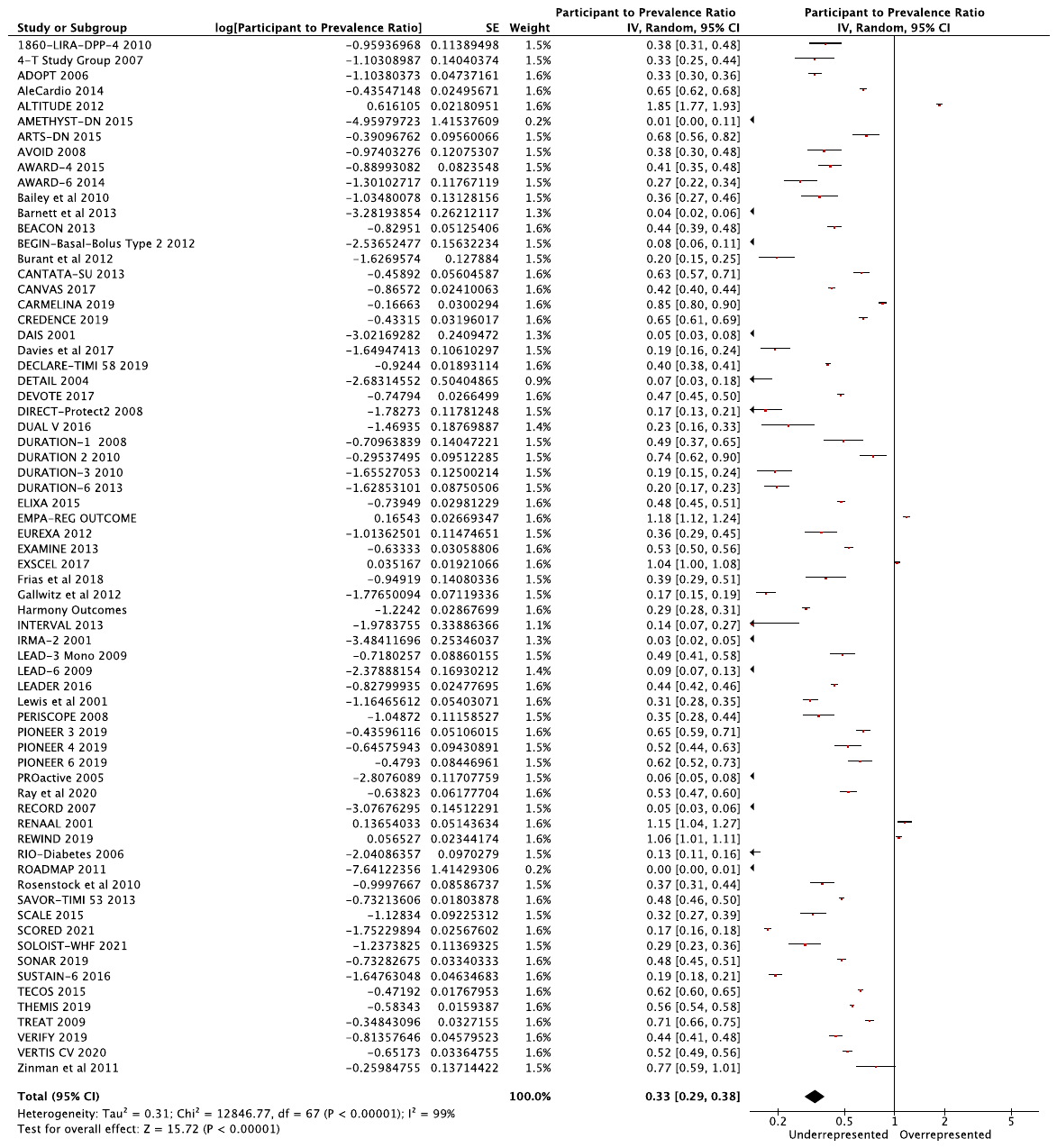
